# Mortality rates among hospitalized patients with COVID-19 treated with convalescent plasma A Systematic review and meta-analysis

**DOI:** 10.1101/2023.01.11.23284347

**Authors:** Jonathon W. Senefeld, Ellen K. Gorman, Patrick W. Johnson, M. Erin Moir, Stephen A. Klassen, Rickey E. Carter, Nigel S. Paneth, David J. Sullivan, Olaf H. Morkeberg, R. Scott Wright, DeLisa Fairweather, Katelyn A. Bruno, Shmuel Shoham, Evan M. Bloch, Daniele Focosi, Jeffrey P. Henderson, Justin E. Juskewitch, Liise-anne Pirofski, Brenda J. Grossman, Aaron A.R. Tobian, Massimo Franchini, Ravindra Ganesh, Ryan T. Hurt, Neil E. Kay, Sameer A. Parikh, Sarah E. Baker, Zachary A. Buchholtz, Matthew R. Buras, Andrew J. Clayburn, Joshua J. Dennis, Juan C. Diaz Soto, Vitaly Herasevich, Allan M. Klompas, Katie L. Kunze, Kathryn F. Larson, John R. Mills, Riley J. Regimbal, Juan G. Ripoll, Matthew A. Sexton, John R.A. Shepherd, James R. Stubbs, Elitza S. Theel, Camille M. van Buskirk, Noud van Helmond, Matthew N.P. Vogt, Emily R. Whelan, Chad C. Wiggins, Jeffrey L. Winters, Arturo Casadevall, Michael J. Joyner

## Abstract

**IMPORTANCE:** Many hospitalized patients with COVID-19 have been treated with convalescent plasma. However, it is uncertain whether this therapy lowers mortality and if so, if the mortality benefit is larger among specific subgroups, such as recipients of plasma with high antibody content and patients treated early in the disease course.

**OBJECTIVE:** To examine the association of COVID-19 convalescent plasma transfusion with mortality and the differences between subgroups in hospitalized patients with COVID-19.

**DATA SOURCES:** On October 26, 2022, a systematic search was performed for clinical studies of COVID-19 convalescent plasma in the literature.

**STUDY SELECTION:** Randomized clinical trials and matched cohort studies investigating COVID-19 convalescent plasma transfusion compared with standard of care treatment or placebo among hospitalized patients with confirmed COVID-19 were included. The electronic search yielded 3,841 unique records, of which 744 were considered for full-text screening. The selection process was performed independently by a panel of five reviewers.

**DATA EXTRACTION AND SYNTHESIS:** The study followed the Preferred Reporting Items for Systematic Reviews and Meta-Analyses (PRISMA) guidelines. Data were extracted by 5 independent reviewers in duplicate and pooled using inverse-variance random-effects model.

**MAIN OUTCOMES AND MEASURES:** Prespecified end point was all-cause mortality during hospitalization.

**RESULTS:** Thirty-nine randomized clinical trials enrolling 21,529 participants and 70 matched cohort studies enrolling 50,160 participants were included in the systematic review. Separate meta-analyses demonstrated that transfusion of COVID-19 convalescent plasma was associated with a significant decrease in mortality compared with the control cohort for both randomized clinical trials (odds ratio (OR), 0.87 [95% CI, 0.76-1.00]) and matched cohort studies (OR, 0.77 [95% CI, 0.64-0.94]). Meta-analysis of subgroups revealed two important findings. First, treatment with convalescent plasma containing high antibody levels was associated with a decrease in mortality compared to convalescent plasma containing low antibody levels (OR, 0.85 [95% CI, 0.73 to 0.99]). Second, earlier treatment with COVID-19 convalescent plasma was associated with a significant decrease in mortality compared with the later treatment cohort (OR, 0.63 [95% CI, 0.48 to 0.82]).

**CONCLUSIONS AND RELEVANCE:** COVID-19 convalescent plasma use was associated with a 13% reduced risk in mortality, implying a mortality benefit for hospitalized patients with COVID-19, particularly those treated with convalescent plasma containing high antibody levels treated earlier in the disease course.

**Key Points:** 

**Question:** What is the evidence regarding the potential mortality benefit associated with transfusion of convalescent plasma in hospitalized patients with COVID-19?

**Findings:** In this meta-analysis of 39 randomized clinical trials enrolling 21,529 participants and 70 matched cohort studies enrolling 50,160 participants, transfusion of convalescent plasma was associated with a 13% mortality benefit. Subgroup analyses revealed that patients treated with plasma containing higher levels of antibodies and patients treated earlier in the course of the disease had a greater mortality benefit associated with COVID-19 convalescent plasma transfusion.

**Meaning:** These findings suggest that transfusion of COVID-19 convalescent plasma is associated with a mortality benefit for hospitalized patients, particularly those treated earlier in the disease course.

## Introduction

The COVID-19 pandemic created a humanitarian crisis that prompted an expeditious search for safe and effective COVID-19 therapies. Before identifying effective antispike monoclonal antibodies and small molecule antivirals, COVID-19 convalescent plasma was proposed as a safe treatment with a promising efficacy profile in early reports.^1-3^ More recently, as new SARS-CoV-2 variants emerged and evaded antispike monoclonal antibodies^4,5^, interest has been renewed in understanding the clinical efficacy of COVID-19 convalescent plasma^6^, particularly among patients who are immunocompromised.^7^ Although COVID-19 convalescent plasma has been widely available and used to treat over half-a-million patients with COVID-19^8^, uncertainty remains about the utility of COVID-19 convalescent plasma and its association with mortality due to heterogenous findings from individual studies.^9^ Heterogeneity in clinical studies is likely due to several key factors: biological diversity of COVID-19 convalescent plasma, evolving and nonstandard treatment protocols, and a wide spectrum of clinical use of COVID-19 convalescent plasma from postexposure prophylaxis to therapy of last resort in patients with multiorgan failure. In this framework, many different approaches have been used to study the mortality of patients with COVID-19 treated with COVID-19 convalescent plasma, including randomized clinical trials (RCTs)^10-48^, real-time pooling of individual patient data from RCTs^6,49^, and meta-analyses.^41,50-53^ The present work was performed to provide an updated, high-quality systematic review and meta-analysis on the use of COVID-19 convalescent plasma.

This systematic review and meta-analysis aimed to evaluate the association of COVID-19 convalescent plasma with mortality among hospitalized patients with COVID-19 by pooling data from RCTs and matched cohort studies. Moreover, prespecified analyses aimed to determine if the potential association between COVID-19 convalescent plasma and mortality benefit differs across patient subgroups based on anti-SARS-CoV-2 antibody levels within COVID-19 convalescent plasma and the timing of the convalescent plasma transfusion in relation to the disease course.^54^ By integrating information from many studies and lines of evidence, we hope that this work provides new insights, clarifies ambiguous areas, and removes biases from the scientific corpus.^55^

## Methods

This systematic review and meta-analysis followed the recommendations in the *Cochrane Handbook for Systematic Review of Interventions*^56^ and reported findings according to the Preferred Reporting Items for Systematic Reviews and Meta-analyses (<u>PRISMA</u>) reporting guidelines. The study protocol has been registered in PROSPERO (<u>CRD42022316321</u>); all changes to the protocol are reported in the Methods section. In accordance with the Code of Federal Regulations, 45 CFR 46.102, this study was exempt from obtaining institutional review board approval from Mayo Clinic and the requirement to obtain informed patient consent because it is secondary use of publicly available data sets.

### Eligibility Criteria

Eligible patients were hospitalized with COVID-19. The intervention investigated was transfusion with COVID-19 convalescent plasma of any dosage. The control group was treated with standard of care according to local treatment guidelines, with or without a placebo. The primary outcome was all cause mortality during hospitalization. Randomized clinical trials and matched cohort studies were eligible for all analyses. For subgroup analyses, where the focus is on differences among patients treated with COVID-19 convalescent plasma, case series were also eligible.

### Information Sources

On October 26, 2022, PubMed, MEDLINE, Google Scholar, and medRχiv were searched for eligible studies published beginning with January 1, 2020— approximating the origins of the COVID-19 pandemic. Keywords used in the search included ((convalescent plasma) OR (convalescent serum)) AND COVID-19 (and medical subject headings).

### Selection and Data Collection Processes

Both the selection process and data collection process were performed in duplicate and independently by two reviewers from a cohort of five potential reviewers (J.W.S., E.K.G., M.E.M., S.A.K., O.H.M.), and all data were independently verified by review from a third reviewer. Disagreements were discussed until consensus. Data abstraction was performed using a standardized data abstraction form. Abstracted data included patient demographic characteristics (sample size, age, sex, need for mechanical ventilation at the time of COVID-19 convalescent plasma transfusion) and COVID-19 convalescent plasma transfusion characteristics (volume transfused, antibody level, and time to transfusion in relation to disease course) as available. Data were abstracted corresponding to the latest available follow-up time for mortality. Further information on the selection process is presented in Figure 1.

**FIGURE 1.**
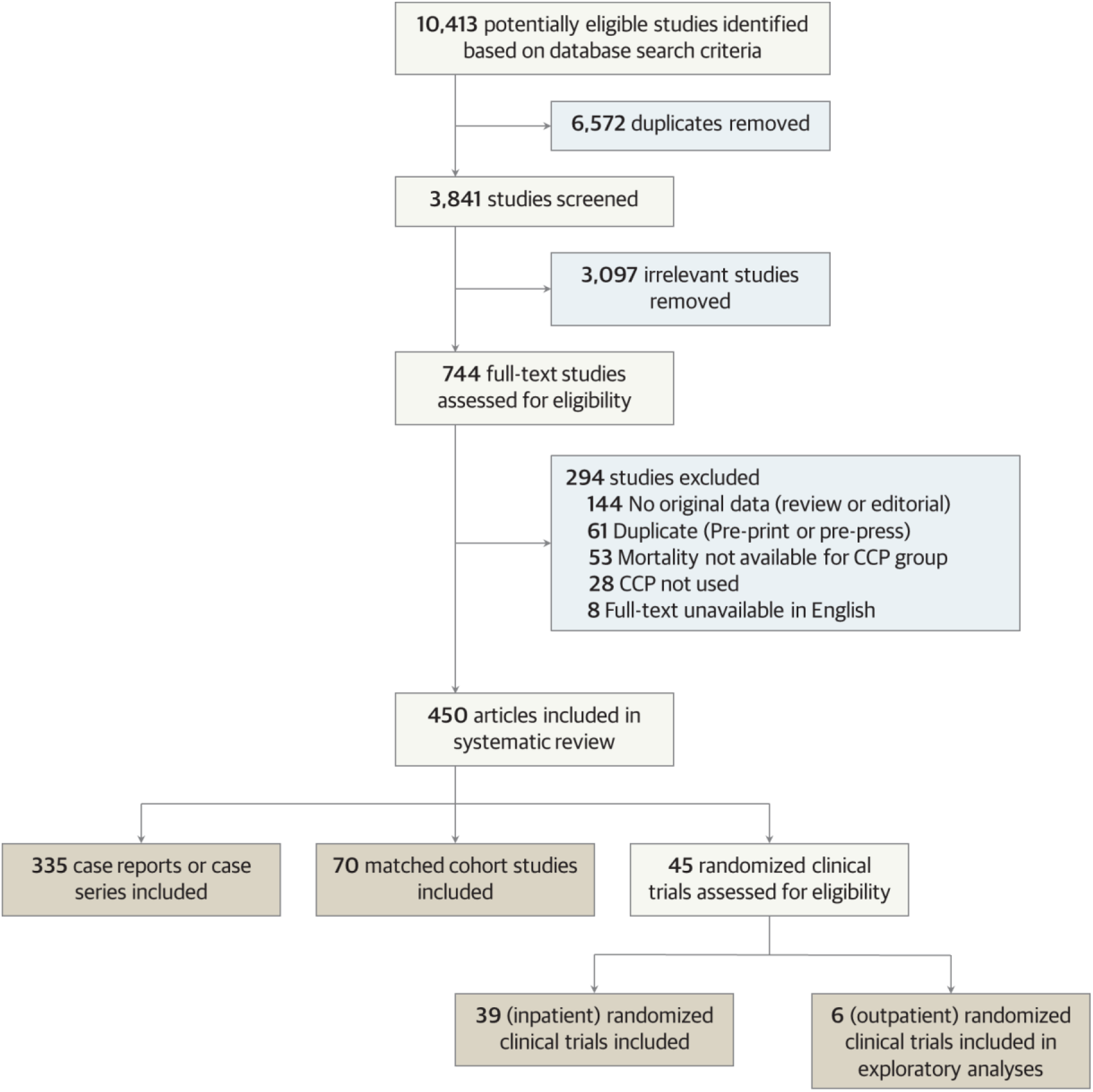
PRISMA flow diagram. Flow diagram displaying the selection of articles through different phases of the systematic review, and categorization of included articles.

### Study Risk of Bias Assessment

A risk of bias assessment was conducted using the Cochrane Risk of Bias 2.0 Tool for randomized clinical trials^57,58^ and the Newcastle-Ottawa Scale for matched cohort studies. Two reviewers from a cohort of three potential reviewers (J.W.S., E.K.G., S.A.K.) applied the risk of bias assessment independently, and verified by a third reviewer. Discrepancies were discussed until a consensus was reached.

### Statistical Analysis

For the primary, dichotomous outcome of mortality, we performed a meta-analysis using a random-effects model. We extracted raw data on mortality events and the number of patients for each group. For each study, we compared the observed number of deaths among patients transfused with COVID-19 convalescent plasma with the expected number if all patients were at equal risk using standard formulae for 2×2 contingency tables. Trials results were combined and weighted using an inverse-variance model. Analyses were done using Comprehensive Meta-analysis software (CMA 2.0, Biostat, Englewood, USA). Results are reported with 95% confidence intervals (CIs), statistical significance was set at α = .05, and all tests were 2-tailed.

Primary meta-analyses were performed separately for randomized clinical trials and matched cohort studies. We performed prespecified subgroup analyses, including a subgroup analysis based on timing of convalescent plasma transfusion (early vs. late treatment) in relation to the COVID-19 disease course and a subgroup analysis based on antibody concentration in the transfused COVID-19 convalescent plasma (high vs. low antibody levels).

Additionally, an exploratory meta-analysis was performed on hospitalization rates among outpatients with recent SARS-CoV-2 exposure or infection.

## Results

### Study Selection and Characteristics

The process of study selection is represented in the PRISMA flow diagram **(Figure 1)**. Thirty-nine randomized clinical trials^10-48^ enrolling 21,529 participants and 70 matched cohort studies^2,59-127^ enrolling 50,160 participants were included in the primary analyses. Although controlled studies were the focus of this systematic review and meta-analysis, secondary analyses on COVID-19 convalescent plasma antibody levels and timing of COVID-19 convalescent plasma transfusion encompass findings from case series, as delineated below.

### Risk Assessment

The results of the risk of bias assessment for randomized clinical trials and matched cohort studies are presented in **eTable 2** and **eTable 3**, respectively, in the Supplement. Because our analyses primarily included controlled trials and focused on a discrete, dichotomous outcome that is unlikely to be influenced by implicit biases of research personnel (all-cause mortality), many studies were determined to have low risk of bias. Matched cohort studies were associated with higher risk of bias because of the open-label trial design.

### Association Between Convalescent Plasma Transfusion and Mortality in Hospitalized Patients with COVID-19

Key findings of the primary meta-analysis of 39 randomized clinical trials including 11,303 patients treated with COVID-19 convalescent plasma and 10,226 patients treated with usual care (controls) are displayed in **Figure 2**. Treatment with COVID-19 convalescent plasma was associated with a 13% reduced risk in mortality rates compared to usual care, with a pooled risk ratio estimate of 0.87 (95% CI, 0.76 to 1.00).

**FIGURE 2.**
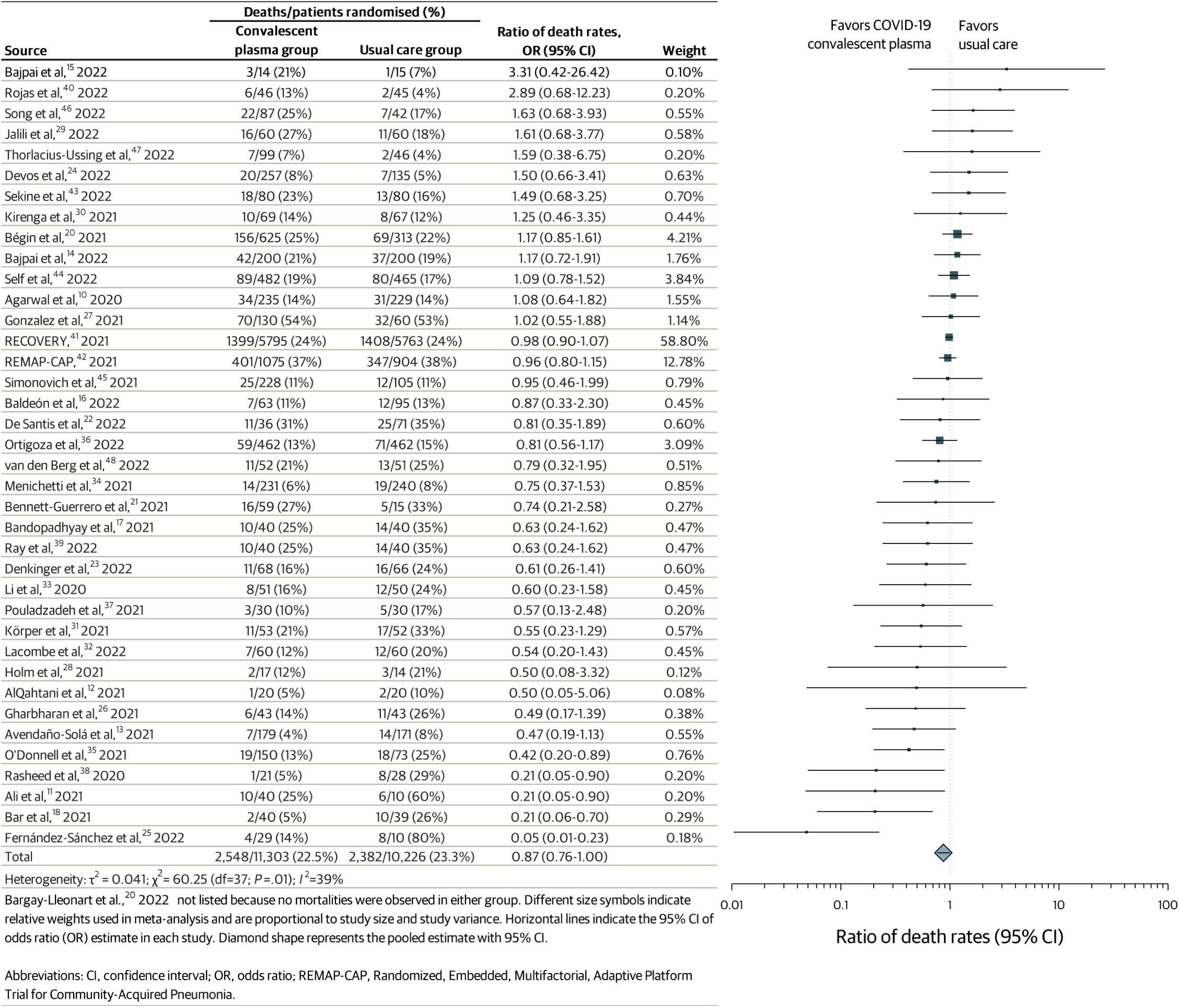
Forest plot of mortality among randomized clinical trials.

The clinical benefit associated with COVID-19 convalescent plasma observed in 39 randomized clinical trials was also supported by meta-analysis of 70 matched cohort studies, which included 14,541 patients treated with COVID-19 convalescent plasma and 35,619 patients treated with usual care (controls). In this meta-analysis of matched cohort studies, treatment with COVID-19 convalescent plasma was associated with a 23% reduced risk in mortality rates compared to usual care, with a pooled risk ratio estimate of 0.77 (95% CI, 0.64 to 0.94). A forest plots associated with these findings is displayed in **eFigure 1** in the Supplement. Although there was heterogeneity between individual studies, there was a high level of concordance between these two separate meta-analyses.

### Exploratory Analyses of Outpatient COVID-19 Convalescent Plasma

Six randomized clinical trials^128-133^ enrolling 2,824 participants were included in an exploratory analysis of hospitalization or mortality rates among outpatients with recent SARS-CoV-2 exposure or infection. In this meta-analysis, treatment with COVID-19 convalescent plasma among outpatients was associated with a 35% decrease in hospitalization rate, with a pooled risk ratio estimate of 0.65 (95% CI, 0.45 to 0.94). Results of this exploratory meta-analysis are shown in the forest plot in **Figure 3**. However, there was no apparent mortality benefit associated with convalescent plasma among outpatients, with a pooled risk ratio estimate of 0.60 (95% CI, 0.16 to 2.28), **eFigure 2**.

**FIGURE 3.**
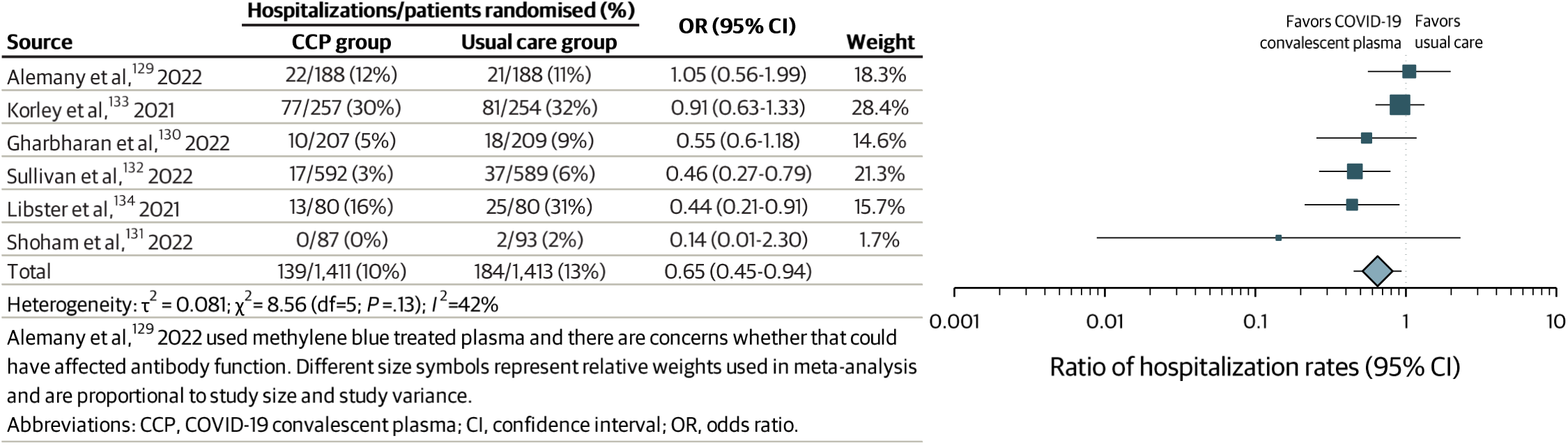
Forest plot of hospitalization among outpatients with recent SARS-CoV-2 exposure or infection in randomized clinical trials.

### Subgroup Analyses

#### COVID-19 convalescent plasma antibody levels

Among patients treated with COVID-19 convalescent plasma, receipt of convalescent plasma with higher levels of antibodies has been suggested to be associated with reduced mortality.^20,54^ Thus, heterogeneity of antibody levels in the COVID-19 convalescent plasma used to treated patients may impact the mortality rates reported. Hence, we examined within-study mortality rates among patients treated with convalescent plasma containing high or low antibody levels. Because several assays were authorized for use in the manufacture of high antibody-titer convalescent plasma and cutpoints used to qualify high antibody-titer convalescent plasma have changed over time, we used study-defined cutpoints to define high and low antibody levels. Information on assay systems and cutpoints used to delineate high and low antibody levels are provided in **eTable 4** in the Supplement.

In this framework, three matched cohort studies ^98,109,114^ enrolling 330 participants and 12 case series ^54,134-144^ enrolling 29,361 participants were included in this subgroup analysis. In this subgroup meta-analysis, treatment with COVID-19 convalescent plasma containing high antibody levels was associated with a 15% decrease in mortality rates compared to convalescent plasma containing low antibody levels, with a pooled risk ratio estimate of 0.85 (95% CI, 0.73 to 0.99). Results of this subgroup meta-analysis are shown in the forest plot in **Figure 4**.

**FIGURE 4.**
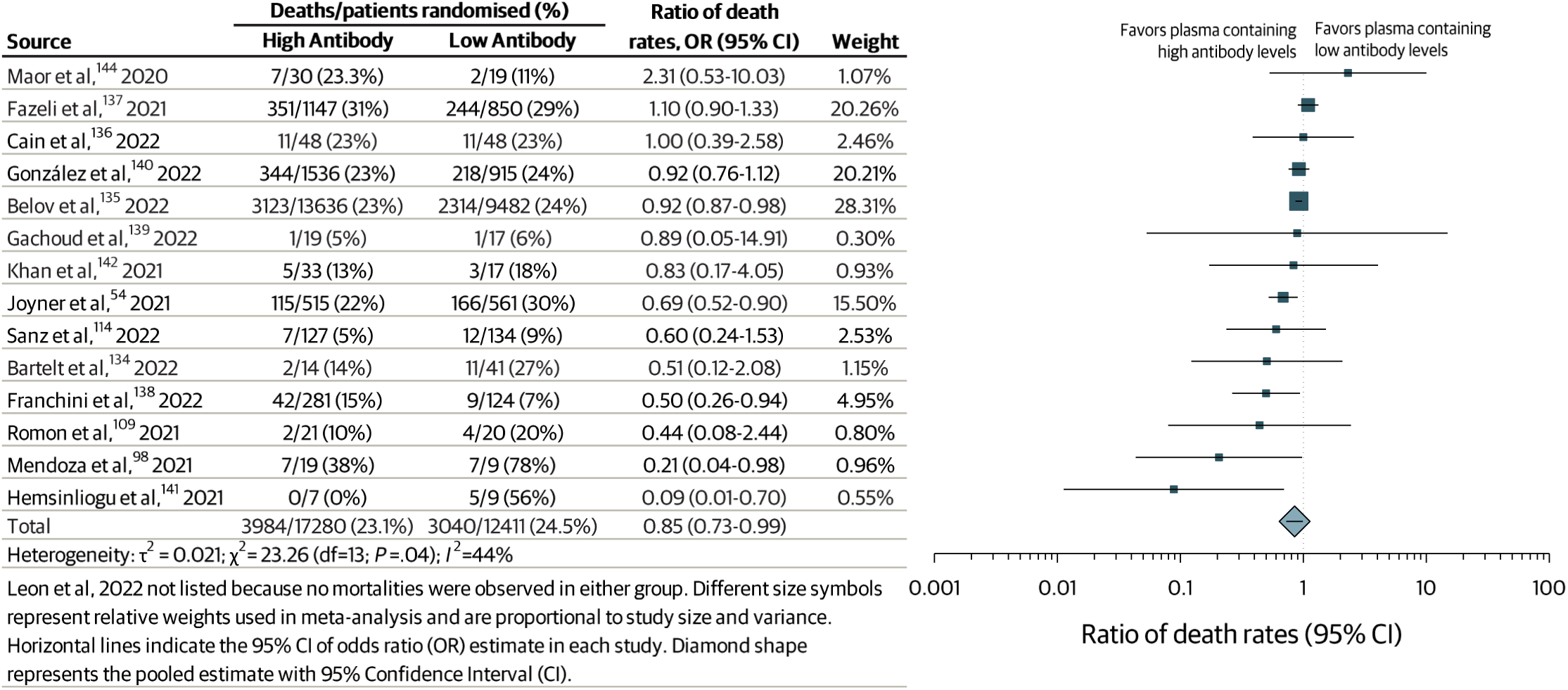
Forest plot of mortality among studies investigating COVID-19 convalescent plasma containing high compared to low antibody levels.

#### Timing of COVID-19 convalescent plasma transfusion

Among hospitalized patients transfused with COVID-19 convalescent plasma, transfusion earlier in the COVID-19 disease course has been suggested to be associated with reduced mortality^52,54,72^, thus, heterogeneity of the time between COVID-19 diagnosis and convalescent plasma transfusion may impact mortality rates. Because there was no standard to define ‘early treatment’ and individual studies used diverse criteria to define ‘early treatment’, we used study-defined cutpoints to delineate early and late treatment with COVID-19 convalescent plasma. Information on cutpoints used to define earlier and later treatment with COVID-19 convalescent plasma are provided in **eTable 5** in the Supplement.

In this context, we examined within study mortality rates among patients treated with convalescent plasma earlier compared to those treated later in the COVID-19 disease course. This subgroup analysis examined 26 studies, including: 2 randomized clinical trials ^14,41^ enrolling 5,990 participants, 7 matched cohort studies ^69,72,97,101,109-111^ enrolling 1,928 participants, and 17 case series ^136-138,140,145-157^ enrolling 10,530 participants. In this subgroup meta-analysis, treatment with convalescent plasma earlier in the COVID-19 disease course was associated with a 37% decrease in mortality rates compared to treatment later in the disease course, with a pooled risk ratio estimate of 0.63 (95% CI, 0.48 to 0.82). Results of this subgroup meta-analysis are shown in the forest plot in **Figure 5**.

**FIGURE 5.**
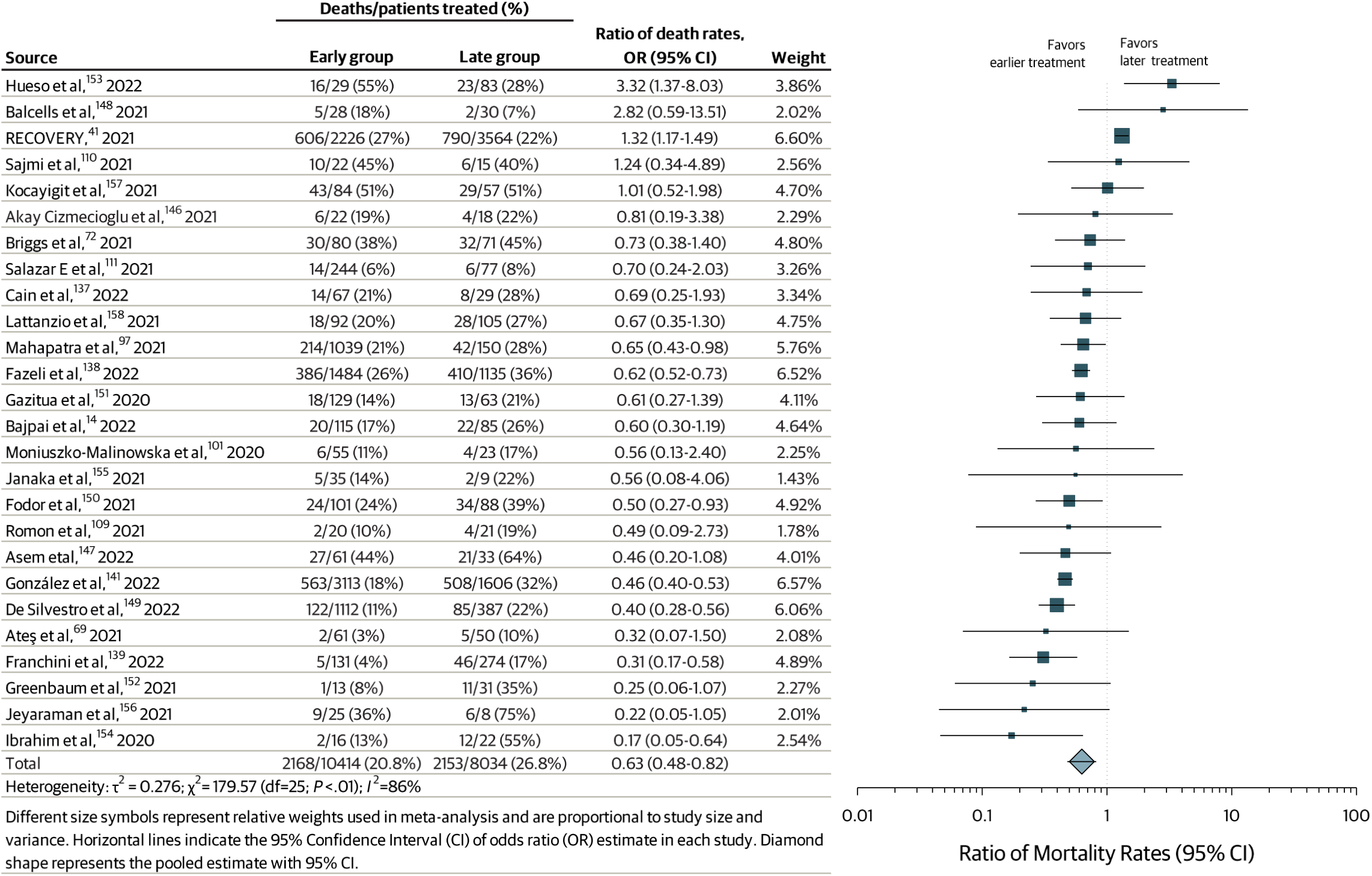
Forest plot of mortality among studies investigating earlier compared to later transfusion of COVID-19 convalescent plasma.

#### Case series and reports

In the United States and many other countries, the regulatory framework during the COVID-19 pandemic enabled broad access to COVID-19 convalescent plasma.^158^ In this context, a large number of single-arm studies evaluated the risk of death from COVID-19 after transfusion with convalescent plasma, including over 300 case series or case reports. The case series and case reports treated 135,949 participants with COVID-19 convalescent plasma and mortality was observed in 33,771 participants (∼25% mortality rate). Study level data are provided in **eTable 5** in the Supplement, and references associated with these case series and case reports are provided in the Supplementary References.

## Discussion

This systematic review and meta-analysis of 39 randomized clinical trials and 70 matched cohort studies of use of COVID-19 convalescent plasma for hospitalized patients with COVID-19 provides new insights into the therapeutic use of convalescent plasma and removes biases from the scientific and clinical literature. This meta-analysis found that convalescent plasma was associated with a clinically meaningful mortality benefit among hospitalized patients with COVID-19. However, given the large diversity of the included trials and heterogeneity of findings, the results must be analyzed and interpreted critically.

Overall, COVID-19 convalescent plasma transfusion was associated with a 13% decrease in mortality rates compared with the (usual care) control group. Subgroup analyses revealed that patients treated with high-titer plasma and patients treated earlier in the course of the disease benefited more from COVID-19 convalescent plasma transfusion than patients treated with lower-titer plasma or patients treated later in the course of the disease. Exploratory analyses also demonstrated that transfusion of COVID-19 convalescent plasma among outpatients was associated with a 35% decrease in hospitalization rates compared with the control group— a finding which is consistent with pooled individual patient data.^159^ Our finding that convalescent plasma transfusion reduced mortality is consistent with the epidemiologic data showing an inverse correlation between plasma use and COVID-19 death in the USA.^8^

Although data were directionally consistent when pooling randomized clinical trials and matched cohort studies, findings of individual trials were heterogenous. This heterogeneity between trials is likely associated with several factors. First, clinical heterogeneity may be based on patient characteristics at the time of COVID-19 convalescent plasma therapy. It is important to consider COVID-19 disease severity, which is directly related to higher odds of death. Among studies in which the patients had more severe disease, mortality was higher and there was often no clinical benefit associated with COVID-19 convalescent plasma transfusion.^160^ This meta-analysis found that treatment with COVID-19 convalescent plasma earlier in the course of the disease was associated with a 37% decrease in mortality rates. Thus, treatment later in the course of the disease may not confer a mortality benefit, unless the patients are immunocompromised.^161,162^

Second, clinical heterogeneity may result from differences in the COVID-19 convalescent plasma intervention itself, particularly the antibody content and geographic provenance of plasma supplies.^163^ Several assays are authorized for use in the manufacture of high antibody-titer convalescent plasma and there are established, assay-specific cutpoints used to define high antibody-titer convalescent plasma. However, there is discordance between assays and the cutpoints to define high antibody-titer convalescent plasma are not directly comparable across assays.^164^ Additionally, the cutpoints used to qualify high antibody-titer convalescent plasma have changed over time, and generally, the required antibody content has increased. In this context, the discordance between assays and changing clinical guidelines may have contributed to heterogeneity of individual trial findings. This meta-analysis found that treatment with high-titer convalescent plasma was associated with a 15% decrease in mortality rates, and treatment with low-titer convalescent plasma may not confer a clinically meaningful mortality benefit.

Critical reporting and sufficient analyses are crucial when it comes to investigating heterogeneity of meta-analyses in systematic reviews. Failure to fully reflect heterogeneity of results may lead to misinterpretations, incorrect assumptions, and incorrect and potentially harmful clinical recommendations. In this framework, this meta-analysis may provide new insights into the therapeutic use of convalescent plasma. Although this meta-analysis revealed that convalescent plasma was associated with a clinically meaningful overall mortality benefit among hospitalized patients with COVID-19, convalescent plasma is not a panacea because it is a non-standardized therapy that requires early use for efficacy.

COVID-19 convalescent plasma may be unlikely to confer a mortality benefit among patients treated with low antibody-titer convalescent plasma later in the COVID-19 disease course. The finding that COVID-19 convalescent plasma was associated with reduced mortality is consistent with the historical experience from the 1918 pandemic where convalescent serum therapy was associated with reduced mortality.^165^

## Limitations

This systematic review and meta-analysis has several limitations. First, the empirical, clinical studies were associated with biological diversity and heterogenous comparator groups. Key factors were continuously evolving during the pandemic, including the pathogen of interest (SARS-CoV-2), contemporary treatment strategies for the disease of interest (COVID-19), and antibody content of the treatment of interest (COVID-19 convalescent plasma). In this context, we believe the high level of concordance among study outcomes despite the biological heterogeneity between studies offer compelling evidence for the therapeutic value of convalescent plasma among COVID-19 patients overall. Second, we did not have access to patient-level data for the studies included in this article. Thus, our subgroup analyses that separated patients by a single baseline characteristic were simplistic. ^166^ Lack of patient-level data does not allow analyses using more complex statistical models that incorporate multiple characteristics^167^, and previous studies have pooled patient level data from clinical trials of COVID-19 convalescent plasma.^6,49^ Third, we limited our focus to a single outcome. Finally, we note that the preponderance of data in this analysis came from studies in the first years of the pandemic involving unvaccinated populations that were immunologically naïve to SARS-CoV-2 and today most individuals in the countries contributing the majority of studies are vaccinated and/or have prior experience with COVID-19, making the populations then and now immunologically different.

## Conclusion

This systematic review and meta-analysis found that convalescent plasma was associated with a 13% decrease in mortality rates in hospitalized patients with COVID-19 compared with a control cohort. Subgroup analyses revealed that patients treated with high-titer plasma and patients treated earlier in the course of the disease benefited more from COVID-19 convalescent plasma transfusion. Thus, reasonable concerns about the use of low antibody-titer convalescent plasma later in the course of the disease remain. These findings can offer experts a new starting point in forming their judgement of the therapeutic effectiveness of COVID-19 convalescent plasma and may help transform subjective and nebulous insider views to more transparent and reliable knowledge base. Finally, our findings support the deployment of convalescent plasma in future epidemic emergencies until better therapies are available with the caveat that convalescent plasma should be administered early in disease using units with the highest antibody content available.

## Data Availability

This manuscript represents a meta-analysis of published studies. In this context, this secondary research did not generate original data, which remain available at the cited references.

## ARTICLE INFORMATION

### Author Contributions

Drs Senefeld and Joyner had full access to all of the data in the study and take responsibility for the integrity of the data and the accuracy of the data analysis.

### Concept and design

Senefeld, Klassen, Carter, Paneth, Casadevall, Joyner

### Acquisition, analysis, or interpretation of data

Senefeld, Gorman, Johnson, Moir, Klassen, Paneth, Morkeberg, Casadevall, Joyner

### Drafting of the manuscript

Senefeld, Gorman, Casadevall, Joyner

### Critical revision of the manuscript for important intellectual content

Senefeld, Gorman, Johnson, Moir, Klassen, Carter, Paneth, Sullivan, Morkeberg, Wright, Fairweather, Bruno, Shoham, Bloch, Focosi, Henderson, Juskewitch, Pirofski, Grossman, Tobian, Franchini, Ganesh, Hurt, Kay, Parikh, Baker, Buchholtz, Buras, Clayburn, Dennis, Diaz Soto, Herasevich, Klompas, Kunze, Larson, Mills, Rea, Regimbal, Ripoll, Sexton, Shepherd, Stubbs, Theel, van Buskirk, van Helmond, Vogt, Whelan, Wiggins, Winters, Casadevall, Joyner

### Statistical analysis

Senefeld, Gorman, Casadevall, Joyner

### Obtained funding

Senefeld, Carter, Paneth, Wright, Fairweather, Bruno, Casadevall, Joyner

*Supervision:*

Senefeld, Carter, Paneth, Casadevall, Joyner

## REFERENCES

1. Shen C, Wang Z, Zhao F, et al. Treatment of 5 Critically Ill Patients With COVID-19 With Convalescent Plasma. JAMA. 2020;323(16):1582–1589.

2. Duan K, Liu B, Li C, et al. Effectiveness of convalescent plasma therapy in severe COVID-19 patients. Proc Natl Acad Sci U S A. 2020;117(17):9490–9496.

3. Ye M, Fu D, Ren Y, et al. Treatment with convalescent plasma for COVID-19 patients in Wuhan, China. J Med Virol. 2020;92(10):1890–1901.

4. Pommeret F, Colomba J, Bigenwald C, et al. Bamlanivimab + etesevimab therapy induces SARS-CoV-2 immune escape mutations and secondary clinical deterioration in COVID-19 patients with B-cell malignancies. Ann Oncol. 2021;32(11):1445–1447.

5. Jary A, Marot S, Faycal A, et al. Spike Gene Evolution and Immune Escape Mutations in Patients with Mild or Moderate Forms of COVID-19 and Treated with Monoclonal Antibodies Therapies. Viruses. 2022;14(2).

6. Troxel AB, Petkova E, Goldfeld K, et al. Association of Convalescent Plasma Treatment With Clinical Status in Patients Hospitalized With COVID-19: A Meta-analysis. JAMA Netw Open. 2022;5(1):e2147331.

7. Senefeld JW, Franchini M, Mengoli C, et al. COVID-19 convalescent plasma for the treatment of immunocompromised patients: a systematic review. JAMA Netw Open. 2023;6(1):e2250647.

8. Casadevall A, Dragotakes Q, Johnson PW, et al. Convalescent plasma use in the USA was inversely correlated with COVID-19 mortality. Elife. 2021;10.

9. Piechotta V, Iannizzi C, Chai KL, et al. Convalescent plasma or hyperimmune immunoglobulin for people with COVID-19: a living systematic review. Cochrane Database Syst Rev. 2021;5:CD013600.

10. Agarwal A, Mukherjee A, Kumar G, et al. Convalescent plasma in the management of moderate covid-19 in adults in India: open label phase II multicentre randomised controlled trial (PLACID Trial). BMJ. 2020;371:m3939.

11. Ali S, Uddin SM, Shalim E, et al. Hyperimmune anti-COVID-19 IVIG (C-IVIG) treatment in severe and critical COVID-19 patients: A phase I/II randomized control trial. EClinicalMedicine. 2021;36:100926.

12. AlQahtani M, Abdulrahman A, Almadani A, et al. Randomized controlled trial of convalescent plasma therapy against standard therapy in patients with severe COVID-19 disease. Sci Rep. 2021;11(1):9927.

13. Avendano-Sola C, Ramos-Martinez A, Munez-Rubio E, et al. A multicenter randomized open-label clinical trial for convalescent plasma in patients hospitalized with COVID-19 pneumonia. J Clin Invest. 2021;131(20).

14. Bajpai M, Maheshwari A, Dogra V, et al. Efficacy of convalescent plasma therapy in the patient with COVID-19: a randomised control trial (COPLA-II trial). BMJ Open. 2022;12(4):e055189.

15. Bajpai M, Maheshwari A, Kumar S, et al. Comparison of safety and efficacy of convalescent plasma with fresh frozen plasma in severe covid-19 patients. An Acad Bras Cienc. 2022;94(4):e20210202.

16. Baldeon ME, Maldonado A, Ochoa-Andrade M, et al. Effect of convalescent plasma as complementary treatment in patients with moderate COVID-19 infection. Transfus Med. 2022;32(2):153–161.

17. Bandopadhyay P, D’Rozario R, Lahiri A, et al. Nature and Dimensions of Systemic Hyperinflammation and its Attenuation by Convalescent Plasma in Severe COVID-19. J Infect Dis. 2021;224(4):565–574.

18. Bar KJ, Shaw PA, Choi GH, et al. A randomized controlled study of convalescent plasma for individuals hospitalized with COVID-19 pneumonia. J Clin Invest. 2021;131(24).

19. Bargay-Lleonart J, Sarubbo F, Arrizabalaga M, et al. Reinforcement of the Standard Therapy with Two Infusions of Convalescent Plasma for Patients with COVID-19: A Randomized Clinical Trial. J Clin Med. 2022;11(11).

20. Begin P, Callum J, Jamula E, et al. Convalescent plasma for hospitalized patients with COVID-19: an open-label, randomized controlled trial. Nat Med. 2021;27(11):2012–2024.

21. Bennett-Guerrero E, Romeiser JL, Talbot LR, et al. Severe Acute Respiratory Syndrome Coronavirus 2 Convalescent Plasma Versus Standard Plasma in Coronavirus Disease 2019 Infected Hospitalized Patients in New York: A Double-Blind Randomized Trial. Crit Care Med. 2021;49(7):1015–1025.

22. De Santis GC, Oliveira LC, Garibaldi PMM, et al. High-Dose Convalescent Plasma for Treatment of Severe COVID-19. Emerg Infect Dis. 2022;28(3):548–555.

23. Denkinger CM, Janssen M, Schäkel U, et al. Anti-SARS-CoV-2 antibody containing plasma improves outcome in patients with hematologic or solid cancer and severe COVID-19 via increased neutralizing antibody activity – a randomized clinical trial. medRxiv. 2022:2022.2010.2010.22280850.

24. Devos T, Van Thillo Q, Compernolle V, et al. Early high antibody titre convalescent plasma for hospitalised COVID-19 patients: DAWn-plasma. Eur Respir J. 2022;59(2).

25. Fernández-Sánchez V, Ventura-Enríquez Y, Cabello-Gutiérrez C, et al. Convalescent Plasma to Treat Covid-19: a Randomized Double Blind 2 Centers Trial. 2022.

26. Gharbharan A, Jordans CCE, GeurtsvanKessel C, et al. Effects of potent neutralizing antibodies from convalescent plasma in patients hospitalized for severe SARS-CoV-2 infection. Nat Commun. 2021;12(1):3189.

27. Gonzalez JLB, González Gámez M, Mendoza Enciso EA, et al. Efficacy and safety of convalescent plasma and intravenous immunoglobulin in critically ill COVID-19 patients. A controlled clinical trial. medRxiv. 2021:2021.2003.2028.21254507.

28. Holm K, Lundgren MN, Kjeldsen-Kragh J, et al. Convalescence plasma treatment of COVID-19: results from a prematurely terminated randomized controlled open-label study in Southern Sweden. BMC Res Notes. 2021;14(1):440.

29. Jalili E, Khazaei S, Mohammadi A, et al. Effect of Convalescent Plasma Therapy on Clinical Improvement of COVID-19 Patients: A Randomized Clinical Trial. Tanaffos. 2022;21(1):24–30.

30. Kirenga B, Byakika-Kibwika P, Muttamba W, et al. Efficacy of convalescent plasma for treatment of COVID-19 in Uganda. BMJ Open Respir Res. 2021;8(1).

31. Korper S, Weiss M, Zickler D, et al. Results of the CAPSID randomized trial for high-dose convalescent plasma in patients with severe COVID-19. J Clin Invest. 2021;131(20).

32. Lacombe K, Hueso T, Porcher R, et al. COVID-19 convalescent plasma to treat hospitalised COVID-19 patients with or without underlying immunodeficiency. medRxiv. 2022:2022.2008.2009.22278329.

33. Li L, Zhang W, Hu Y, et al. Effect of Convalescent Plasma Therapy on Time to Clinical Improvement in Patients With Severe and Life-threatening COVID-19: A Randomized Clinical Trial. JAMA. 2020;324(5):460–470.

34. Menichetti F, Popoli P, Puopolo M, et al. Effect of High-Titer Convalescent Plasma on Progression to Severe Respiratory Failure or Death in Hospitalized Patients With COVID-19 Pneumonia: A Randomized Clinical Trial. JAMA Netw Open. 2021;4(11):e2136246.

35. O’Donnell MR, Grinsztejn B, Cummings MJ, et al. A randomized double-blind controlled trial of convalescent plasma in adults with severe COVID-19. J Clin Invest. 2021;131(13).

36. Ortigoza MB, Yoon H, Goldfeld KS, et al. Efficacy and Safety of COVID-19 Convalescent Plasma in Hospitalized Patients: A Randomized Clinical Trial. JAMA Intern Med. 2022;182(2):115–126.

37. Pouladzadeh M, Safdarian M, Eshghi P, et al. A randomized clinical trial evaluating the immunomodulatory effect of convalescent plasma on COVID-19-related cytokine storm. Intern Emerg Med. 2021;16(8):2181–2191.

38. Rasheed AM, Fatak DF, Hashim HA, et al. The therapeutic potential of convalescent plasma therapy on treating critically-ill COVID-19 patients residing in respiratory care units in hospitals in Baghdad, Iraq. Infez Med. 2020;28(3):357–366.

39. Ray Y, Paul SR, Bandopadhyay P, et al. A phase 2 single center open label randomised control trial for convalescent plasma therapy in patients with severe COVID-19. Nat Commun. 2022;13(1):383.

40. Rojas M, Rodriguez Y, Hernandez JC, et al. Safety and efficacy of convalescent plasma for severe COVID-19: a randomized, single blinded, parallel, controlled clinical study. BMC Infect Dis. 2022;22(1):575.

41. Group RC. Convalescent plasma in patients admitted to hospital with COVID-19 (RECOVERY): a randomised controlled, open-label, platform trial. Lancet. 2021;397(10289):2049–2059.

42. Writing Committee for the R-CAPI, Estcourt LJ, Turgeon AF, et al. Effect of Convalescent Plasma on Organ Support-Free Days in Critically Ill Patients With COVID-19: A Randomized Clinical Trial. JAMA. 2021;326(17):1690–1702.

43. Sekine L, Arns B, Fabro BR, et al. Convalescent plasma for COVID-19 in hospitalised patients: an open-label, randomised clinical trial. Eur Respir J. 2022;59(2).

44. Self WH, Wheeler AP, Stewart TG, et al. Neutralizing COVID-19 Convalescent Plasma in Adults Hospitalized With COVID-19: A Blinded, Randomized, Placebo-Controlled Trial. Chest. 2022;162(5):982–994.

45. Simonovich VA, Burgos Pratx LD, Scibona P, et al. A Randomized Trial of Convalescent Plasma in Covid-19 Severe Pneumonia. N Engl J Med. 2021;384(7):619–629.

46. Song ATW, Rocha V, Mendrone-Junior A, et al. Treatment of severe COVID-19 patients with either low- or high-volume of convalescent plasma versus standard of care: A multicenter Bayesian randomized open-label clinical trial (COOP-COVID-19-MCTI). Lancet Reg Health Am. 2022;10:100216.

47. Thorlacius-Ussing L, Brooks PT, Nielsen H, et al. A randomized placebo-controlled trial of convalescent plasma for adults hospitalized with COVID-19 pneumonia. Sci Rep. 2022;12(1):16385.

48. van den Berg K, Glatt TN, Vermeulen M, et al. Convalescent plasma in the treatment of moderate to severe COVID-19 pneumonia: a randomized controlled trial (PROTECT-Patient Trial). Sci Rep. 2022;12(1):2552.

49. Park H, Tarpey T, Liu M, et al. Development and Validation of a Treatment Benefit Index to Identify Hospitalized Patients With COVID-19 Who May Benefit From Convalescent Plasma. JAMA Netw Open. 2022;5(1):e2147375.

50. Axfors C, Janiaud P, Schmitt AM, et al. Association between convalescent plasma treatment and mortality in COVID-19: a collaborative systematic review and meta-analysis of randomized clinical trials. BMC Infect Dis. 2021;21(1):1170.

51. Siemieniuk RA, Bartoszko JJ, Diaz Martinez JP, et al. Antibody and cellular therapies for treatment of covid-19: a living systematic review and network meta-analysis. BMJ. 2021;374:n2231.

52. Klassen SA, Senefeld JW, Johnson PW, et al. The Effect of Convalescent Plasma Therapy on Mortality Among Patients With COVID-19: Systematic Review and Meta-analysis. Mayo Clin Proc. 2021;96(5):1262–1275.

53. Millat-Martinez P, Gharbharan A, Alemany A, et al. Prospective individual patient data meta-analysis of two randomized trials on convalescent plasma for COVID-19 outpatients. Nat Commun. 2022;13(1):2583.

54. Joyner MJ, Carter RE, Senefeld JW, et al. Convalescent Plasma Antibody Levels and the Risk of Death from Covid-19. N Engl J Med. 2021;384(11):1015–1027.

55. Ioannidis JPA.Systematic reviews for basic scientists: a different beast. Physiol Rev. 2023;103(1):1–5.

56. Higgins JP, Thomas J, Chandler J, et al. Cochrane Handbook for Systematic Reviews of Interventions, Version 6.2. John Wiley & Sons; 2019.

57. Sterne JAC, Savovic J, Page MJ, et al. RoB 2: a revised tool for assessing risk of bias in randomised trials. BMJ. 2019;366:l4898.

58. Higgins JP, Altman DG, Gotzsche PC, et al. The Cochrane Collaboration’s tool for assessing risk of bias in randomised trials. BMJ. 2011;343:d5928.

59. Abolghasemi H, Eshghi P, Cheraghali AM, et al. Clinical efficacy of convalescent plasma for treatment of COVID-19 infections: Results of a multicenter clinical study. Transfus Apher Sci. 2020;59(5):102875.

60. Abuzakouk M, Saleh K, Algora M, et al. Convalescent Plasma Efficacy in Life-Threatening COVID-19 Patients Admitted to the ICU: A Retrospective Cohort Study. J Clin Med. 2021;10(10).

61. Acosta-Ampudia Y, Monsalve DM, Rojas M, et al. COVID-19 convalescent plasma composition and immunological effects in severe patients. J Autoimmun. 2021;118:102598.

62. Alamgir J, Abid MR, Garibaldi B, et al. LACK OF ASSOCIATION BETWEEN CONVALESCENT PLASMA ADMINISTRATION AND LENGTH OF HOSPITAL STAY: A HOSPITAL-DAY STRATIFIED MULTI-CENTER RETROSPECTIVE COHORT STUDY. medRxiv. 2021:2021.2005.2004.21256627.

63. Al Harthi S, Osali MA, Kindi NA, et al. Characteristics of the First 102 Severe COVID-19 Cases Treated With Convalescent Plasma or Tocilizumab or Both in Al-Nahdha Hospital, Oman. Health Serv Res Manag Epidemiol. 2021;8:2333392820986639.

64. Allahyari A, Seddigh-Shamsi M, Mahmoudi M, et al. Efficacy and safety of convalescent plasma therapy in severe COVID-19 patients with acute respiratory distress syndrome. Int Immunopharmacol. 2021;93:107239.

65. Alsharidah S, Ayed M, Ameen RM, et al. COVID-19 convalescent plasma treatment of moderate and severe cases of SARS-CoV-2 infection: A multicenter interventional study. Int J Infect Dis. 2021;103:439–446.

66. AlShehry N, Zaidi SZA, AlAskar A, et al. Safety and Efficacy of Convalescent Plasma for Severe COVID-19: Interim Report of a Multicenter Phase II Study from Saudi Arabia. Saudi J Med Med Sci. 2021;9(1):16–23.

67. Altuntas F, Ata N, Yigenoglu TN, et al. Convalescent plasma therapy in patients with COVID-19. Transfus Apher Sci. 2021;60(1):102955.

68. Arnold Egloff SA, Junglen A, Restivo JS, et al. Convalescent plasma associates with reduced mortality and improved clinical trajectory in patients hospitalized with COVID-19. J Clin Invest. 2021;131(20).

69. Ates I, Erden A, Guven SC, et al. Should timing be considered before abandoning convalescent plasma in covid-19? Results from the Turkish experience. Transfus Apher Sci. 2021;60(6):103238.

70. Biernat MM, Kolasinska A, Kwiatkowski J, et al. Early Administration of Convalescent Plasma Improves Survival in Patients with Hematological Malignancies and COVID-19. Viruses. 2021;13(3).

71. Bihariesingh R, Bansie R, Froberg J, et al. Mortality reduction in ICU-admitted COVID-19 patients in Suriname after treatment with convalescent plasma acquired via gravity filtration. Anesthesia & Clinical Research. 2021;2(2):2–12.

72. Briggs N, Gormally MV, Li F, et al. Early but not late convalescent plasma is associated with better survival in moderate-to-severe COVID-19. PLoS One. 2021;16(7):e0254453.

73. Budhiraja S, Dewan A, Aggarwal R, et al. Effectiveness of convalescent plasma in Indian patients with COVID-19. Blood Cells Mol Dis. 2021;88:102548.

74. Cacilhas P, Caberlon E, Angoleri L, Fassina K, Ribeiro RN, Pinto LC. Convalescent plasma therapy in COVID-19 patients: a non-randomized case-control study with concurrent control. Braz J Med Biol Res. 2022;55:e12235.

75. Chauhan L, Pattee J, Ford J, et al. A Multicenter, Prospective, Observational, Cohort-Controlled Study of Clinical Outcomes Following Coronavirus Disease 2019 (COVID-19) Convalescent Plasma Therapy in Hospitalized Patients With COVID-19. Clin Infect Dis. 2022;75(1):e466–e472.

76. Cho K, Keithly SC, Kurgansky KE, et al. Early Convalescent Plasma Therapy and Mortality Among US Veterans Hospitalized With Nonsevere COVID-19: An Observational Analysis Emulating a Target Trial. J Infect Dis. 2021;224(6):967–975.

77. Cristelli MP, Langhi Junior DM, Viana LA, et al. Efficacy of Convalescent Plasma to Treat Mild to Moderate COVID-19 in Kidney Transplant Patients: A Propensity Score Matching Analysis. Transplantation. 2022;106(1):e92–e94.

78. Dai W, Wu J, Li T, et al. Clinical outcomes for COVID-19 patients with diabetes mellitus treated with convalescent plasma transfusion in Wuhan, China. J Med Virol. 2021;93(4):2321–2331.

79. Donato ML, Park S, Baker M, et al. Clinical and laboratory evaluation of patients with SARS-CoV-2 pneumonia treated with high-titer convalescent plasma. JCI Insight. 2021;6(6).

80. Eren E, Ulu-Kilic A, Korkmaz S, et al. Retrospective analysis on efficacy of convalescent plasma in acute respiratory distress syndrome due to COVID-19. Sao Paulo Med J. 2022;140(1):12–16.

81. Garcia-Munoz R, Farfan-Quiroga G, Ruiz-de-Lobera N, et al. Serology-based therapeutic strategy in SARS-CoV-2-infected patients. Int Immunopharmacol. 2021;101(Pt B):108214.

82. Hatzl S, Posch F, Sareban N, et al. Convalescent plasma therapy and mortality in COVID-19 patients admitted to the ICU: a prospective observational study. Ann Intensive Care. 2021;11(1):73.

83. Hegerova L, Gooley TA, Sweerus KA, et al. Use of convalescent plasma in hospitalized patients with COVID-19: case series. Blood. 2020;136(6):759–762.

84. Hoepler WP, Weidner L, Traugott MT, et al. Adjunctive treatment with high-titre convalescent plasma in severely and critically ill COVID-19 patients - a safe but futile intervention. A comparative cohort study. Infect Dis (Lond). 2021;53(11):820–829.

85. Huang L, Zhang C, Zhou X, et al. Convalescent plasma is of limited clinical benefit in critically ill patients with coronavirus disease-2019: a cohort study. J Transl Med. 2021;19(1):365.

86. Jiang W, Li W, Xiong L, et al. Clinical efficacy of convalescent plasma therapy on treating COVID-19 patients: Evidence from matched study and a meta-analysis. Clin Transl Med. 2020;10(8):e259.

87. Khamis F, Al Arimi Z, Al Naamani H, et al. Convalescent Plasma Therapy in Critically Ill COVID-19 Patients: An Open Label Trial. Oman Med J. 2021;36(5):e296.

88. Klapholz M, Pentakota SR, Zertuche JP, et al. Matched Cohort Study of Convalescent COVID-19 Plasma Treatment in Severely or Life Threateningly Ill COVID-19 Patients. Open Forum Infect Dis. 2021;8(2):ofab001.

89. Klein MN, Wang EW, Zimand P, et al. Kinetics of SARS-CoV-2 antibody responses pre-COVID-19 and post-COVID-19 convalescent plasma transfusion in patients with severe respiratory failure: an observational case-control study. J Clin Pathol. 2022;75(8):564–571.

90. Koirala J, Gyanwali P, Gerzoff RB, et al. Experience of Treating COVID-19 With Remdesivir and Convalescent Plasma in a Resource-Limited Setting: A Prospective, Observational Study. Open Forum Infect Dis. 2021;8(8):ofab391.

91. Kuno T, Takahashi M, Egorova NN. The Association Between Convalescent Plasma Treatment and Survival of Patients with COVID-19. J Gen Intern Med. 2021;36(8):2528–2531.

92. Kurnianda J, Hardianti MS, Triyono T, et al. Efficacy and safety of convalescent plasma therapy in patients with moderate-to-severe COVID-19: A non-randomized comparative study with historical control in a referral hospital in Indonesia. J Infect Public Health. 2022;15(1):100–108.

93. Kurtz P, Righy C, Gadelha M, et al. Effect of Convalescent Plasma in Critically Ill Patients With COVID-19: An Observational Study. Front Med (Lausanne). 2021;8:630982.

94. Lanza F, Monaco F, Ciceri F, et al. Lack of efficacy of convalescent plasma in COVID-19 patients with concomitant hematological malignancies: An Italian retrospective study. Hematol Oncol. 2022;40(5):857–863.

95. Liao M, Liao X, Yuan J, et al. The concentrated antibody from convalescent plasma balanced the dysfunctional immune responses in patients with critical COVID-19. Clin Transl Med. 2021;11(11):e571.

96. Liu STH, Lin HM, Baine I, et al. Convalescent plasma treatment of severe COVID-19: a propensity score-matched control study. Nat Med. 2020;26(11):1708–1713.

97. Mahapatra S, Rattan R, Mohanty CBK. Convalescent Plasma Therapy in the management of COVID-19 patients-The newer dimensions. Transfus Clin Biol. 2021;28(3):246–253.

98. Mendoza RP, Fyke W, Daniel D, et al. Administration of high titer convalescent anti-SARS-CoV-2 plasma: From donor selection to monitoring recipient outcomes. Hum Immunol. 2021;82(4):255–263.

99. Mesina FZ, Mangahas CG, Gatchalian EM, Ariola-Ramos MS, Torres RP. Use of Convalescent Plasma Therapy among Hospitalized Coronavirus Disease 2019 (COVID-19) Patients: A Single-Center Experience. medRxiv. 2021:2021.2002.2016.21251824.

100. Mesina F, Julian J, Relos J, et al. Use of Convalescent Plasma Therapy with Best Available Treatment (BAT) among Hospitalized COVID-19 Patients: A Multi-Center Study. medRxiv. 2022:2022.2002.2023.22271424.

101. Moniuszko-Malinowska A, Czupryna P, Zarebska-Michaluk D, et al. Convalescent Plasma Transfusion for the Treatment of COVID-19-Experience from Poland: A Multicenter Study. J Clin Med. 2020;10(1).

102. Novacescu AN, Duma G, Buzzi B, et al. Therapeutic plasma exchange followed by convalescent plasma transfusion in severe and critically ill COVID-19 patients: A single centre non-randomized controlled trial. Exp Ther Med. 2022;23(1):76.

103. Omrani AS, Zaqout A, Baiou A, et al. Convalescent plasma for the treatment of patients with severe coronavirus disease 2019: A preliminary report. J Med Virol. 2021;93(3):1678–1686.

104. Pan C, Chen H, Xie J, et al. The Efficiency of Convalescent Plasma Therapy in the Management of Critically Ill Patients Infected With COVID-19: A Matched Cohort Study. Front Med (Lausanne). 2022;9:822821.

105. Pappa V, Bouchla A, Terpos E, et al. A Phase II Study on the Use of Convalescent Plasma for the Treatment of Severe COVID-19-A Propensity Score-Matched Control Analysis. Microorganisms. 2021;9(4).

106. Perotti C, Baldanti F, Bruno R, et al. Mortality reduction in 46 severe Covid-19 patients treated with hyperimmune plasma. A proof of concept single arm multicenter trial. Haematologica. 2020;105(12):2834–2840.

107. Rogers R, Shehadeh F, Mylona EK, et al. Convalescent Plasma for Patients With Severe Coronavirus Disease 2019 (COVID-19): A Matched Cohort Study. Clin Infect Dis. 2021;73(1):e208–e214.

108. Rollas K, Emgin O, Caliskan T, et al. Convalescent plasma for COVID-19 in the intensive care unit. Anaesthesiol Intensive Ther. 2021;53(5):398–402.

109. Romon I, Dominguez-Garcia JJ, Arroyo JL, et al. Convalescent plasma treatment for patients of 80 years and older with COVID-19 pneumonia. BMC Geriatr. 2021;21(1):566.

110. Sajmi S, Goutham K, Arumugam V, et al. Efficacy and safety of convalescent plasma therapy in SARS-CoV2 patients on hemodialysis. Hemodial Int. 2021;25(4):515–522.

111. Salazar E, Christensen PA, Graviss EA, et al. Significantly Decreased Mortality in a Large Cohort of Coronavirus Disease 2019 (COVID-19) Patients Transfused Early with Convalescent Plasma Containing High-Titer Anti-Severe Acute Respiratory Syndrome Coronavirus 2 (SARS-CoV-2) Spike Protein IgG. Am J Pathol. 2021;191(1):90–107.

112. Salazar MR, Gonzalez SE, Regairaz L, et al. Risk factors for COVID-19 mortality: The effect of convalescent plasma administration. PLoS One. 2021;16(4):e0250386.

113. Sammartino D, Jafri F, Cook B, et al. Predictors for inpatient mortality during the first wave of the SARS-CoV-2 pandemic: A retrospective analysis. PLoS One. 2021;16(5):e0251262.

114. Sanz C, Nomdedeu M, Pereira A, et al. Efficacy of early transfusion of convalescent plasma with high-titer SARS-CoV-2 neutralizing antibodies in hospitalized patients with COVID-19. Transfusion. 2022;62(5):974–981.

115. Semedi BP, Ramadhania NN, Tambunan BA, Bintoro SUY, Soedarsono S, Prakoeswa CRS. Prolonged ICU Stay in Severe and Critically-Ill COVID-19 Patients Who Received Convalescent Plasma Therapy. Crit Care Res Pract. 2022;2022:1594342.

116. Shenoy AG, Hettinger AZ, Fernandez SJ, Blumenthal J, Baez V. Early mortality benefit with COVID-19 convalescent plasma: a matched control study. Br J Haematol. 2021;192(4):706–713.

117. Sostin OV, Rajapakse P, Cruser B, Wakefield D, Cruser D, Petrini J. A matched cohort study of convalescent plasma therapy for COVID-19. J Clin Apher. 2021;36(4):523–532.

118. Sturek JM, Thomas TA, Gorham JD, et al. Convalescent Plasma for Preventing Critical Illness in COVID-19: a Phase 2 Trial and Immune Profile. Microbiol Spectr. 2022;10(1):e0256021.

119. Tang J, Grubbs G, Lee Y, Golding H, Khurana S. Impact of Convalescent Plasma Therapy on Severe Acute Respiratory Syndrome Coronavirus 2 (SARS-CoV-2) Antibody Profile in Coronavirus Disease 2019 (COVID-19) Patients. Clin Infect Dis. 2022;74(2):327–334.

120. Thompson MA, Henderson JP, Shah PK, et al. Association of Convalescent Plasma Therapy With Survival in Patients With Hematologic Cancers and COVID-19. JAMA Oncol. 2021;7(8):1167–1175.

121. Tworek A, Jaron K, Uszynska-Kaluza B, et al. Convalescent plasma treatment is associated with lower mortality and better outcomes in high-risk COVID-19 patients - propensity-score matched case-control study. Int J Infect Dis. 2021;105:209–215.

122. Weisser M, Khanna N, Hedstueck A, et al. Characterization of pathogen-inactivated COVID-19 convalescent plasma and responses in transfused patients. Transfusion. 2022;62(10):1997–2011.

123. Xia X, Li K, Wu L, et al. Improved clinical symptoms and mortality among patients with severe or critical COVID-19 after convalescent plasma transfusion. Blood. 2020;136(6):755–759.

124. Xiao K, Lin Y, Fan Z, et al. Effect of transfusion convalescent recovery plasma in patients with coronavirus disease 2019. Zhong Nan Da Xue Xue Bao Yi Xue Ban. 2020;45(5):565–570.

125. Yoon HA, Bartash R, Gendlina I, et al. Treatment of severe COVID-19 with convalescent plasma in Bronx, NYC. JCI Insight. 2021;6(4).

126. Zeng QL, Yu ZJ, Gou JJ, et al. Effect of Convalescent Plasma Therapy on Viral Shedding and Survival in Patients With Coronavirus Disease 2019. J Infect Dis. 2020;222(1):38–43.

127. Zhou CK, Bennett MM, Villa CH, et al. Multi-center matched cohort study of convalescent plasma for hospitalized patients with COVID-19. PLoS One. 2022;17(8):e0273223.

128. Alemany A, Millat-Martinez P, Corbacho-Monne M, et al. High-titre methylene blue-treated convalescent plasma as an early treatment for outpatients with COVID-19: a randomised, placebo-controlled trial. Lancet Respir Med. 2022;10(3):278–288.

129. Gharbharan A, Jordans C, Zwaginga L, et al. Outpatient convalescent plasma therapy for high-risk patients with early COVID-19: a randomized placebo-controlled trial. Clin Microbiol Infect. 2022.

130. Shoham S, Bloch EM, Casadevall A, et al. Transfusing convalescent plasma as post-exposure prophylaxis against SARS-CoV-2 infection: a double-blinded, phase 2 randomized, controlled trial. Clin Infect Dis. 2022.

131. Sullivan DJ, Gebo KA, Shoham S, et al. Early Outpatient Treatment for Covid-19 with Convalescent Plasma. N Engl J Med. 2022;386(18):1700–1711.

132. Korley FK, Durkalski-Mauldin V, Yeatts SD, et al. Early Convalescent Plasma for High-Risk Outpatients with Covid-19. N Engl J Med. 2021;385(21):1951–1960.

133. Libster R, Perez Marc G, Wappner D, et al. Early High-Titer Plasma Therapy to Prevent Severe Covid-19 in Older Adults. N Engl J Med. 2021;384(7):610–618.

134. Bartelt LA, Markmann AJ, Nelson B, et al. Outcomes of Convalescent Plasma with Defined High versus Lower Neutralizing Antibody Titers against SARS-CoV-2 among Hospitalized Patients: CoronaVirus Inactivating Plasma (CoVIP) Study. mBio. 2022;13(5):e0175122.

135. Belov A, Huang Y, Villa CH, et al. Early administration of COVID-19 convalescent plasma with high titer antibody content by live viral neutralization assay is associated with modest clinical efficacy. Am J Hematol. 2022;97(6):770–779.

136. Cain WV, Sill AM, Solipuram V, Weiss JJ, Miller CB, Jelsma PF. Efficacy of COVID-19 Convalescent Plasma Based on Antibody Concentration. Adv Hematol. 2022;2022:7992927.

137. Fazeli A, Sharifi S, Behdad F, et al. Early high-titer convalescent plasma therapy in patients with moderate and severe COVID-19. Transfus Apher Sci. 2022;61(2):103321.

138. Franchini M, Glingani C, Donno G, et al. Convalescent Plasma for Hospitalized COVID-19 Patients: A Single-Center Experience. Life (Basel). 2022;12(3).

139. Gachoud D, Pillonel T, Tsilimidos G, et al. Antibody response and intra-host viral evolution after plasma therapy in COVID-19 patients pre-exposed or not to B-cell-depleting agents. Br J Haematol. 2022;199(4):549–559.

140. Gonzalez SE, Regairaz L, Salazar MR, et al. Timing of convalescent plasma administration and 28-day mortality in COVID-19 pneumonia. J Investig Med. 2022;70(5):1258–1264.

141. Hemsinlioglu C, Pelit N, Yalcin K, et al. THE EFFECTIVENESS OF ACB-IP 1.0 UNIVERSAL PATHOGEN FREE CONCENTRATED COCKTAIL CONVALESCENT PLASMA IN COVID-19 INFECTION. medRxiv. 2021:2021.2003.2005.21251413.

142. Khan TNS, Mukry SN, Masood S, et al. Usefulness of convalescent plasma transfusion for the treatment of severely ill COVID-19 patients in Pakistan. BMC Infect Dis. 2021;21(1):1014.

143. Leon J, Merrill AE, Rogers K, et al. SARS-CoV-2 antibody changes in patients receiving COVID-19 convalescent plasma from normal and vaccinated donors. Transfus Apher Sci. 2022;61(2):103326.

144. Maor Y, Cohen D, Paran N, et al. Compassionate use of convalescent plasma for treatment of moderate and severe pneumonia in COVID-19 patients and association with IgG antibody levels in donated plasma. EClinicalMedicine. 2020;26:100525.

145. Akay Cizmecioglu H, Goktepe MH, Demircioglu S, et al. Efficacy of convalescent plasma therapy in severe COVID-19 patients. Transfus Apher Sci. 2021;60(4):103158.

146. Asem N, Massoud HH, Serag I, et al. Clinical Efficacy of Early Administration of Convalescent Plasma among COVID-19 Cases in Egypt. Open Access Macedonian Journal of Medical Sciences. 2022;10(B):1698–1705.

147. Balcells ME, Rojas L, Le Corre N, et al. Early versus deferred anti-SARS-CoV-2 convalescent plasma in patients admitted for COVID-19: A randomized phase II clinical trial. PLoS Med. 2021;18(3):e1003415.

148. De Silvestro G, Marson P, La Raja M, et al. Outcome of SARS CoV-2 inpatients treated with convalescent plasma: One-year of data from the Veneto region (Italy) Registry. Eur J Intern Med. 2022;97:42–49.

149. Fodor E, Muller V, Ivanyi Z, et al. Early Transfusion of Convalescent Plasma Improves the Clinical Outcome in Severe SARS-CoV2 Infection. Infect Dis Ther. 2022;11(1):293–304.

150. Gazitúa R, Briones JL, Selman C, et al. Convalescent Plasma in COVID-19. Mortality-Safety First Results of the Prospective Multicenter FALP 001-2020 Trial. medRxiv. 2020:2020.2011.2030.20218560.

151. Greenbaum U, Klein K, Martinez F, et al. High Levels of Common Cold Coronavirus Antibodies in Convalescent Plasma Are Associated With Improved Survival in COVID-19 Patients. Front Immunol. 2021;12:675679.

152. Hueso T, Godron AS, Lanoy E, et al. Convalescent plasma improves overall survival in patients with B-cell lymphoid malignancy and COVID-19: a longitudinal cohort and propensity score analysis. Leukemia. 2022;36(4):1025–1034.

153. Ibrahim D, Dulipsingh L, Zapatka L, et al. Factors Associated with Good Patient Outcomes Following Convalescent Plasma in COVID-19: A Prospective Phase II Clinical Trial. Infect Dis Ther. 2020;9(4):913–926.

154. Janaka SK, Hartman W, Mou H, et al. Donor Anti-Spike Immunity is Related to Recipient Recovery and Can Predict the Efficacy of Convalescent Plasma Units. medRxiv. 2021:2021.2002.2025.21252463.

155. Jeyaraman P, Agrawal N, Bhargava R, et al. Convalescent plasma therapy for severe Covid-19 in patients with hematological malignancies. Transfus Apher Sci. 2021;60(3):103075.

156. Kocayigit H, Demir G, Karacan A, et al. Effects on mortality of early vs late administration of convalescent plasma in the treatment of Covid-19. Transfus Apher Sci. 2021;60(4):103148.

157. Lattanzio N, Acosta-Diaz C, Villasmil RJ, et al. Effectiveness of COVID-19 Convalescent Plasma Infusion Within 48 Hours of Hospitalization With SARS-CoV-2 Infection. Cureus. 2021;13(7):e16746.

158. Senefeld JW, Johnson PW, Kunze KL, et al. Access to and safety of COVID-19 convalescent plasma in the United States Expanded Access Program: A national registry study. PLoS Med. 2021;18(12):e1003872.

159. Levine AC, Fukuta Y, Huaman MA, et al. COVID-19 Convalescent Plasma Outpatient Therapy to Prevent Outpatient Hospitalization: A Meta-analysis of Individual Participant Data From Five Randomized Trials. medRxiv. 2022:2022.2012.2016.22283585.

160. Ma T, Wiggins CC, Kornatowski BM, et al. The Role of Disease Severity and Demographics in the Clinical Course of COVID-19 Patients Treated With Convalescent Plasma. Front Med (Lausanne). 2021;8:707895.

161. Ripoll JG, Gorman EK, Juskewitch JE, et al. Vaccine-boosted convalescent plasma therapy for patients with immunosuppression and COVID-19. Blood Adv. 2022;6(23):5951–5955.

162. Senefeld JW, Franchini M, Mengoli C, et al. COVID-19 Convalescent Plasma for the Treatment of Immunocompromised Patients: A Systematic Review and Meta-analysis. JAMA Netw Open. 2023;6(‘):e2250647.

163. Kunze KL, Johnson PW, van Helmond N, et al. Mortality in individuals treated with COVID-19 convalescent plasma varies with the geographic provenance of donors. Nat Commun. 2021;12(1):4864.

164. Larkey NE, Ewaisha R, Lasho MA, et al. Limited Correlation between SARS-CoV-2 Serologic Assays for Identification of High-Titer COVID-19 Convalescent Plasma Using FDA Thresholds. Microbiol Spectr. 2022;10(4):e0115422.

165. Luke TC, Kilbane EM, Jackson JL, Hoffman SL. Meta-analysis: convalescent blood products for Spanish influenza pneumonia: a future H5N1 treatment? Ann Intern Med. 2006;145(8):599–609.

166. Kent DM, Steyerberg E, van Klaveren D. Personalized evidence based medicine: predictive approaches to heterogeneous treatment effects. BMJ. 2018;363:k4245.

167. Kent DM, Paulus JK, van Klaveren D, et al. The Predictive Approaches to Treatment effect Heterogeneity (PATH) Statement. Ann Intern Med. 2020;172(1):35–45.

168. Focosi D, Casadevall A. Convalescent plasma in outpatients with COVID-19. Lancet Respir Med. 2022;10(3):226–228.

