## Supplemental Material for "Mortality rates among hospitalized patients with COVID-19 treated with convalescent plasma A Systematic review and meta-analysis"

© 2022 Senefeld JW et al.

### eResults

#### Case series and reports

In the United States and many other countries, the regulatory framework during the COVID-19 pandemic enabled broad access to COVID-19 convalescent plasma. In this context, a large number of single-arm studies evaluated the risk of death from COVID-19 after transfusion with convalescent plasma, including over 300 case series or case reports.[1-335] The case series and case reports treated 135,949 participants with COVID-19 convalescent plasma and mortality was observed in 33,771 participants (~25% mortality rate). Mortality results associated with case series and reports are provided in eTable6.

 © 2022 Senefeld JW et al.

| Source | Deaths/patients randomised (%) Convalescent plasma group | Usual care group | Ratio of death rates, OR (95% CI) | Weight |
|---|---|---|---|---|
| Al Harthi et al,[63] 2021 | 3/41 (7%) | 0/61 (0%) | 12.66 (1.23-130.22) | 0.54% |
| Liao et al,[95] 2021 | 3/7 (43%) | 0/6 (0%) | 9.29 (0.77-111.62) | 0.49% |
| Koirala et al,[90] 2021 | 88/173 (51%) | 146/910 (16%) | 7.80 (5.26-11.57) | 2.01% |
| Garcia-Muñoz et al,[81] 2021 | 1/26 (4%) | 0/20 (0%) | 5.87 (0.11-305.80) | 0.22% |
| Arnold Egloff et al,[68] 2021 | 505/4337 (12%) | 478/10687 (5%) | 3.23 (2.80-3.73) | 2.16% |
| Moniuszko-Malinowska et al,[101] 2020 | 10/78 (13%) | 43/715 (6%) | 2.97 (1.17-7.58) | 1.46% |
| Dai et al,[78] 2021 | 3/39 (8%) | 12/328 (4%) | 2.79 (0.52-14.89) | 0.85% |
| Huang et al,[85] 2021 | 3/14 (21%) | 1/10 (10%) | 2.20 (0.26-18.55) | 0.61% |
| Acosta-Ampudia et al,[61] 2021 | 2/9 (22%) | 1/9 (11%) | 2.13 (0.19-23.69) | 0.51% |
| Abuzakouk et al,[60] 2021 | 10/32 (31%) | 15/78 (19%) | 1.97 (0.74-5.24) | 1.42% |
| Cho et al,[76] 2021 | 40/402 (10%) | 343/4654 (7%) | 1.45 (0.98-2.12) | 2.02% |
| Mesina et al,[100] 2022 | 18/65 (28%) | 14/65 (22%) | 1.39 (0.63-3.08) | 1.61% |
| Zhou et al,[127] 2022 | 179/1245 (14%) | 143/1245 (12%) | 1.29 (1.02-1.63) | 2.12% |
| Chauhan et al,[75] 2021 | 27/188 (14%) | 23/188 (12%) | 1.20 (0.66-2.18) | 1.82% |
| Klein et al,[89] 2021 | 9/34 (26%) | 8/34 (24%) | 1.17 (0.39-3.47) | 1.31% |
| Ateş et al,[69] 2021 | 7/113 (6%) | 6/110 (5%) | 1.14 (0.37-3.50) | 1.28% |
| Klapholz et al,[88] 2021 | 10/47 (21%) | 9/47 (19%) | 1.14 (0.42-3.10) | 1.39% |
| Alamgir et al,[62] 2021 | 37/197 (19%) | 35/197 (18%) | 1.07 (0.64-1.78) | 1.91% |
| Tang et al,[119] 2022 | 4/8 (50%) | 4/8 (50%) | 1.00 (0.15-6.67) | 0.72% |
| Kuno et al,[91] 2021 | 241/960 (25%) | 250/960 (26%) | 0.95 (0.78-1.17) | 2.14% |
| Mesina et al,[99] 2021 | 19/75 (25%) | 20/75 (27%) | 0.93 (0.45-1.93) | 1.68% |
| Sajmi et al,[110] 2021 | 16/37 (43%) | 14/31 (45%) | 0.93 (0.36-2.41) | 1.44% |
| Shenoy et al,[116] 2021 | 67/263 (25%) | 71/263 (27%) | 0.93 (0.63-1.36) | 2.02% |
| Cristelli et al,[77] 2022 | 13/58 (22%) | 28/116 (24%) | 0.91 (0.43-1.91) | 1.67% |
| Mendoza et al,[98] 2021 | 14/28 (50%) | 9/17 (53%) | 0.89 (0.27-2.94) | 1.21% |
| Altuntas et al,[67] 2021 | 219/888 (25%) | 246/888 (28%) | 0.86 (0.69-1.06) | 2.13% |
| Mahapatra et al,[97] 2021 | 527/1189 (44%) | 608/1243 (49%) | 0.83 (0.71-0.98) | 2.16% |
| Lanza et al,[94] 2022 | 19/79 (24%) | 46/158 (29%) | 0.78 (0.42-1.42) | 1.81% |
| Sammartino et al,[113] 2021 | 32/105 (30%) | 38/105 (36%) | 0.77 (0.44-1.37) | 1.84% |
| Sostin et al,[117] 2021 | 7/35 (20%) | 15/61 (25%) | 0.77 (0.29-2.07) | 1.41% |
| Rollas et al,[108] 2021 | 53/89 (60%) | 65/99 (66%) | 0.77 (0.43-1.39) | 1.83% |
| Rogers et al,[107] 2021 | 8/64 (13%) | 28/177 (16%) | 0.77 (0.35-1.72) | 1.61% |
| Kurtz et al,[93] 2021 | 20/41 (49%) | 40/72 (56%) | 0.76 (0.36-1.64) | 1.64% |
| Kurnianda et al,[92] 2022 | 5/15 (33%) | 6/15 (40%) | 0.76 (0.18-3.26) | 0.99% |
| Yoon et al,[125] 2021 | 23/73 (32%) | 28/73 (38%) | 0.74 (0.38-1.46) | 1.74% |
| Briggs et al,[72] 2021 | 52/132 (39%) | 64/132 (48%) | 0.69 (0.43-1.13) | 1.93% |
| Budhiraja et al,[73] 2021 | 85/333 (26%) | 120/361 (33%) | 0.69 (0.50-0.96) | 2.06% |
| Semedi et al,[115] 2022 | 23/48 (48%) | 76/131 (58%) | 0.67 (0.34-1.29) | 1.75% |
| Sanz et al,[114] 2022 | 20/260 (8%) | 176/1343 (13%) | 0.60 (0.40-0.91) | 2.00% |
| Xia et al,[123] 2020 | 3/138 (2%) | 59/1430 (4%) | 0.60 (0.24-1.47) | 1.50% |
| Hoepler et al,[84] 2021 | 13/55 (24%) | 49/139 (35%) | 0.59 (0.30-1.15) | 1.75% |
| AlShehry et al,[66] 2021 | 10/40 (25%) | 46/124 (37%) | 0.59 (0.28-1.24) | 1.66% |
| Salazar MR et al,[112] 2021 | 221/868 (25%) | 1010/2661 (38%) | 0.58 (0.49-0.68) | 2.16% |
| Khamis et al,[87] 2021 | 14/73 (19%) | 6/21 (29%) | 0.57 (0.18-1.87) | 1.22% |
| Eren et al,[80] 2022 | 1/11 (9%) | 3/19 (16%) | 0.57 (0.07-4.89) | 0.61% |
| Pan et al,[104] 2022 | 12/26 (46%) | 47/78 (60%) | 0.57 (0.23-1.38) | 1.51% |
| Sturek et al,[118] 2022 | 2/29 (7%) | 6/49 (12%) | 0.56 (0.13-2.54) | 0.96% |
| Hatzl et al,[82] 2021 | 15/48 (31%) | 33/72 (46%) | 0.55 (0.26-1.15) | 1.67% |
| Donato et al,[79] 2021 | 11/51 (22%) | 985/2761 (36%) | 0.54 (0.30-0.96) | 1.84% |
| Abolghasemi et al,[59] 2020 | 17/115 (15%) | 18/74 (24%) | 0.53 (0.25-1.13) | 1.66% |
| Thompson et al,[120] 2021 | 19/143 (13%) | 204/823 (25%) | 0.52 (0.34-0.80) | 1.99% |
| Jiang et al,[86] 2020 | 8/163 (5%) | 15/163 (9%) | 0.52 (0.22-1.21) | 1.56% |
| Liu et al,[96] 2020 | 5/39 (13%) | 38/156 (24%) | 0.51 (0.22-1.19) | 1.56% |
| Salazar E et al,[111] 2021 | 20/321 (6%) | 73/584 (13%) | 0.51 (0.32-0.79) | 1.96% |
| Romon et al,[109] 2021 | 6/41 (15%) | 22/82 (27%) | 0.50 (0.21-1.22) | 1.51% |
| Bihariesingh-Sanchit et al,[71] 2021 | 5/28 (18%) | 18/50 (36%) | 0.42 (0.15-1.16) | 1.39% |
| Novacescu et al,[102] 2022 | 10/19 (53%) | 14/19 (74%) | 0.41 (0.11-1.52) | 1.12% |
| Alsharidah et al,[65] 2021 | 24/135 (18%) | 90/233 (39%) | 0.38 (0.24-0.60) | 1.96% |
| Allahyari et al,[64] 2021 | 7/32 (22%) | 14/32 (44%) | 0.38 (0.13-1.06) | 1.36% |
| Weisser et al,[122] 2022 | 1/15 (7%) | 6/30 (21%) | 0.37 (0.07-2.01) | 0.83% |
| Zeng et al,[126] 2020 | 5/6 (83%) | 14/15 (93%) | 0.33 (0.01-7.71) | 0.33% |
| Tworek et al,[121] 2021 | 14/102 (14%) | 35/102 (34%) | 0.33 (0.17-0.62) | 1.77% |
| Hegerova et al,[83] 2020 | 2/20 (10%) | 6/20 (30%) | 0.30 (0.06-1.37) | 0.94% |
| Pappa et al,[105] 2021 | 2/59 (3%) | 8/59 (15%) | 0.27 (0.08-0.99) | 1.13% |
| Biernat et al,[70] 2021 | 3/23 (13%) | 9/22 (41%) | 0.25 (0.07-0.92) | 1.11% |
| Omrani et al,[103] 2021 | 1/40 (3%) | 5/40 (13%) | 0.24 (0.05-1.26) | 0.86% |
| Perotti et al,[106] 2020 | 3/46 (7%) | 7/23 (30%) | 0.15 (0.04-0.61) | 1.03% |
| Cacilhas et al,[74] 2022 | 1/12 (8%) | 14/24 (61%) | 0.14 (0.03-0.54) | 1.05% |
| Duan et al,[2] 2020 | 0/10 (0%) | 3/10 (30%) | 0.11 (0.01-1.17) | 0.52% |
| Total | 2877/14541 (19.8%) | 6086/35619 (17.1%) | 0.77 (0.64-0.94) | |

Heterogeneity: $\tau^2 = 0.459$; $\chi^2 = 576.24$ (df=68; P<.001); $I^2$=46%

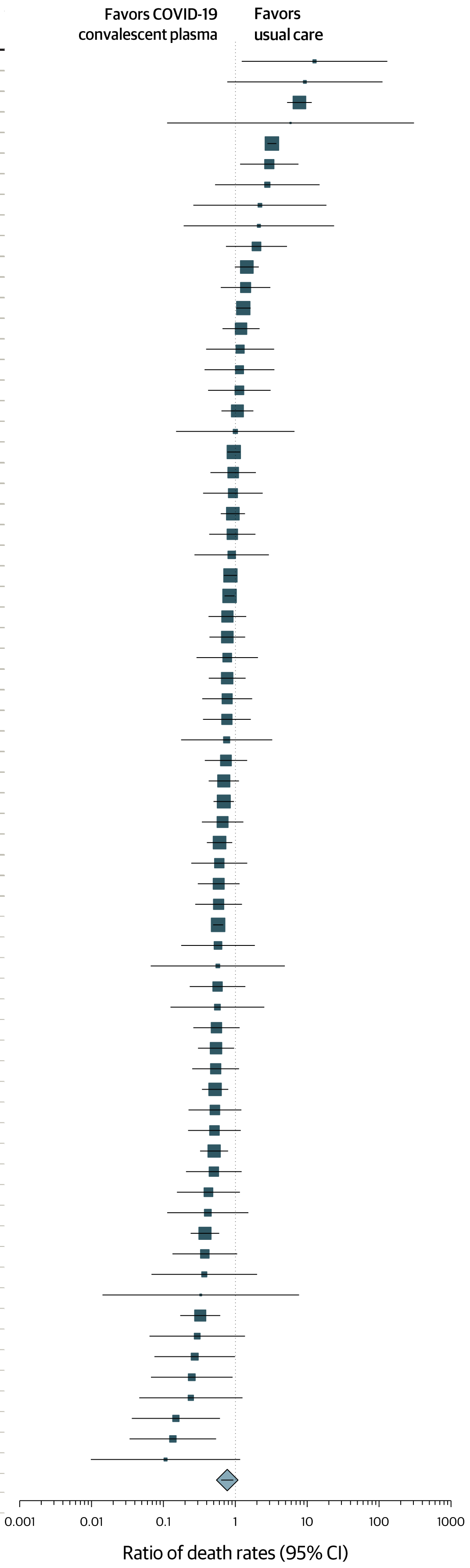

Xiao et al.,[124] 2020 not listed because no mortalities were observed in either group. The size of each square is proportional to the weight of each study. Horizontal lines indicate the 95% CI of odds ratio (OR) estimate in each study. Diamond shape represents the pooled estimate with 95% CI.

**eFigure 1. Forest plot of mortality among matched cohort studies.**

© 2022 Senefeld JW et al.

| Source | Deaths/patients randomised (%) CCP group | Usual care group | RR (95% CI) | Weight |
|---|---|---|---|---|
| Korley et al,[133] 2021 | 5/257 (2%) | 1/254 (0%) | 3.80 (0.76-18.97) | 25.9% |
| Gharbharan et al,[130] 2022 | 1/207 (0%) | 1/209 (0%) | 1.01 (0.06-16.20) | 14.8% |
| Libster et al,[134] 2021 | 2/80 (3%) | 4/80 (5%) | 0.50 (0.10-2.55) | 25.7% |
| Alemany et al,[129] 2022 | 0/188 (0%) | 2/188 (1%) | 0.14 (0.01-2.16) | 14.8% |
| Sullivan et al,[132] 2022 | 0/592 (0%) | 3/589 (1%) | 0.13 (0.01-1.29) | 18.9% |
| Total | 8/1411 (1%) | 11/1416 (1%) | 0.60 (0.16-2.28) | |

Heterogeneity: $\tau^2 = 1.094$; $\chi^2 = 7.87$ (df=4; $P$ =.10); $I^2$=49%

Shoham et al,[131] 2022 not listed because no mortalities were observed in either group. Alemany et al,[129] 2022used methylene blue treated plasma and there are concerns whether that could have affected antibody function. Different size symbols represent relative weights used in meta-analysis and are proportional to study size and study variance. Abbreviations: CCP, COVID-19 convalescent plasma; CI, confidence interval.

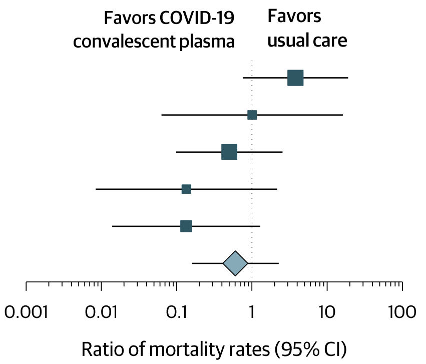

**eFigure 2**. Forest plot of mortality among outpatients with recent SARS-CoV-2 exposure or infection in randomized clinical trials.

© 2022 Senefeld JW et al.

**eTable 1. PRISMA 2020 Checklist.**

| Section and Topic | Item # | Checklist item | Location where item is reported |
|---|---|---|---|
| **TITLE** | | | |
| Title | 1 | Identify the report as a systematic review. | Title page |
| **ABSTRACT** | | | |
| Abstract | 2 | See the PRISMA 2020 for Abstracts checklist. | Structured Abstract |
| **INTRODUCTION** | | | |
| Rationale | 3 | Describe the rationale for the review in the context of existing knowledge. | Introduction |
| Objectives | 4 | Provide an explicit statement of the objective(s) or question(s) the review addresses. | Introduction |
| **METHODS** | | | |
| Eligibility criteria | 5 | Specify the inclusion and exclusion criteria for the review and how studies were grouped for the syntheses. | Eligibility Criteria |
| Information sources | 6 | Specify all databases, registers, websites, organisations, reference lists and other sources searched or consulted to identify studies. Specify the date when each source was last searched or consulted. | Information Sources |
| Search strategy | 7 | Present the full search strategies for all databases, registers and websites, including any filters and limits used. | Information Sources |
| Selection process | 8 | Specify the methods used to decide whether a study met the inclusion criteria of the review, including how many reviewers screened each record and each report retrieved, whether they worked independently, and if applicable, details of automation tools used in the process. | Selection and Data Collection Processes |
| Data collection process | 9 | Specify the methods used to collect data from reports, including how many reviewers collected data from each report, whether they worked independently, any processes for obtaining or confirming data from study investigators, and if applicable, details of automation tools used in the process. | Selection and Data Collection Processes |
| Data items | 10a | List and define all outcomes for which data were sought. Specify whether all results that were compatible with each outcome domain in each study were sought (e.g. for all measures, time points, analyses), and if not, the methods used to decide which results to collect. | Selection and Data Collection Processes |
| | 10b | List and define all other variables for which data were sought (e.g. participant and intervention characteristics, funding sources). Describe any assumptions made about any missing or unclear information. | Selection and Data Collection Processes |
| Study risk of bias assessment | 11 | Specify the methods used to assess risk of bias in the included studies, including details of the tool(s) used, how many reviewers assessed each study and whether they worked independently, and if applicable, details of automation tools used in the process. | Study Risk of Bias Assessment |
| Effect measures | 12 | Specify for each outcome the effect measure(s) (e.g. risk ratio, mean difference) used in the synthesis or presentation of results. | Statistical Analysis |
| Synthesis methods | 13a | Describe the processes used to decide which studies were eligible for each synthesis (e.g. tabulating the study intervention characteristics and comparing against the planned groups for each synthesis (item #5)). | Statistical Analysis |
| | 13b | Describe any methods required to prepare the data for presentation or synthesis, such as handling of missing summary statistics, or data | Statistical |

© 2022 Senefeld JW et al.

| Section and Topic | Item # | Checklist item | Location where item is reported |
|---|---|---|---|
| | | conversions. | Analysis |
| | 13c | Describe any methods used to tabulate or visually display results of individual studies and syntheses. | Statistical Analysis |
| | 13d | Describe any methods used to synthesize results and provide a rationale for the choice(s). If meta-analysis was performed, describe the model(s), method(s) to identify the presence and extent of statistical heterogeneity, and software package(s) used. | Statistical Analysis |
| | 13e | Describe any methods used to explore possible causes of heterogeneity among study results (e.g. subgroup analysis, meta-regression). | Statistical Analysis |
| | 13f | Describe any sensitivity analyses conducted to assess robustness of the synthesized results. | Statistical Analysis |
| Reporting bias assessment | 14 | Describe any methods used to assess risk of bias due to missing results in a synthesis (arising from reporting biases). | Risk Assessment |
| Certainty assessment | 15 | Describe any methods used to assess certainty (or confidence) in the body of evidence for an outcome. | Risk Assessment |
| **RESULTS** | | | |
| Study selection | 16a | Describe the results of the search and selection process, from the number of records identified in the search to the number of studies included in the review, ideally using a flow diagram. | Figure 1 |
| | 16b | Cite studies that might appear to meet the inclusion criteria, but which were excluded, and explain why they were excluded. | Figure 1 |
| Study characteristics | 17 | Cite each included study and present its characteristics. | Figure 2; eFigure1; eTable6 |
| Risk of bias in studies | 18 | Present assessments of risk of bias for each included study. | eTable2; eTable3 |
| Results of individual studies | 19 | For all outcomes, present, for each study: (a) summary statistics for each group (where appropriate) and (b) an effect estimate and its precision (e.g. confidence/credible interval), ideally using structured tables or plots. | Figures 2-5; eFigures 1 & 2 |
| Results of syntheses | 20a | For each synthesis, briefly summarise the characteristics and risk of bias among contributing studies. | Risk Assessment |
| | 20b | Present results of all statistical syntheses conducted. If meta-analysis was done, present for each the summary estimate and its precision (e.g. confidence/credible interval) and measures of statistical heterogeneity. If comparing groups, describe the direction of the effect. | Results |
| | 20c | Present results of all investigations of possible causes of heterogeneity among study results. | Results |
| | 20d | Present results of all sensitivity analyses conducted to assess the robustness of the synthesized results. | Results |
| Reporting biases | 21 | Present assessments of risk of bias due to missing results (arising from reporting biases) for each synthesis assessed. | Results |
| Certainty of evidence | 22 | Present assessments of certainty (or confidence) in the body of evidence for each outcome assessed. | Results |
| **DISCUSSION** | | | |
| Discussion | 23a | Provide a general interpretation of the results in the context of other evidence. | Discussion |
| | 23b | Discuss any limitations of the evidence included in the review. | Limitations |
| | 23c | Discuss any limitations of the review processes used. | Limitations |

© 2022 Senefeld JW et al.

| Section and Topic | Item # | Checklist item | Location where item is reported |
|---|---|---|---|
| | 23d | Discuss implications of the results for practice, policy, and future research. | Conclusion |
| **OTHER INFORMATION** | | | |
| Registration and protocol | 24a | Provide registration information for the review, including register name and registration number, or state that the review was not registered. | Methods |
| | 24b | Indicate where the review protocol can be accessed, or state that a protocol was not prepared. | Methods |
| | 24c | Describe and explain any amendments to information provided at registration or in the protocol. | Methods |
| Support | 25 | Describe sources of financial or non-financial support for the review, and the role of the funders or sponsors in the review. | Article Information |
| Competing interests | 26 | Declare any competing interests of review authors. | Article Information |
| Availability of data, code and other materials | 27 | Report which of the following are publicly available and where they can be found: template data collection forms; data extracted from included studies; data used for all analyses; analytic code; any other materials used in the review. | Article Information |

© 2022 Senefeld JW et al.

eTable 2. Risk of bias among randomized clinical trials.

| Source[b] | Risk of bias[a] Randomization process | Deviations from intended interventions | Missing outcome data | Measurement of the outcome | Selection of the reported result | Overall |
|---|---|---|---|---|---|---|
| Agarwal et al,[10] 2020 | Low | Some concerns | Low | Low | Low | Some concerns |
| Ali et al,[11] 2021 | Low | Low | Low | Low | Low | Low |
| AlQahtani et al,[12] 2021 | Low | Low | Low | Low | Some concerns | Some concerns |
| Avendaño-Solá et al,[13] 2021 | Low | Some concerns | Low | Low | Low | Some concerns |
| Bajpai et al,[14] 2022 | Low | Low | Low | Low | Low | Low |
| Bajpai et al,[15] 2022 | Low | Low | Low | Low | Low | Low |
| Baldeón et al,[16] 2022 | Low | Low | Low | Low | Low | Low |
| Bandopadhyay et al,[17] 2021 | Some concerns | Low | Low | Low | Low | Some concerns |
| Bar et al,[18] 2021 | Low | Low | Low | Low | Low | Low |
| Bargay-Lleonart et al,[19] 2022 | Low | Low | Low | Low | Low | Low |
| Bégin et al,[20] 2021 | Low | Some concerns | Low | Low | Low | Some concerns |
| Bennett-Guerrero et al,[21] 2021 | Low | Low | Low | Low | Low | Low |
| De Santis et al,[22] 2022 | Low | Low | Low | Low | Low | Low |
| Denkinger et al,[23] 2022 | Low | High | Low | Low | Low | High |
| Devos et al,[24] 2022 | Low | Low | Low | Low | Low | Low |
| Fernández-Sánchez et al,[25] 2022 | Some concerns | Low | Low | Low | Low | Some concerns |
| Gharbharan et al,[26] 2021 | Low | Some concerns | Low | Low | Low | Some concerns |
| Gonzalez et al,[27] 2021 | Low | Low | Low | Low | Low | Low |
| Holm et al,[28] 2021 | Some concerns | Some concerns | Low | Low | Low | Some concerns |
| Jalili et al,[29] 2022 | Low | Low | Low | Low | Low | Low |
| Kirenga et al,[30] 2021 | Low | Low | Low | Low | Low | Low |
| Körper et al,[31] 2021 | Low | High | Low | Low | Low | High |
| Lacombe et al,[32] 2022 | Low | Low | Low | Low | Low | Low |
| Li et al,[33] 2020 | Low | Some concerns | Low | Low | Low | Some concerns |
| Menichetti et al,[34] 2021 | Low | Low | Low | Low | Low | Low |
| O'Donnell et al,[35] 2021 | Low | Low | Low | Low | Low | Low |
| Ortigoza et al,[36] 2022 | Low | Low | Low | Low | Low | Low |
| Pouladzadeh et al,[37] 2021 | Low | Low | Low | Low | Low | Low |
| Rasheed et al,[38] 2020 | Low | Low | Low | Low | Low | Low |
| Ray et al,[39] 2022 | Low | Some concerns | Low | Low | Some concerns | Some concerns |
| RECOVERY,[41] 2021 | Low | Some concerns | Low | Low | Low | Some concerns |
| REMAP-CAP,[42] 2021 | Low | Low | Low | Low | Low | Low |

© 2022 Senefeld JW et al.

| | | | | | | |
|---|---|---|---|---|---|---|
| Rojas et al,[40] 2022 | Some concerns | Some concerns | Low | Low | Low | Some concerns |
| Sekine et al,[43] 2022 | Low | Low | Low | Low | Low | Low |
| Self et al,[44] 2022 | Low | Low | Low | Low | Low | Low |
| Simonovich et al,[45] 2021 | Low | Low | Low | Low | Low | Low |
| Song et al,[46] 2022 | Low | Low | Low | Low | Low | Low |
| Thorlacius-Ussing et al,[47] 2022 | Low | Low | Low | Low | Low | Low |
| van den Berg et al,[48] 2022 | Low | Low | Low | Low | Low | Low |

**Footnotes.** [a]Risk of bias assessed using the Cochrane Risk of Bias 2 tool.
[b]Reference numbers refer to references in primary text.

© 2022 Senefeld JW et al.

eTable 3. Risk of bias among matched cohort studies.

| Source[b] | Risk of bias[a] | | | |
|---|---|---|---|---|
| | Selection | Comparability | Outcome | Overall |
| Al Harthi et al,[63] 2021 | **** | ** | *** | 9 |
| Abolghasemi et al,[59] 2020 | ** | ** | *** | 7 |
| Abuzakouk et al,[60] 2021 | **** | * | *** | 8 |
| Acosta-Ampudia et al,[61] 2021 | **** | * | *** | 8 |
| Alamgir et al,[62] 2021 | *** | ** | *** | 8 |
| Allahyari et al,[64] 2021 | *** | ** | *** | 8 |
| Alsharidah et al,[65] 2021 | ** | ** | *** | 7 |
| AlShehry et al,[66] 2021 | *** | ** | *** | 8 |
| Altuntas et al,[67] 2021 | *** | * | *** | 7 |
| Arnold Egloff et al,[68] 2021 | *** | ** | *** | 8 |
| Ateş et al,[69] 2021 | *** | ** | *** | 8 |
| Biernat et al,[70] 2021 | ** | * | *** | 6 |
| Bihariesingh-Sanchit et al,[71] 2021 | **** | ** | *** | 9 |
| Briggs et al,[72] 2021 | **** | ** | *** | 9 |
| Budhiraja et al,[73] 2021 | ** | ** | *** | 7 |
| Cacilhas et al,[74] 2022 | **** | ** | *** | 9 |
| Chauhan et al,[75] 2021 | **** | ** | *** | 9 |
| Cho et al,[76] 2021 | *** | * | *** | 7 |
| Cristelli et al,[77] 2022 | *** | ** | *** | 8 |
| Dai et al,[78] 2021 | *** | * | *** | 7 |
| Donato et al,[79] 2021 | *** | * | ** | 6 |
| Duan et al,[2] 2020 | *** | ** | ** | 7 |
| Eren et al,[80] 2022 | **** | ** | *** | 9 |
| Garcia-Muñoz et al,[81] 2021 | ** | * | *** | 6 |
| Hatzl et al,[82] 2021 | **** | 0 | *** | 7 |
| Hegerova et al,[83] 2020 | ** | ** | ** | 6 |
| Hoepler et al,[84] 2021 | **** | * | *** | 8 |
| Huang et al,[85] 2021 | **** | * | *** | 8 |
| Jiang et al,[86] 2020 | *** | ** | *** | 8 |
| Khamis et al,[87] 2021 | ** | ** | *** | 7 |
| Klapholz et al,[88] 2021 | *** | ** | ** | 7 |
| Klein et al,[89] 2021 | *** | ** | *** | 8 |
| Koirala et al,[90] 2021 | *** | 0 | *** | 6 |

© 2022 Senefeld JW et al.

| | | | | |
|---|---|---|---|---|
| Kuno et al,[91] 2021 | **** | ** | *** | 9 |
| Kurnianda et al,[92] 2022 | *** | ** | *** | 8 |
| Kurtz et al,[93] 2021 | **** | * | *** | 8 |
| Lanza et al,[94] 2022 | *** | ** | *** | 8 |
| Liao et al,[95] 2021 | *** | ** | *** | 8 |
| Liu et al,[96] 2020 | **** | ** | *** | 9 |
| Mahapatra et al,[97] 2021 | **** | ** | *** | 9 |
| Mendoza et al,[98] 2021 | **** | 0 | *** | 7 |
| Mesina et al,[99] 2021 | **** | ** | *** | 9 |
| Mesina et al,[100] 2022 | **** | 0 | *** | 7 |
| Moniuszko-Malinowska et al,[101] 2020 | *** | 0 | *** | 6 |
| Novacescu et al,[102] 2022 | **** | ** | *** | 9 |
| Omrani et al,[103] 2021 | *** | ** | *** | 8 |
| Pan et al,[104] 2022 | **** | * | *** | 8 |
| Pappa et al,[105] 2021 | **** | ** | *** | 9 |
| Perotti et al,[106] 2020 | ** | * | ** | 5 |
| Rogers et al,[107] 2021 | *** | ** | *** | 8 |
| Rollas et al,[108] 2021 | **** | ** | *** | 9 |
| Romon et al,[109] 2021 | *** | ** | *** | 8 |
| Sajmi et al,[110] 2021 | *** | ** | *** | 8 |
| Salazar E et al,[111] 2021 | **** | ** | *** | 9 |
| Salazar MR et al,[112] 2021 | **** | * | *** | 8 |
| Sammartino et al,[113] 2021 | **** | ** | *** | 9 |
| Sanz et al,[114] 2022 | **** | ** | *** | 9 |
| Semedi et al,[115] 2022 | **** | * | *** | 8 |
| Shenoy et al,[116] 2021 | **** | ** | *** | 9 |
| Sostin et al,[117] 2021 | **** | ** | *** | 9 |
| Sturek et al,[118] 2022 | *** | ** | *** | 8 |
| Tang et al,[119] 2022 | **** | ** | *** | 9 |
| Thompson et al,[120] 2021 | *** | ** | *** | 8 |
| Tworek et al,[121] 2021 | **** | ** | *** | 9 |
| Weisser et al,[122] 2022 | **** | ** | *** | 9 |
| Xia et al,[123] 2020 | **** | ** | *** | 9 |
| Xiao et al,[124] 2020 | **** | 0 | *** | 7 |
| Yoon et al,[125] 2021 | *** | ** | *** | 8 |
| Zeng et al,[126] 2020 | ** | ** | *** | 7 |

© 2022 Senefeld JW et al.

| | | | | |
|---|---|---|---|---|
| Zhou et al,[127] 2022 | **** | ** | *** | 9 |

**Footnotes.** [a]Risk of bias assessed using the Newcastle-Ottawa Quality Assessment Tool for cohort studies.
[b]Reference numbers refer to references in primary text.

© 2022 Senefeld JW et al.

eTable 4. Assay systems and cutpoints used to delineate high and low antibody levels.

| Source[a] | Antibody Assay | Notes |
|---|---|---|
| Bartelt et al,[29] 2022 | On-site SARS-CoV-2 WA1 viral reporter neutralization assay | High, >1:640; Low, 1:160 to 1:640 |
| Belov et al,[33] 2022 | BROAD PRNT | High, >250; Low, <1:250 |
| Cain et al,[46] 2022 | Allermetrix Inc. Quantitative Assay | High, >8.25; Low, <8.25 |
| Fazeli et al,[82] 2022 | Euroimmun ELISA | High, >1:320; Low, <1:320 |
| Franchini et al,[89] 2022 | Inhouse Microneutralization Assay | High, >320; Low, <320 |
| Gachoud et al,[93] 2022 | Euroimmun ELISA | High, Vax-Plasma (>8 S/Co); Low, Convalescent Plasma (5 S/Co (median)) |
| González et al,[105] 2022 | Unspecified | High, >1:1600; Low, <1:1600 |
| Hemsinliogu et al,[121] 2021 | Inhouse Microneutralization Assay | High, Concentrated, pooled plasma (32.7 S/Co); Low, Single-donor Plasma (9.21 S/Co) |
| Joyner et al,[152] 2021 | Ortho-Clinical Diagnostics VITROS CLIA | High, >18.45 S/Co; Low, <4.62 S/Co |
| Khan et al,[161] 2021 | Unspecified | High, 1:160 - 1:320; Low, 1:80 - 1:160 |
| Leon et al,[179] 2022 | DiaSorin LIAISON CLIA | High: Vax-Plasma (5192 AU/mL); Low: Unvaccinated Donor Plasma (107 AU/mL) |
| Maor et al,[197] 2020 | Euroimmun ELISA | High, >4; Low, <4 |
| Mendoza et al,[98] 2021 | Sigma-Aldrich SIGMAFAST OPD ELISA | High, >1:1024; Low, <1:1024 |
| Romon et al,[109] 2021 | Ortho-Clinical Diagnostics VITROS CLIA | High, >1:250; Low, <1:250 |
| Sanz et al,[114] 2022 | Euroimmun ELISA | High, >7.6; Low, <7.6 |

Footnotes. aReference numbers in **black font** refer to Supplementary References. Reference numbers in **pink font** refer to references in primary text.

© 2022 Senefeld JW et al.

eTable 5. Cutpoints used to define early and late treatment with COVID-19 convalescent plasma.

| Source | Cutpoint | Reference | Notes |
|---|---|---|---|
| Akay Cizmecioglu et al,[146] 2021 | 5 | Admission | Early, < 5 days; Late, >5 days |
| Asem et al,[147] 2022 | 7 | Admission | Early, < 7 days; Late, >7 days |
| Ateş et al,[69] 2021 | 7 | Symptoms | Early, < 7 days; Late, > 7 days |
| Bajpai et al,[14] 2022 | 3 | Admission | Early, < 3 days; Late, > 3 days |
| Balcells et al,[148] 2021 | 7 | Enrollment | Early, Upon enrollment; Late, > 7 days |
| Briggs et al,[72] 2021 | 6 | Admission | Early, < 6 days; Late, > 6 days |
| Cain et al,[137] 2022 | 10 | Symptoms | Early, < 10 days; Late > 10 days |
| De Silvestro et al,[149] 2022 | 2 | Admission | Early, < 2 days; Late, > 2 days |
| Fazeli et al,[138] 2022 | 5 | Admission | Early, < 5 days; Late, > 5 days |
| Fodor et al,[150] 2022 | 3 | Admission | Early, < 3 days; Late, > 3 days |
| Franchini et al,[139] 2022 | 7 | Symptoms | Early, < 7 days; Late, > 7 days |
| Gazitúa et al,[96] 2020 | 7 | Symptoms | Early, < 7 days; Late, > 7 days |
| Ghadami et al,[98] 2022 | 6 | Admission | Early, < 6 days; Late, > 6 days |
| González et al,[105] 2022 | 3 | Admission | Early, < 3 days; Late, >3 days |
| Greenbaum et al,[107] 2021 | 3 | Diagnosis | Early, < 3 days; Late > 3 days |
| Hegerova et al,[83] 2020 | 7 | Admission | Early, < 7 days; Late, > 7 days |
| Hueso et al,[130] 2022 | 10 | Symptoms | Early, < 10 days; Late, > 10 days |
| Ibrahim et al,[134] 2020 | 5 | Admission | Early, < 5 days; Late, > 5 days |
| Janaka et al,[144] 2021 | 2 | Admission | Early, < 2 days; Late, >2 days |
| Jeyaraman et al,[147] 2021 | 7 | Diagnosis | Early, 7 days; Late, > 7 days |
| Kocayigit et al,[169] 2021 | 5 | Admission | Early, < 5 days; Late, > 5 days |
| Lattanzio et al,[175] 2021 | 2 | Admission | Early, < 2 days; Late, > 2 days |
| Mahapatra et al,[97] 2021 | 7 | Admission | Early, < 7 days; Late, > 7 days |
| Moniuszko-Malinowska et al,[101] 2020 | 7 | Diagnosis | Early, < 7 days; Late, > 7 days |
| RECOVERY,[41] 2021 | 7 | Symptoms | Early, < 7 days; Late, > 7 days |
| Romon et al,[109] 2021 | 7 | Symptoms | Early, < 7 days; Late, > 7 days |
| Sajmi et al,[110] 2021 | 3 | Admission | Early, < 3 days; Late, >3 days |
| Salazar et al,[111] 2021 | 3 | Admission | Early, < 3 days; Late , > 3 days |

Footnotes. [a]Reference numbers in **black font** refer to Supplementary References. Reference numbers in **pink font** refer to references in primary text.

© 2022 Senefeld JW et al.

eTable 6. Mortality among case series and case reports.

| Ref. No. | Source[a] | Survivor, n | Non-Survivor, n | Mortality, % | Patients, n |
|---|---|---|---|---|---|
| 1 | Abdullah et al | 2 | 0 | 0% | 2 |
| 2 | Abid et al | 1 | 1 | 50% | 2 |
| 3 | Acharya et al | 1 | 0 | 0% | 1 |
| 4 | Adan et al | 7 | 0 | 0% | 7 |
| 5 | Adedoyin et al | 2 | 1 | 33% | 3 |
| 6 | Agrawal et al | 87 | 54 | 38% | 141 |
| 7 | Ahn et al | 2 | 0 | 0% | 2 |
| 8 | Akay Cizmecioglu et al | 40 | 10 | 20% | 50 |
| 9 | Akilli et al | 1 | 0 | 0% | 1 |
| 10 | Aksoy et al | 1 | 0 | 0% | 1 |
| 11 | Al Helai et al | 1 | 0 | 0% | 1 |
| 12 | Ali et al | 1 | 0 | 0% | 1 |
| 13 | Amrutiya et al | 0 | 1 | 100% | 1 |
| 14 | Anderson et al | 1 | 0 | 0% | 1 |
| 15 | Antony et al | 1 | 0 | 0% | 1 |
| 16 | Anupama et al | 1 | 0 | 0% | 1 |
| 17 | Arrieta et al | 12 | 1 | 8% | 13 |
| 18 | Asem et al | 46 | 48 | 51% | 94 |
| 19 | Avanzato et al | 1 | 0 | 0% | 1 |
| 20 | Aviv et al | 1 | 0 | 0% | 1 |
| 21 | Ayed et al | 0 | 3 | 100% | 3 |
| 22 | Baang et al | 0 | 1 | 100% | 1 |
| 23 | Bader et al | 1 | 0 | 0% | 1 |
| 24 | Baek et al | 1 | 0 | 0% | 1 |
| 25 | Bakhsh et al | 1 | 0 | 0% | 1 |
| 26 | Balashov et al | 1 | 0 | 0% | 1 |
| 27 | Balcells et al | 51 | 7 | 12% | 58 |
| 28 | Bao et al | 1 | 0 | 0% | 1 |
| 29 | Bartelt et al | 42 | 13 | 24% | 55 |
| 30 | Basheer et al | 1 | 0 | 0% | 1 |
| 31 | Bayrak et al | 1 | 0 | 0% | 1 |
| 32 | Belcari et al | 1 | 0 | 0% | 1 |
| 33 | Belov et al | 17,681 | 5437 | 24% | 23,118 |
| 34 | Bereanu et al | 9 | 14 | 61% | 23 |
| 35 | Betrains et al | 4 | 1 | 20% | 5 |
| 36 | Bharat et al | 3 | 0 | 0% | 3 |
| 37 | Bhumbra et al | 1 | 1 | 50% | 2 |

 © 2022 Senefeld JW et al.

| | | | | | |
|---|---|---|---|---|---|
| 38 | Birschmann et al | 10 | 0 | 0% | 10 |
| 39 | Bobek et al | 2 | 0 | 0% | 2 |
| 40 | Bosnjak et al | 1 | 0 | 0% | 1 |
| 41 | Bradfute et al | 10 | 2 | 17% | 12 |
| 42 | Brener et al | 1 | 0 | 0% | 1 |
| 43 | Bronstein et al | 2 | 0 | 0% | 2 |
| 44 | Brown et al | 5 | 1 | 17% | 6 |
| 45 | Bruiners et al | 1 | 0 | 0% | 1 |
| 46 | Budhiraja et al | 30 | 5 | 14% | 35 |
| 47 | Cain et al | 74 | 22 | 23% | 96 |
| 48 | Camou et al | 12 | 2 | 14% | 14 |
| 49 | Carboni Bisso et al | 72 | 18 | 20% | 90 |
| 50 | Casarola et al | 1 | 0 | 0% | 1 |
| 51 | Cesilia et al | 2 | 0 | 0% | 2 |
| 52 | Chen et al | 0 | 1 | 100% | 1 |
| 53 | Cho et al | 15 | 5 | 25% | 20 |
| 54 | Choudhury et al | 1 | 0 | 0% | 1 |
| 55 | Chowdhry et al | 174 | 152 | 53% | 326 |
| 56 | Christensen et al | 0 | 1 | 100% | 1 |
| 57 | Cinar et al | 1 | 0 | 0% | 1 |
| 58 | Clark et al | 1 | 0 | 0% | 1 |
| 59 | Colombo et al | 1 | 0 | 0% | 1 |
| 60 | Cusi et al | 3 | 0 | 0% | 3 |
| 61 | D'Abramo et al | 7 | 0 | 0% | 7 |
| 62 | Dale et al | 2 | 0 | 0% | 2 |
| 63 | Das et al | 1 | 0 | 0% | 1 |
| 64 | Das et al | 1 | 0 | 0% | 1 |
| 65 | De Rienzo et al | 2 | 3 | 60% | 5 |
| 66 | De Silvestro et al | 1308 | 209 | 14% | 1517 |
| 67 | Delgado-Fernandez et al | 3 | 0 | 0% | 3 |
| 68 | Dell'Isola et al | 1 | 0 | 0% | 1 |
| 69 | Destras et al | 1 | 0 | 0% | 1 |
| 70 | Deveci et al | 3 | 0 | 0% | 3 |
| 71 | Diorio et al | 3 | 1 | 25% | 4 |
| 72 | Di Palma et al | 1 | 0 | 0% | 1 |
| 73 | Donzelli et al | 1 | 0 | 0% | 1 |
| 74 | Duan et al | 75 | 0 | 0% | 75 |
| 75 | Dulipsingh et al | 20 | 19 | 49% | 39 |
| 76 | Easterlin et al | 1 | 0 | 0% | 1 |
| 77 | Einollahi et al | 1 | 0 | 0% | 1 |

© 2022 Senefeld JW et al.

| | | | | | |
|---|---|---|---|---|---|
| 78 | Elliott et al | 14 | 0 | 0% | 14 |
| 79 | Erber et al | 2 | 4 | 67% | 6 |
| 80 | Erkurt et al | 20 | 6 | 23% | 26 |
| 81 | Faltin et al | 1 | 0 | 0% | 1 |
| 82 | Fanning et al | 26 | 9 | 26% | 35 |
| 83 | Fazeli et al | 2190 | 907 | 29% | 3097 |
| 84 | Ferrari et al | 7 | 0 | 0% | 7 |
| 85 | Figlerowicz et al | 1 | 0 | 0% | 1 |
| 86 | Fisher et al | 1 | 0 | 0% | 1 |
| 87 | Fodor et al | 183 | 84 | 31% | 267 |
| 88 | Fox et al | 1 | 0 | 0% | 1 |
| 89 | Franchini et al | 54 | 0 | 0% | 54 |
| 90 | Franchini et al | 19 | 3 | 14% | 22 |
| 91 | Franchini et al | 354 | 51 | 13% | 405 |
| 92 | Fung et al | 4 | 0 | 0% | 4 |
| 93 | Furlan et al | 4 | 0 | 0% | 4 |
| 94 | Gachoud et al | 34 | 2 | 6% | 36 |
| 95 | Garcia-Vidal et al | 0 | 1 | 100% | 1 |
| 96 | Gattuso et al | 1 | 0 | 0% | 1 |
| 97 | Gazitua et al | 161 | 31 | 16% | 192 |
| 98 | Gemici et al | 25 | 15 | 38% | 40 |
| 99 | Ghadami et al | 65 | 24 | 27% | 89 |
| 100 | Gharbharan et al | 21 | 4 | 16% | 25 |
| 101 | Ghimire et al | 19 | 2 | 10% | 21 |
| 102 | Gibson et al | 1 | 0 | 0% | 1 |
| 103 | Gibson et al | 1 | 0 | 0% | 1 |
| 104 | Gomes et al | 1 | 0 | 0% | 1 |
| 105 | Gonzalez et al | 201 | 71 | 26% | 272 |
| 106 | González et al | 3647 | 1072 | 23% | 4719 |
| 107 | Gordon et al | 14 | 0 | 0% | 14 |
| 108 | Greenbaum et al | 32 | 12 | 27% | 44 |
| 109 | Grisolia et al | 1 | 0 | 0% | 1 |
| 110 | Guo et al | 1 | 0 | 0% | 1 |
| 111 | Gupta et al | 9 | 1 | 10% | 10 |
| 112 | Hacibekiroglu et al | 12 | 16 | 57% | 28 |
| 113 | Hahn et al | 1 | 0 | 0% | 1 |
| 114 | Halfmann et al | 1 | 0 | 0% | 1 |
| 115 | Haller et al | 1 | 0 | 0% | 1 |
| 116 | Hanssen et al | 1 | 0 | 0% | 1 |
| 117 | Hartman et al | 1 | 0 | 0% | 1 |

© 2022 Senefeld JW et al.

| | | | | | |
|---|---|---|---|---|---|
| 118 | Hartman et al | 27 | 4 | 13% | 31 |
| 119 | Hartman et al | 1 | 0 | 0% | 1 |
| 120 | He et al | 37 | 1 | 3% | 38 |
| 121 | Helleberg et al | 8 | 2 | 20% | 10 |
| 122 | Hemsinlioglu et al | 11 | 5 | 31% | 16 |
| 123 | Herman et al | 14 | 5 | 26% | 19 |
| 124 | Honjo et al | 1 | 0 | 0% | 1 |
| 125 | Hopwood et al | 1 | 0 | 0% | 1 |
| 126 | Hovey et al | 1 | 0 | 0% | 1 |
| 127 | Hu et al | 1 | 0 | 0% | 1 |
| 128 | Hu et al | 7 | 0 | 0% | 7 |
| 129 | Huang et al | 3 | 0 | 0% | 3 |
| 130 | Huang et al | 20 | 4 | 17% | 24 |
| 131 | Hueso et al | 16 | 1 | 6% | 17 |
| 132 | Hueso et al | 73 | 39 | 35% | 112 |
| 133 | Hughes et al | 1 | 0 | 0% | 1 |
| 134 | Iaboni et al | 1 | 0 | 0% | 1 |
| 135 | Ibrahim et al | 24 | 14 | 37% | 38 |
| 136 | Iglesias et al | 27 | 44 | 62% | 71 |
| 137 | Im et al | 1 | 0 | 0% | 1 |
| 138 | Ipe et al | 127 | 38 | 23% | 165 |
| 139 | Jacobs et al | 43 | 56 | 57% | 99 |
| 140 | Jacobson et al | 1 | 0 | 0% | 1 |
| 141 | Jafari et al | 1 | 0 | 0% | 1 |
| 142 | Jaiswal et al | 10 | 4 | 29% | 14 |
| 143 | Jamir et al | 1 | 0 | 0% | 1 |
| 144 | Jamous et al | 24 | 5 | 17% | 29 |
| 145 | Janaka et al | 37 | 7 | 16% | 44 |
| 146 | Jassem et al | 1 | 0 | 0% | 1 |
| 147 | Jasuja et al | 7 | 15 | 68% | 22 |
| 148 | Jeyaraman et al | 18 | 15 | 45% | 33 |
| 149 | Ji et al | 8 | 0 | 0% | 8 |
| 150 | Jiang et al | 1 | 0 | 0% | 1 |
| 151 | Jin et al | 6 | 0 | 0% | 6 |
| 152 | Jin et al | 3 | 0 | 0% | 3 |
| 153 | Joyner et al | 2252 | 830 | 27% | 3082 |
| 154 | Karaolidou et al | 1 | 0 | 0% | 1 |
| 155 | Karatas et al | 1 | 0 | 0% | 1 |
| 156 | Katz-Greenberg et al | 4 | 0 | 0% | 4 |
| 157 | Keitel et al | 1 | 0 | 0% | 1 |

© 2022 Senefeld JW et al.

| | | | | | |
|---|---|---|---|---|---|
| 158 | Kemp et al | 0 | 1 | 100% | 1 |
| 159 | Kenig et al | 8 | 0 | 0% | 8 |
| 160 | Ketels et al | 1 | 0 | 0% | 1 |
| 161 | Khamis et al | 91 | 19 | 17% | 110 |
| 162 | Khan et al | 1 | 0 | 0% | 1 |
| 163 | Khan et al | 42 | 8 | 16% | 50 |
| 164 | Khatamzas et al | 2 | 1 | 33% | 3 |
| 165 | Khatri et al | 0 | 1 | 100% | 1 |
| 166 | Khot et al | 1 | 0 | 0% | 1 |
| 167 | King et al | 1 | 0 | 0% | 1 |
| 168 | Kluger et al | 3 | 0 | 0% | 3 |
| 169 | Ko et al | 51 | 21 | 29% | 72 |
| 170 | Kocayigit et al | 69 | 72 | 51% | 141 |
| 171 | Kochanek et al | 1 | 0 | 0% | 1 |
| 172 | Kong et al | 1 | 0 | 0% | 1 |
| 173 | Kremer et al | 3 | 0 | 0% | 3 |
| 174 | Lancman et al | 1 | 0 | 0% | 1 |
| 175 | Lang-Meli et al | 16 | 0 | 0% | 16 |
| 176 | Lattanzio et al | 151 | 46 | 23% | 197 |
| 177 | Lazzari et al | 1 | 0 | 0% | 1 |
| 178 | Lee et al | 32 | 2 | 6% | 34 |
| 179 | Lemus et al | 1 | 0 | 0% | 1 |
| 180 | Leon et al | 25 | 0 | 0% | 25 |
| 181 | Levy et al | 33 | 16 | 33% | 49 |
| 182 | Lima et al | 2 | 0 | 0% | 2 |
| 183 | Lindemann et al | 3 | 1 | 25% | 4 |
| 184 | Lindemann et al | 3 | 5 | 63% | 8 |
| 185 | Liu et al | 1 | 0 | 0% | 1 |
| 186 | Liu et al | 3 | 0 | 0% | 3 |
| 187 | Liu et al | 1 | 0 | 0% | 1 |
| 188 | Ljungquist et al | 22 | 6 | 21% | 28 |
| 189 | London et al | 1 | 0 | 0% | 1 |
| 190 | López et al | 9 | 1 | 10% | 10 |
| 191 | Lubnow et al | 1 | 0 | 0% | 1 |
| 192 | Luetkens et al | 1 | 0 | 0% | 1 |
| 193 | Madariaga et al | 9 | 1 | 10% | 10 |
| 194 | Magallanes-Garza et al | 1 | 0 | 0% | 1 |
| 195 | Magyari et al | 20 | 0 | 0% | 20 |
| 196 | Malecki et al | 13 | 0 | 0% | 13 |
| 197 | Malsy et al | 1 | 0 | 0% | 1 |

© 2022 Senefeld JW et al.

| | | | | | |
|---|---|---|---|---|---|
| 198 | Maor et al | 40 | 9 | 18% | 49 |
| 199 | Marconato et al | 29 | 1 | 3% | 30 |
| 200 | Martens et al | 1 | 0 | 0% | 1 |
| 201 | Martínez-Barranco et al | 4 | 1 | 20% | 5 |
| 202 | Martínez-Chinchilla et al | 2 | 0 | 0% | 2 |
| 203 | Martinez-Resendez et al | 8 | 0 | 0% | 8 |
| 204 | Martinot et al | 1 | 0 | 0% | 1 |
| 205 | Marwah et al | 1 | 0 | 0% | 1 |
| 206 | Mastroianni et al | 2 | 0 | 0% | 2 |
| 207 | McCuddy et al | 3 | 0 | 0% | 3 |
| 208 | McKemey et al | 1 | 0 | 0% | 1 |
| 209 | Mehta et al | 1 | 1 | 50% | 2 |
| 210 | Mekonnen et al | 0 | 1 | 100% | 1 |
| 211 | Mendes-Correa et al | 1 | 0 | 0% | 1 |
| 212 | Merin et al | 8 | 2 | 20% | 10 |
| 213 | Meshram et al | 5 | 0 | 0% | 5 |
| 214 | Milosevic et al | 1 | 0 | 0% | 1 |
| 215 | Milota et al | 4 | 0 | 0% | 4 |
| 216 | Mira et al | 1 | 0 | 0% | 1 |
| 217 | Mirihagalle et al | 1 | 0 | 0% | 1 |
| 218 | Mohanty et al | 66 | 111 | 63% | 177 |
| 219 | Mohseni et al | 1 | 0 | 0% | 1 |
| 220 | Moniuszko-Malinowska et al | 1 | 0 | 0% | 1 |
| 221 | Monrad et al | 1 | 0 | 0% | 1 |
| 222 | Moore et al | 1 | 0 | 0% | 1 |
| 223 | Moura et al | 1 | 0 | 0% | 1 |
| 224 | Moutinho-Pereira et al | 1 | 0 | 0% | 1 |
| 225 | Mukhina et al | 1 | 0 | 0% | 1 |
| 226 | Mullaguri et al | 0 | 2 | 100% | 2 |
| 227 | Naeem et al | 3 | 0 | 0% | 3 |
| 228 | Negi et al | 26 | 37 | 59% | 63 |
| 229 | Ng et al | 2 | 1 | 33% | 3 |
| 230 | Nguyen et al | 2 | 0 | 0% | 2 |
| 231 | Niu et al | 1 | 0 | 0% | 1 |
| 232 | Novacescu et al | 1 | 0 | 0% | 1 |
| 233 | Nowak et al | 1 | 0 | 0% | 1 |
| 234 | Nussenblatt et al | 1 | 0 | 0% | 1 |
| 235 | Nyström et al | 1 | 0 | 0% | 1 |
| 236 | Oehler et al | 1 | 0 | 0% | 1 |
| 237 | Oliva et al | 5 | 1 | 17% | 6 |

© 2022 Senefeld JW et al.

| | | | | | |
|---|---|---|---|---|---|
| 238 | Olivares et al | 1 | 0 | 0% | 1 |
| 239 | Olivares-Gazca et al | 8 | 2 | 20% | 10 |
| 240 | Oliveira et al | 40 | 9 | 18% | 49 |
| 241 | Ordaya et al | 1 | 0 | 0% | 1 |
| 242 | Ormazabal Velez et al | 2 | 0 | 0% | 2 |
| 243 | Padilla et al | 74 | 21 | 22% | 95 |
| 244 | Pagan et al | 7 | 0 | 0% | 7 |
| 245 | Pal et al | 15 | 2 | 12% | 17 |
| 246 | Pei et al | 19 | 0 | 0% | 19 |
| 247 | Pelayo et al | 1 | 0 | 0% | 1 |
| 248 | Peng et al | 1 | 0 | 0% | 1 |
| 249 | Peppers et al | 76 | 26 | 25% | 102 |
| 250 | Plotnikov et al | 30 | 10 | 25% | 40 |
| 251 | Pommeret et al | 4 | 1 | 20% | 5 |
| 252 | Prasad et al | 1 | 0 | 0% | 1 |
| 253 | Radhakrishnan Nair et al | 1 | 0 | 0% | 1 |
| 254 | Ragab et al | 1 | 0 | 0% | 1 |
| 255 | Rahardjo et al | 8 | 0 | 0% | 8 |
| 256 | Rahimi-Levene et al | 92 | 20 | 18% | 112 |
| 257 | Rahman et al | 10 | 3 | 23% | 13 |
| 258 | Ramalingam et al | 1 | 0 | 0% | 1 |
| 259 | Rejeki et al | 8 | 2 | 20% | 10 |
| 260 | Ribeiro et al | 1 | 0 | 0% | 1 |
| 261 | Ripoll et al | 26 | 5 | 16% | 31 |
| 262 | Rizvi et al | 1 | 0 | 0% | 1 |
| 263 | Rnjak et al | 1 | 0 | 0% | 1 |
| 264 | Rodriguez et al | 0 | 1 | 100% | 1 |
| 265 | Rodriguez et al | 1 | 0 | 0% | 1 |
| 266 | Rodriguez-Pla et al | 1 | 0 | 0% | 1 |
| 267 | Roshandel et al | 4 | 1 | 20% | 5 |
| 268 | Rosner-Tenerowicz et al | 1 | 0 | 0% | 1 |
| 269 | Rüfenacht et al | 1 | 0 | 0% | 1 |
| 270 | Saad Shaukat et al | 1 | 0 | 0% | 1 |
| 271 | Saha et al | 3 | 5 | 63% | 8 |
| 272 | Sait et al | 41 | 3 | 7% | 44 |
| 273 | Salazar et al | 24 | 1 | 4% | 25 |
| 274 | Sampaio et al | 1 | 0 | 0% | 1 |
| 275 | Schenker et al | 1 | 0 | 0% | 1 |
| 276 | Schreiber et al | 1 | 0 | 0% | 1 |
| 277 | Schwartz et al | 4 | 0 | 0% | 4 |

© 2022 Senefeld JW et al.

| | | | | | |
|---|---|---|---|---|---|
| 278 | Senefeld et al | 70527 | 23760 | 25% | 94287 |
| 279 | Sepulcri et al | 0 | 1 | 100% | 1 |
| 280 | Sethi et al | 3 | 3 | 50% | 6 |
| 281 | Shah et al | 1 | 0 | 0% | 1 |
| 282 | Shaikh et al | 1 | 0 | 0% | 1 |
| 283 | Shankar et al | 1 | 0 | 0% | 1 |
| 284 | Shankar et al | 1 | 0 | 0% | 1 |
| 285 | Sharma et al | 52 | 8 | 13% | 60 |
| 286 | Shen et al | 5 | 0 | 0% | 5 |
| 287 | Shields et al | 3 | 2 | 40% | 5 |
| 288 | Sobelman et al | 1 | 0 | 0% | 1 |
| 289 | Soleimani et al | 1 | 0 | 0% | 1 |
| 290 | Spinicci et al | 1 | 0 | 0% | 1 |
| 291 | Spinicci et al | 1 | 0 | 0% | 1 |
| 292 | Steiner et al | 1 | 1 | 50% | 2 |
| 293 | Swenson et al | 41 | 8 | 16% | 49 |
| 294 | Szwebel et al | 1 | 0 | 0% | 1 |
| 295 | Taha et al | 1 | 0 | 0% | 1 |
| 296 | Tan et al | 1 | 0 | 0% | 1 |
| 297 | Thabrani et al | 2 | 0 | 0% | 2 |
| 298 | Thomopoulos et al | 29 | 2 | 6% | 31 |
| 299 | Thota et al | 1 | 0 | 0% | 1 |
| 300 | Tirnea et al | 2 | 3 | 60% | 5 |
| 301 | Tran et al | 1 | 0 | 0% | 1 |
| 302 | Tremblay et al | 14 | 10 | 42% | 24 |
| 303 | Trimarchi et al | 1 | 0 | 0% | 1 |
| 304 | Umapathi et al | 1 | 0 | 0% | 1 |
| 305 | Valentini et al | 70 | 17 | 20% | 87 |
| 306 | Van Damme et al | 1 | 0 | 0% | 1 |
| 307 | van Oers et al | 1 | 0 | 0% | 1 |
| 308 | Vlachogianni et al | 0 | 1 | 100% | 1 |
| 309 | Wang et al | 1 | 0 | 0% | 1 |
| 310 | Wang et al | 2 | 3 | 60% | 5 |
| 311 | Wei et al | 2 | 0 | 0% | 2 |
| 312 | Weinbergerova et al | 29 | 3 | 9% | 32 |
| 313 | Weinbergerova et al | 16 | 3 | 16% | 19 |
| 314 | Werthman-Ehrenreich et al | 0 | 1 | 100% | 1 |
| 315 | Whang et al | 12 | 4 | 25% | 16 |
| 316 | Wirz et al | 12 | 4 | 25% | 16 |
| 317 | Wright et al | 1 | 0 | 0% | 1 |

© 2022 Senefeld JW et al.

| | | | | | |
|---|---|---|---|---|---|
| 318 | Wu et al | 24 | 3 | 11% | 27 |
| 319 | Xu et al | 1 | 0 | 0% | 1 |
| 320 | Yang et al | 7 | 3 | 30% | 10 |
| 321 | Yang et al | 1 | 0 | 0% | 1 |
| 322 | Yaqoub et al | 1 | 0 | 0% | 1 |
| 323 | Yatam Ganesh et al | 2 | 1 | 33% | 3 |
| 324 | Ye et al | 6 | 0 | 0% | 6 |
| 325 | Yi et al | 1 | 0 | 0% | 1 |
| 326 | Yokoyama et al | 91 | 11 | 11% | 102 |
| 327 | Yulianti et al | 2 | 0 | 0% | 2 |
| 328 | Zeng et al | 8 | 0 | 0% | 8 |
| 329 | Zhang et al | 4 | 0 | 0% | 4 |
| 330 | Zhang et al | 1 | 0 | 0% | 1 |
| 331 | Zhang et al | 2 | 0 | 0% | 2 |
| 332 | Zhang et al | 1 | 0 | 0% | 1 |
| 333 | Zimmerli et al | 1 | 0 | 0% | 1 |
| 334 | Zimmermann et al | 0 | 1 | 100% | 1 |
| 335 | Zlamal et al | 1 | 0 | 0% | 1 |
| | **Total** | 102178 | 33771 | 24.8% | 135949 |

Footnotes. [a]Reference numbers (ref no.) refer to Supplementary References.

© 2022 Senefeld JW et al.

### Supplementary References

1. Abdullah HM, Hama-Ali HH, Ahmed SN, et al. A severe refractory COVID-19 patient responding to convalescent plasma; A case series. *Ann Med Surg (Lond).* 2020;56:125-127.
2. Abid MB, Chhabra S, Buchan B, et al. Bronchoalveolar lavage-based COVID-19 testing in patients with cancer. *Hematol Oncol Stem Cell Ther.* 2021;14(1):65-70.
3. Acharya S, Thibault M, Lee J, et al. COVID-19-Induced Left Sciatic Neuropathy Requiring Prolonged Physical Medicine and Rehabilitation. *Cureus.* 2021;13(6):e15803.
4. Adan H, Harb D, Hazari K, et al. Use of convalescent plasma in pregnant women with early stage COVID-19 infection in a tertiary care hospital in Dubai, February to March 2021: a case series study. *BMC Pregnancy Childbirth.* 2022;22(1):730.
5. Adedoyin O, Brijmohan S, Lavine R, Lisung FG. Undetectable SARS-CoV-2 active adaptive immunity-post-vaccination or post-COVID-19 severe disease-after immunosuppressants use. *BMJ Case Rep.* 2021;14(11).
6. Agrawal A, Jha T, Gogoi P, et al. Effect of convalescent plasma therapy on mortality in moderate-to-severely Ill COVID-19 patients. *Transfus Apher Sci.* 2022;61(6):103455.
7. Ahn JY, Sohn Y, Lee SH, et al. Use of Convalescent Plasma Therapy in Two COVID-19 Patients with Acute Respiratory Distress Syndrome in Korea. *J Korean Med Sci.* 2020;35(14):e149.
8. Akay Cizmecioglu H, Goktepe MH, Demircioglu S, et al. Efficacy of convalescent plasma therapy in severe COVID-19 patients. *Transfus Apher Sci.* 2021;60(4):103158.
9. Akilli NB, Yosunkaya A. Part of the Covid19 puzzle: Acute parkinsonism. *Am J Emerg Med.* 2021;47:333 e331-333 e333.
10. Aksoy E, Oztutgan T. COVID-19 Presentation in Association with Myasthenia Gravis: A Case Report and Review of the Literature. *Case Rep Infect Dis.* 2020;2020:8845844.
11. Al Helali AA, Saeed GA, Elholiby TI, Kukkady MA, Mazrouei SSA. Radiological and clinical improvement in a patient with COVID-19 pneumonia postconvalescent plasma transfusion: A case report. *Radiol Case Rep.* 2020;15(11):2171-2174.
12. Ali S, Khalid S, Afridi M, Akhtar S, Khader YS, Akhtar H. Notes From the Field: The Combined Effects of Tocilizumab and Remdesivir in a Patient With Severe COVID-19 and Cytokine Release Syndrome. *JMIR Public Health Surveill.* 2021;7(5):e27609.
13. Amrutiya V, Patel R, Baghal M, et al. Transfusion-related acute lung injury in a COVID-19-positive convalescent plasma recipient: a case report. *J Int Med Res.* 2021;49(8):3000605211032814.
14. Anderson J, Schauer J, Bryant S, Graves CR. The use of convalescent plasma therapy and remdesivir in the successful management of a critically ill obstetric patient with novel coronavirus 2019 infection: A case report. *Case Rep Womens Health.* 2020;27:e00221.
15. Antony SJ, Singh J, de Jesus M, Lance J. Early use of tocilizumab in respiratory failure associated with acute COVID -19 pneumonia in recipients with solid organ transplantation. *IDCases.* 2020;21:e00888.
16. Anupama BK, Thapa SS, Amzuta I. Transient Cardiomyopathy in a Patient With Coronavirus Disease-2019. *J Investig Med High Impact Case Rep.* 2020;8:2324709620947577.
17. Arrieta A, Galvis AE, Morphew T, et al. Safety and Antibody Kinetics of COVID-19 Convalescent Plasma for the Treatment of Moderate to Severe Cases of SARS-CoV-2 Infection in Pediatric Patients. *Pediatr Infect Dis J.* 2021;40(7):606-611.
18. Asem N, Massoud HH, Serag I, et al. Clinical efficacy of Early Administration of Convalescent Plasma among COVID-19 Cases in Egypt. *medRxiv.* 2021.
19. Avanzato VA, Matson MJ, Seifert SN, et al. Case Study: Prolonged Infectious SARS-CoV-2 Shedding from an Asymptomatic Immunocompromised Individual with Cancer. *Cell.* 2020;183(7):1901-1912 e1909.

© 2022 Senefeld JW et al.

20. Aviv R, Weber A, Anzum T, Federbush M, Horowitz D, Singas E. Prolonged Coronavirus Disease 2019 in a Patient With Rheumatoid Arthritis on Rituximab Therapy. *J Infect Dis.* 2021;224(3):557-559.
21. Ayed M, Borahmah AA, Yazdani A, Sultan A, Mossad A, Rawdhan H. Assessment of Clinical Characteristics and Mortality-Associated Factors in COVID-19 Critical Cases in Kuwait. *Med Princ Pract.* 2021;30(2):185-192.
22. Baang JH, Smith C, Mirabelli C, et al. Prolonged Severe Acute Respiratory Syndrome Coronavirus 2 Replication in an Immunocompromised Patient. *J Infect Dis.* 2021;223(1):23-27.
23. Bader N, Khattab M, Farah F. Severe reinfection with severe acute respiratory syndrome coronavirus 2 in a nursing home resident: a case report. *J Med Case Rep.* 2021;15(1):392.
24. Baek AR, Choo EJ, Kim JY, et al. A Transient Effect of Convalescent Plasma Therapy in a Patient with Severe Covonavirus Disease 2019: A Case Report. *Infect Chemother.* 2022;54(3):553-558.
25. Bakhsh A, AlSaeed M, Ibrahim MA, et al. Recovery From COVID-19 Pneumonia in a Heart Transplant Recipient: A Case Report. *Infect Dis Clin Pract (Baltim Md).* 2021;29(6):e401-e403.
26. Balashov D, Trakhtman P, Livshits A, et al. SARS-CoV-2 convalescent plasma therapy in pediatric patient after hematopoietic stem cell transplantation. *Transfus Apher Sci.* 2021;60(1):102983.
27. Balcells ME, Rojas L, Le Corre N, et al. Early versus deferred anti-SARS-CoV-2 convalescent plasma in patients admitted for COVID-19: A randomized phase II clinical trial. *PLoS Med.* 2021;18(3):e1003415.
28. Bao Y, Lin SY, Cheng ZH, et al. Clinical Features of COVID-19 in a Young Man with Massive Cerebral Hemorrhage-Case Report. *SN Compr Clin Med.* 2020;2(6):703-709.
29. Bartelt LA, Markmann AJ, Nelson B, et al. Outcomes of Convalescent Plasma with Defined High versus Lower Neutralizing Antibody Titers against SARS-CoV-2 among Hospitalized Patients: CoronaVirus Inactivating Plasma (CoVIP) Study. *mBio.* 2022;13(5):e0175122.
30. Basheer M, Saad E, Laskar O, et al. Clearance of the SARS-CoV-2 Virus in an Immunocompromised Patient Mediated by Convalescent Plasma without B-Cell Recovery. *Int J Mol Sci.* 2021;22(16).
31. Bayrak M, Cadirci K. Successful pulsed methylprednisolone and convalescent plasma treatment in a case of a renal transplant recipient with COVID-19 positive pneumonia: a case report. *Pan Afr Med J.* 2021;38:273.
32. Belcari G, Conti A, Mazzoni A, Lanza M, Mazzetti P, Focosi D. Clinical and Virological Response to Convalescent Plasma in a Chronic Lymphocytic Leukemia Patient with COVID-19 Pneumonia. *Life (Basel).* 2022;12(7).
33. Belov A, Huang Y, Villa CH, et al. Early administration of COVID-19 convalescent plasma with high titer antibody content by live viral neutralization assay is associated with modest clinical efficacy. *Am J Hematol.* 2022;97(6):770-779.
34. Bereanu A, Crisan O, Constantin AM, et al. The Effect of Convalescent Plasma in Patients With Covid-19 in Intensive Care Unit. *In Vivo.* 2022;36(3):1342-1348.
35. Betrains A, Godinas L, Woei AJF, et al. Convalescent plasma treatment of persistent severe acute respiratory syndrome coronavirus-2 (SARS-CoV-2) infection in patients with lymphoma with impaired humoral immunity and lack of neutralising antibodies. *Br J Haematol.* 2021;192(6):1100-1105.
36. Bharat A, Querrey M, Markov NS, et al. Lung transplantation for patients with severe COVID-19. *Sci Transl Med.* 2020;12(574).
37. Bhumbra S, Malin S, Kirkpatrick L, et al. Clinical Features of Critical Coronavirus Disease 2019 in Children. *Pediatr Crit Care Med.* 2020;21(10):e948-e953.
38. Birschmann I, von Bargen K, Teune M, Flottmann C, Knuttgen F, Knabbe C. Retrospective study shows that early administration of convalescent plasma in hospitalized COVID-19 patients may have a positive effect on disease progression. *Health Sci Rep.* 2022;5(5):e714.

© 2022 Senefeld JW et al.

39. Bobek I, Gopcsa L, Reti M, et al. [Successful administration of convalescent plasma in critically ill COVID-19 patients in Hungary: the first two cases]. *Orv Hetil.* 2020;161(27):1111-1121.
40. Bosnjak B, Odak I, Ritter C, et al. Case Report: Convalescent Plasma Therapy Induced Anti-SARS-CoV-2 T Cell Expansion, NK Cell Maturation and Virus Clearance in a B Cell Deficient Patient After CD19 CAR T Cell Therapy. *Front Immunol.* 2021;12:721738.
41. Bradfute SB, Hurwitz I, Yingling AV, et al. Severe Acute Respiratory Syndrome Coronavirus 2 Neutralizing Antibody Titers in Convalescent Plasma and Recipients in New Mexico: An Open Treatment Study in Patients With Coronavirus Disease 2019. *J Infect Dis.* 2020;222(10):1620-1628.
42. Brener ZZ, Brenner A. Effectiveness of convalescent plasma therapy in a patient with severe COVID-19-associated acute kidney injury. *Clin Nephrol Case Stud.* 2021;9:67-71.
43. Bronstein Y, Adler A, Katash H, Halutz O, Herishanu Y, Levytskyi K. Evolution of spike mutations following antibody treatment in two immunocompromised patients with persistent COVID-19 infection. *J Med Virol.* 2022;94(3):1241-1245.
44. Brown LK, Moran E, Goodman A, et al. Treatment of chronic or relapsing COVID-19 in immunodeficiency. *J Allergy Clin Immunol.* 2022;149(2):557-561 e551.
45. Bruiners N, Guerrini V, Ukey R, et al. Longitudinal Analysis of Biologic Correlates of COVID-19 Resolution: Case Report. *Front Med (Lausanne).* 2022;9:915367.
46. Budhiraja S, Soni A, Jha V, et al. Clinical Profile of First 1000 COVID-19 Cases Admitted at Tertiary Care Hospitals and the Correlates of their Mortality: An Indian Experience. *medRxiv.* 2020:2020.2011.2016.20232223.
47. Cain WV, Sill AM, Solipuram V, Weiss JJ, Miller CB, Jelsma PF. Efficacy of COVID-19 Convalescent Plasma Based on Antibody Concentration. *Adv Hematol.* 2022;2022:7992927.
48. Camou F, Tinevez C, Beguet-Yachine M, et al. Feasibility of convalescent plasma therapy in severe COVID-19 patients with persistent SARS-CoV-2 viremia. *J Med Virol.* 2021;93(9):5594-5598.
49. Carboni Bisso I, Huespe I, Lockhart C, et al. Clinical characteristics of critically ill patients with COVID-19. *Medicina (B Aires).* 2021;81(4):527-535.
50. Casarola G, D'Abbondanza M, Curcio R, et al. Efficacy of convalescent plasma therapy in immunocompromised patients with COVID-19: A case report. *Clin Infect Pract.* 2021;12:100096.
51. Cesilia C, Ridar E, Suryawan N, Nataprawira HM. Convalescent plasma therapy in obese severe COVID-19 adolescents: Two cases report. *Ann Med Surg (Lond).* 2021;72:103084.
52. Chen L, Zody MC, Di Germanio C, et al. Emergence of Multiple SARS-CoV-2 Antibody Escape Variants in an Immunocompromised Host Undergoing Convalescent Plasma Treatment. *mSphere.* 2021;6(4):e0048021.
53. Cho Y, Sohn Y, Hyun J, et al. Effectiveness of Convalescent Plasma Therapy in Severe or Critically Ill COVID-19 Patients: A Retrospective Cohort Study. *Yonsei Med J.* 2021;62(9):799-805.
54. Choudhury A, Reddy GS, Venishetty S, et al. COVID-19 in Liver Transplant Recipients - A Series with Successful Recovery. *J Clin Transl Hepatol.* 2020;8(4):467-473.
55. Chowdhry M, Hussain M, Singh P, et al. Convalescent plasma - An insight into a novel treatment of covid-19 ICU patients. *Transfus Apher Sci.* 2022;61(6):103497.
56. Christensen J, Kumar D, Moinuddin I, et al. Coronavirus Disease 2019 Viremia, Serologies, and Clinical Course in a Case Series of Transplant Recipients. *Transplant Proc.* 2020;52(9):2637-2641.
57. Cinar OE, Sayinalp B, Aladag Karakulak E, et al. Convalescent (immune) plasma treatment in a myelodysplastic COVID-19 patient with disseminated tuberculosis. *Transfus Apher Sci.* 2020;59(5):102821.

 © 2022 Senefeld JW et al.

58. Clark E, Guilpain P, Filip IL, et al. Convalescent plasma for persisting COVID-19 following therapeutic lymphocyte depletion: a report of rapid recovery. *Br J Haematol.* 2020;190(3):e154-e156.
59. Colombo D, Gatti A, Alabardi P, et al. COVID-19-Associated Pneumonia in a B-Cell-Depleted Patient With Non-Hodgkin Lymphoma: Recovery With Hyperimmune Plasma. *J Hematol.* 2022;11(2):77-80.
60. Cusi MG, Conticini E, Gandolfo C, et al. Hyperimmune plasma in three immuno-deficient patients affected by non-severe, prolonged COVID-19: a single-center experience. *BMC Infect Dis.* 2021;21(1):630.
61. D'Abramo A, Vita S, Maffongelli G, et al. Clinical Management of Patients With B-Cell Depletion Agents to Treat or Prevent Prolonged and Severe SARS-COV-2 Infection: Defining a Treatment Pathway. *Front Immunol.* 2022;13:911339.
62. Dale M, Sogawa H, Seyedsaadat SM, et al. Successful Management of COVID-19 Infection in 2 Early Post-Liver Transplant Recipients. *Transplant Proc.* 2021;53(4):1175-1179.
63. Das SK, Ranabhat K, Bhattarai S, et al. Combination of convalescent plasma therapy and repurposed drugs to treat severe COVID-19 patient with multimorbidity. *Clin Case Rep.* 2021;9(4):2132-2137.
64. Das SS, Chaudhuri K, Biswas RN. Successful Convalescent Plasma Therapy in a Child With Severe Coronavirus Disease. *Indian Pediatrics.* 2021;58(1):89-90.
65. De Rienzo M, Foddai ML, Conti L, et al. Long-Term Persistence and Relevant Therapeutic Impact of High-Titer Viral-Neutralizing Antibody in a Convalescent COVID-19 Plasma Super-Donor: A Case Report. *Front Immunol.* 2021;12:690322.
66. De Silvestro G, Marson P, La Raja M, et al. Outcome of SARS CoV-2 inpatients treated with convalescent plasma: One-year of data from the Veneto region (Italy) Registry. *Eur J Intern Med.* 2022;97:42-49.
67. Delgado-Fernandez M, Garcia-Gemar GM, Fuentes-Lopez A, et al. Treatment of COVID-19 with convalescent plasma in patients with humoral immunodeficiency - Three consecutive cases and review of the literature. *Enferm Infecc Microbiol Clin (Engl Ed).* 2021;40(9):507-516.
68. Dell'Isola GB, Felicioni M, Ferraro L, et al. Case Report: Remdesivir and Convalescent Plasma in a Newly Acute B Lymphoblastic Leukemia Diagnosis With Concomitant Sars-CoV-2 Infection. *Front Pediatr.* 2021;9:712603.
69. Destras G, Bal A, Simon B, Lina B, Josset L. Sotrovimab drives SARS-CoV-2 omicron variant evolution in immunocompromised patients. *Lancet Microbe.* 2022;3(8):e559.
70. Deveci B, Saba R. Prolonged viral positivity induced recurrent coronavirus disease 2019 (COVID-19) pneumonia in patients receiving anti-CD20 monoclonal antibody treatment: Case reports. *Medicine (Baltimore).* 2021;100(52):e28470.
71. Di Palma M, Gentilini E, Masucci C, et al. Management of Relapsed/Refractory All with Inotuzumab During COVID-19. A Case Report. *Mediterr J Hematol Infect Dis.* 2022;14(1):e2022043.
72. Diorio C, Anderson EM, McNerney KO, et al. Convalescent plasma for pediatric patients with SARS-CoV-2-associated acute respiratory distress syndrome. *Pediatr Blood Cancer.* 2020;67(11):e28693.
73. Donzelli M, Ippolito M, Catalisano G, et al. Prone positioning and convalescent plasma therapy in a critically ill pregnant woman with COVID-19. *Clin Case Rep.* 2020;8(12):3352-3358.
74. Duan L, Xie Y, Wang Q, et al. Research on antibody changes and nucleic acid clearance in COVID-19 patients treated with convalescent plasma. *Am J Transl Res.* 2022;14(4):2655-2667.
75. Dulipsingh L, Ibrahim D, Schaefer EJ, et al. SARS-CoV-2 serology and virology trends in donors and recipients of convalescent plasma. *Transfus Apher Sci.* 2020;59(6):102922.

 © 2022 Senefeld JW et al.

76. Easterlin MC, De Beritto T, Yeh AM, Wertheimer FB, Ramanathan R. Extremely Preterm Infant Born to a Mother With Severe COVID-19 Pneumonia. *J Investig Med High Impact Case Rep.* 2020;8:2324709620946621.
77. Einollahi B, Cegolon L, Abolghasemi H, et al. A patient affected by critical COVID-19 pneumonia, successfully treated with convalescent plasma. *Transfus Apher Sci.* 2020;59(6):102995.
78. Elliott BP, Buchek GM, Koroscil MT. Characteristics, Treatment, and Outcomes of Patients With Severe or Life-threatening COVID-19 at a Military Treatment Facility-A Descriptive Cohort Study. *Mil Med.* 2022;187(9-10):e1043-e1046.
79. Erber J, Wiessner JR, Huberle C, et al. Convalescent plasma therapy in B-cell-depleted and B-cell sufficient patients with life-threatening COVID-19 - A case series. *Transfus Apher Sci.* 2021;60(6):103278.
80. Erkurt MA, Sarici A, Berber I, Kuku I, Kaya E, Ozgul M. Life-saving effect of convalescent plasma treatment in covid-19 disease: Clinical trial from eastern Anatolia. *Transfus Apher Sci.* 2020;59(5):102867.
81. Faltin K, Lewandowska Z, Malecki P, et al. SARS-CoV-2 attacks the weakest point - COVID-19 course in a pediatric patient with Friedreich's ataxia. *Int J Infect Dis.* 2022;117:284-286.
82. Fanning SL, Korngold R, Yang Z, et al. Elevated cytokines and chemokines in peripheral blood of patients with SARS-CoV-2 pneumonia treated with high-titer convalescent plasma. *PLoS Pathog.* 2021;17(10):e1010025.
83. Fazeli A, Sharifi S, Behdad F, et al. Early high-titer convalescent plasma therapy in patients with moderate and severe COVID-19. *Transfus Apher Sci.* 2022;61(2):103321.
84. Ferrari S, Caprioli C, Weber A, Rambaldi A, Lussana F. Convalescent hyperimmune plasma for chemo-immunotherapy induced immunodeficiency in COVID-19 patients with hematological malignancies. *Leuk Lymphoma.* 2021;62(6):1490-1496.
85. Figlerowicz M, Mania A, Lubarski K, et al. First case of convalescent plasma transfusion in a child with COVID-19-associated severe aplastic anemia. *Transfus Apher Sci.* 2020;59(5):102866.
86. Fisher DL, Pavel A, Malnick S. Rapid recovery of taste and smell in a patient with SARS-CoV-2 following convalescent plasma therapy. *QJM.* 2021;114(5):319-320.
87. Fodor E, Muller V, Ivanyi Z, et al. Early Transfusion of Convalescent Plasma Improves the Clinical Outcome in Severe SARS-CoV2 Infection. *Infect Dis Ther.* 2022;11(1):293-304.
88. Fox H, Gummert JF, Sommer P, Knabbe C, Sohns C. Synergistic effects of levosimendan and convalescence plasma as bailout strategy in acute cardiogenic shock in COVID-19: A case report. *Cardiol J.* 2022;29(1):157-159.
89. Franchini M, Glingani C, Bellani A, et al. Early and persistent viral clearance in COVID-19 patients treated with convalescent plasma. *Transfus Clin Biol.* 2021;28(3):309-310.
90. Franchini M, Glingani C, Donno G, et al. Convalescent Plasma for Hospitalized COVID-19 Patients: A Single-Center Experience. *Life (Basel).* 2022;12(3).
91. Franchini M, Glingani C, Morandi M, et al. Safety and Efficacy of Convalescent Plasma in Elderly COVID-19 Patients: The RESCUE Trial. *Mayo Clin Proc Innov Qual Outcomes.* 2021;5(2):403-412.
92. Fung M, Nambiar A, Pandey S, et al. Treatment of immunocompromised COVID-19 patients with convalescent plasma. *Transpl Infect Dis.* 2021;23(2):e13477.
93. Furlan A, Forner G, Cipriani L, et al. Dramatic Response to Convalescent Hyperimmune Plasma in Association With an Extended Course of Remdesivir in 4 B Cell-Depleted Non-Hodgkin Lymphoma Patients With SARS-Cov-2 Pneumonia After Rituximab Therapy. *Clin Lymphoma Myeloma Leuk.* 2021;21(9):e731-e735.
94. Gachoud D, Pillonel T, Tsilimidos G, et al. Antibody response and intra-host viral evolution after plasma therapy in COVID-19 patients pre-exposed or not to B-cell-depleting agents. *Br J Haematol.* 2022;199(4):549-559.
 © 2022 Senefeld JW et al.

95. Garcia-Vidal C, Iglesias-Caballero M, Puerta-Alcalde P, et al. Emergence of Progressive Mutations in SARS-CoV-2 From a Hematologic Patient With Prolonged Viral Replication. *Front Microbiol.* 2022;13:826883.
96. Gattuso G, Schiavello E, Oltolini C, et al. Prolonged COVID-19 infection in a child with lymphoblastic non-Hodgkin lymphoma: which is the best management? *Tumori.* 2022;108(6):NP1-NP4.
97. Gazitúa R, Briones JL, Selman C, et al. Convalescent Plasma in COVID-19. Mortality-Safety First Results of the Prospective Multicenter FALP 001-2020 Trial. *medRxiv.* 2020:2020.2011.2030.20218560.
98. Gemici A, Bilgen H, Erdogan C, et al. A single center cohort of 40 severe COVID-19 patients who were treated with convalescent plasma. *Turk J Med Sci.* 2020;50(8):1781-1785.
99. Ghadami L, Hasibi M, Asadollahi-Amin A, Asanjarani B, Farahmand M, Abdollahi H. Convalescent plasma therapy in patients with severe COVID-19, A single-arm, retrospective study. *Microb Pathog.* 2022;165:105482.
100. Gharbharan A, GeurtsvanKessel CH, Jordans CCE, et al. Effects of Treatment of Coronavirus Disease 2019 With Convalescent Plasma in 25 B-Cell-Depleted Patients. *Clin Infect Dis.* 2022;74(7):1271-1274.
101. Ghimire P, Pokharel A, Aryal P, Bhandari R, Bhandari B. Clinical Profile of COVID-19 Patients Admitted in a COVID Designated Hospital. *J Nepal Health Res Counc.* 2021;19(3):587-595.
102. Gibson EG, Pender M, Angerbauer M, et al. Prolonged SARS-CoV-2 Illness in a Patient Receiving Ocrelizumab for Multiple Sclerosis. *Open Forum Infect Dis.* 2021;8(7):ofab176.
103. Gibson RL, Sletten ZJ, Sarkisian SA, Sjulin TJ. Treating COVID-19 Acute Severe Hypoxemic Respiratory Failure: First Case Report Utilizing Dexamethasone, Remdesivir, and Convalescent Plasma in Operation Inherent Resolve. *Med J (Ft Sam Houst Tex).* 2021(PB 8-21-01/02/03):28-33.
104. Gomes AMC, Farias GB, Trombetta AC, et al. Phenotype of BTK-lacking myeloid cells during prolonged COVID-19 and upon convalescent plasma. *Eur J Haematol.* 2022.
105. Gonzalez SE, Regairaz L, Ferrando NS, Gonzalez Martinez VV, Salazar MR, Estenssoro E. [Convalescent plasma therapy in COVID-19 patients, in the Province of Buenos Aires]. *Medicina (B Aires).* 2020;80(5):417-424.
106. Gonzalez SE, Regairaz L, Salazar MR, et al. Timing of convalescent plasma administration and 28-day mortality in COVID-19 pneumonia. *J Investig Med.* 2022;70(5):1258-1264.
107. Gordon O, Brosnan MK, Yoon S, et al. Pharmacokinetics of high-titer anti-SARS-CoV-2 human convalescent plasma in high-risk children. *JCI Insight.* 2022;7(2).
108. Greenbaum U, Klein K, Martinez F, et al. High Levels of Common Cold Coronavirus Antibodies in Convalescent Plasma Are Associated With Improved Survival in COVID-19 Patients. *Front Immunol.* 2021;12(1466):675679.
109. Grisolia G, Franchini M, Glingani C, et al. Convalescent plasma for coronavirus disease 2019 in pregnancy: a case report and review. *Am J Obstet Gynecol MFM.* 2020;2(3):100174.
110. Guo F, Deng C, Shi T, Yan Y. Recovery from respiratory failure after 49-day extracorporeal membrane oxygenation support in a critically ill patient with COVID-19: case report. *Eur Heart J Case Rep.* 2021;5(1):ytaa462.
111. Gupta A, Kute VB, Patel HV, et al. Feasibility of Convalescent Plasma Therapy in Kidney Transplant Recipients With Severe COVID-19: A Single-Center Prospective Cohort Study. *Exp Clin Transplant.* 2021;19(4):304-309.
112. Hacibekiroglu T, Kalpakci Y, Genc AC, et al. Efficacy of convalescent plasma according to blood groups in COVID-19 patients. *Turk J Med Sci.* 2021;51(1):45-48.
113. Hahn M, Condori MEH, Totland A, Kristoffersen EK, Hervig TA. [A patient with severe COVID-19 treated with convalescent plasma]. *Tidsskr Nor Laegeforen.* 2020;140(12).

 **© 2022 Senefeld JW et al.**

114. Halfmann PJ, Minor NR, Haddock Iii LA, et al. Evolution of a globally unique SARS-CoV-2 Spike E484T monoclonal antibody escape mutation in a persistently infected, immunocompromised individual. *Virus Evolution.* 2022.
115. Haller ML, Brunner ME, Zender HO. [Intensive care unit treatment for patients suffering from COVID-19: the Neuchatel experience]. *Rev Med Suisse.* 2020;16(716):2284-2286.
116. Hanssen JLJ, Stienstra J, Boers SA, et al. Convalescent Plasma in a Patient with Protracted COVID-19 and Secondary Hypogammaglobulinemia Due to Chronic Lymphocytic Leukemia: Buying Time to Develop Immunity? *Infect Dis Rep.* 2021;13(4):855-864.
117. Hartman W, Hess A, Connor J. Use of COVID-19 convalescent plasma as prophylaxis in a patient with new onset ALL. *Clin Oncol Case Reports.* 2020;4(1).
118. Hartman WR, Hess AS, Connor JP. Hospitalized COVID-19 patients treated with convalescent plasma in a mid-size city in the Midwest. *Transl Med Commun.* 2020;5(1):17.
119. Hartman WR, Hess AS, Connor JP. Unusual Cardiac Presentation of COVID-19 and Use of Convalescent Plasma. *Case Rep Cardiol.* 2020;2020:8863195.
120. He R, Wang J, Wang Z, et al. A possible dose-response equation: Viral load after plasma infusion in COVID-19 patients and anti-SARS-CoV-2 antibody titers in convalescent plasma. *Transfus Med.* 2022;32(2):162-167.
121. Helleberg M, Niemann CU, Moestrup KS, Kirk O, Lebech AM, Lundgren J. Response to Aviv et al. *J Infect Dis.* 2021;224(3):559-561.
122. Hemsinlioglu C, Pelit NB, Yalcin K, et al. THE EFFECTIVENESS OF ACB-IP 1.0 UNIVERSAL PATHOGEN FREE CONCENTRATED COCKTAIL CONVALESCENT PLASMA IN COVID-19 INFECTION. *medRxiv.* 2021:2021.2003.2005.21251413.
123. Herman JD, Wang C, Loos C, et al. Functional convalescent plasma antibodies and pre-infusion titers shape the early severe COVID-19 immune response. *Nat Commun.* 2021;12(1):6853.
124. Honjo K, Russell RM, Li R, et al. Convalescent plasma-mediated resolution of COVID-19 in a patient with humoral immunodeficiency. *Cell Rep Med.* 2021;2(1):100164.
125. Hopwood AJ, Jordan-Villegas A, Gutierrez LD, et al. Severe Acute Respiratory Syndrome Coronavirus-2 Pneumonia in a Newborn Treated With Remdesivir and Coronavirus Disease 2019 Convalescent Plasma. *J Pediatric Infect Dis Soc.* 2021;10(5):691-694.
126. Hovey JG, Tolbert D, Howell D. Burton's Agammaglobulinemia and COVID-19. *Cureus.* 2020;12(11):e11701.
127. Hu R, Qiu W, Zhao X, et al. [Management of immunocompromised renal transplant patients infected with coronavirus disease 2019]. *Zhonghua Wei Zhong Bing Ji Jiu Yi Xue.* 2022;34(5):492-496.
128. Hu X, Hu C, Jiang D, et al. Effectiveness of Convalescent Plasma Therapy for COVID-19 Patients in Hunan, China. *Dose Response.* 2020;18(4):1559325820979921.
129. Huang J, Weng H, Lan C, Li H. Convalescent plasma therapy for patients with severe COVID-19: A case series study. *Medicine (Baltimore).* 2022;101(31):e29912.
130. Huang S, Shen C, Xia C, Huang X, Fu Y, Tian L. A Retrospective Study on the Effects of Convalescent Plasma Therapy in 24 Patients Diagnosed with COVID-19 Pneumonia in February and March 2020 at 2 Centers in Wuhan, China. *Med Sci Monit.* 2020;26:e928755.
131. Hueso T, Godron AS, Lanoy E, et al. Convalescent plasma improves overall survival in patients with B-cell lymphoid malignancy and COVID-19: a longitudinal cohort and propensity score analysis. *Leukemia.* 2022;36(4):1025-1034.
132. Hueso T, Pouderoux C, Pere H, et al. Convalescent plasma therapy for B-cell-depleted patients with protracted COVID-19. *Blood.* 2020;136(20):2290-2295.
133. Hughes CM, Gregory GP, Pierce AB, et al. Clinical illness with viable severe acute respiratory coronavirus virus 2 (SARS-CoV-2) virus presenting 72 days after infection in an immunocompromised patient. *Infect Control Hosp Epidemiol.* 2022;43(6):820-822.

 © 2022 Senefeld JW et al.

134. Iaboni A, Wong N, Betschel SD. A Patient with X-Linked Agammaglobulinemia and COVID-19 Infection Treated with Remdesivir and Convalescent Plasma. *J Clin Immunol.* 2021;41(5):923-925.
135. Ibrahim D, Dulipsingh L, Zapatka L, et al. Factors Associated with Good Patient Outcomes Following Convalescent Plasma in COVID-19: A Prospective Phase II Clinical Trial. *Infect Dis Ther.* 2020;9(4):913-926.
136. Iglesias JI, Vassallo AV, Sullivan JB, et al. Retrospective analysis of anti-inflammatory therapies during the first wave of COVID-19 at a community hospital. *World J Crit Care Med.* 2021;10(5):244-259.
137. Im JH, Nahm CH, Baek JH, Kwon HY, Lee JS. Convalescent Plasma Therapy in Coronavirus Disease 2019: a Case Report and Suggestions to Overcome Obstacles. *J Korean Med Sci.* 2020;35(26):e239.
138. Ipe TS, Ugwumba B, Spencer HJ, et al. Treatment of COVID-19 Patients with Two Units of Convalescent Plasma in a Resource-Constrained State. *Lab Med.* 2022;53(6):623-628.
139. Jacobs JP, Stammers AH, St Louis JD, et al. Multi-institutional Analysis of 200 COVID-19 Patients Treated With Extracorporeal Membrane Oxygenation: Outcomes and Trends. *Ann Thorac Surg.* 2022;113(5):1452-1460.
140. Jacobson J, Antony K, Beninati M, Alward W, Hoppe KK. Use of dexamethasone, remdesivir, convalescent plasma and prone positioning in the treatment of severe COVID-19 infection in pregnancy: A case report. *Case Rep Womens Health.* 2021;29:e00273.
141. Jafari R, Jonaidi-Jafari N, Dehghanpoor F, Saburi A. Convalescent plasma therapy in a pregnant COVID-19 patient with a dramatic clinical and imaging response: A case report. *World J Radiol.* 2020;12(7):137-141.
142. Jaiswal V, Nasa P, Raouf M, et al. Therapeutic plasma exchange followed by convalescent plasma transfusion in critical COVID-19-An exploratory study. *Int J Infect Dis.* 2021;102:332-334.
143. Jamir I, Lohia P, Pande RK, Setia R, Singhal AK, Chaudhary A. Convalescent plasma therapy and remdesivir duo successfully salvaged an early liver transplant recipient with severe COVID-19 pneumonia. *Ann Hepatobiliary Pancreat Surg.* 2020;24(4):526-532.
144. Jamous F, Meyer N, Buus D, et al. Critical Illness Due to Covid-19: A Description of the Surge in a Single Center in Sioux Falls. *S D Med.* 2020;73(7):312-317.
145. Janaka SK, Hartman W, Mou H, et al. Donor Anti-Spike Immunity is Related to Recipient Recovery and Can Predict the Efficacy of Convalescent Plasma Units. *medRxiv.* 2021:2021.2002.2025.21252463.
146. Jassem J, Marek-Trzonkowska NM, Smiatacz T, et al. Successful Treatment of Persistent SARS-CoV-2 Infection in a B-Cell Depleted Patient with Activated Cytotoxic T and NK Cells: A Case Report. *Int J Mol Sci.* 2021;22(20).
147. Jasuja S, Sagar G, Bahl A, Verma S. COVID-19 Infection Clinical Profile, Management, Outcome, and Antibody Response in Kidney Transplant Recipients: A Single Centre Experience. *Int J Nephrol.* 2021;2021:3129411.
148. Jeyaraman P, Agrawal N, Bhargava R, et al. Convalescent plasma therapy for severe Covid-19 in patients with hematological malignancies. *Transfus Apher Sci.* 2021;60(3):103075.
149. Ji F, Liu W, Hao DA, et al. Use of convalescent plasma therapy in eight individuals with mild COVID-19. *New Microbes New Infect.* 2021;39:100814.
150. Jiang J, Miao Y, Zhao Y, et al. Convalescent plasma therapy: Helpful treatment of COVID-19 in a kidney transplant recipient presenting with severe clinical manifestations and complex complications. *Clin Transplant.* 2020;34(9):e14025.
151. Jin C, Gu J, Yuan Y, et al. Treatment of Six COVID-19 Patients with Convalescent Plasma. *medRxiv.* 2020:2020.2005.2021.20109512.

 **© 2022 Senefeld JW et al.**

152. Jin H, Reed JC, Liu STH, et al. Three patients with X-linked agammaglobulinemia hospitalized for COVID-19 improved with convalescent plasma. *J Allergy Clin Immunol Pract.* 2020;8(10):3594-3596 e3593.
153. Joyner MJ, Carter RE, Senefeld JW, et al. Convalescent Plasma Antibody Levels and the Risk of Death from Covid-19. *N Engl J Med.* 2021;384(11):1015-1027.
154. Karaolidou F, Loutsidi NE, Mellios Z, et al. Convalescent plasma therapy in an immunocompromised patient with multiple COVID-19 flares: a case report. *Respirol Case Rep.* 2021;9(12):e0858.
155. Karatas A, Inkaya AC, Demiroglu H, et al. Prolonged viral shedding in a lymphoma patient with COVID-19 infection receiving convalescent plasma. *Transfus Apher Sci.* 2020;59(5):102871.
156. Katz-Greenberg G, Yadav A, Gupta M, et al. Outcomes of COVID-19-positive kidney transplant recipients: A single-center experience. *Clin Nephrol.* 2020;94(6):318-321.
157. Keitel V, Bode JG, Feldt T, et al. Case Report: Convalescent Plasma Achieves SARS-CoV-2 Viral Clearance in a Patient With Persistently High Viral Replication Over 8 Weeks Due to Severe Combined Immunodeficiency (SCID) and Graft Failure. *Front Immunol.* 2021;12:645989.
158. Kemp SA, Collier DA, Datir RP, et al. SARS-CoV-2 evolution during treatment of chronic infection. *Nature.* 2021;592(7853):277-282.
159. Kenig A, Ishay Y, Kharouf F, Rubin L. Treatment of B-cell depleted COVID-19 patients with convalescent plasma and plasma-based products. *Clin Immunol.* 2021;227:108723.
160. Ketels T, Gisolf J, Claassen M, et al. Short Communication: Prolonged COVID-19 Infection in a Patient with Newly Diagnosed HIV/AIDS. *AIDS Res Hum Retroviruses.* 2022;38(5):399-400.
161. Khamis F, Memish Z, Al Bahrani M, et al. The Role of Convalescent Plasma and Tocilizumab in the Management of COVID-19 Infection: A Cohort of 110 Patients from a Tertiary Care Hospital in Oman. *J Epidemiol Glob Health.* 2021;11(2):216-223.
162. Khan AM, Ajmal Z, Raval M, Tobin E. Concurrent Diagnosis of Acute Myeloid Leukemia and COVID-19: A Management Challenge. *Cureus.* 2020;12(8):e9629.
163. Khan TNS, Mukry SN, Masood S, et al. Usefulness of convalescent plasma transfusion for the treatment of severely ill COVID-19 patients in Pakistan. *BMC Infect Dis.* 2021;21(1):1014.
164. Khatamzas E, Antwerpen MH, Rehn A, et al. Accumulation of mutations in antibody and CD8 T cell epitopes in a B cell depleted lymphoma patient with chronic SARS-CoV-2 infection. *Nat Commun.* 2022;13(1):5586.
165. Khatri A, Chang KM, Berlinrut I, Wallach F. Mucormycosis after Coronavirus disease 2019 infection in a heart transplant recipient - Case report and review of literature. *J Mycol Med.* 2021;31(2):101125.
166. Khot R, Kumbhalkar SD, Juneja R, Joshi PP. The Development of Ecchymosis after Administration of Convalescent Serum in a Patient with COVID-19 Associated with Thrombocytosis. *Eur J Case Rep Intern Med.* 2021;8(9):002782.
167. King CS, Sahjwani D, Brown AW, et al. Outcomes of mechanically ventilated patients with COVID-19 associated respiratory failure. *PLoS One.* 2020;15(11):e0242651.
168. Kluger MA, Czogalla J, Schmidt-Lauber C, et al. Convalescent plasma treatment for early post-kidney transplant acquired COVID-19. *Transpl Infect Dis.* 2021;23(4):e13685.
169. Ko JJ, Wu C, Mehta N, Wald-Dickler N, Yang W, Qiao R. A Comparison of Methylprednisolone and Dexamethasone in Intensive Care Patients With COVID-19. *J Intensive Care Med.* 2021;36(6):673-680.
170. Kocayigit H, Demir G, Karacan A, et al. Effects on mortality of early vs late administration of convalescent plasma in the treatment of Covid-19. *Transfus Apher Sci.* 2021;60(4):103148.
171. Kochanek M, Garcia Borrega J, Beckmann L, et al. Plasma exchange with COVID-19 convalescent plasma in a patient with severe ANCA-associated vasculitis and COVID-19 pneumonia after rituximab therapy. *Clin Kidney J.* 2022;15(1):162-164.

 © 2022 Senefeld JW et al.

172. Kong Y, Cai C, Ling L, et al. Successful treatment of a centenarian with coronavirus disease 2019 (COVID-19) using convalescent plasma. *Transfus Apher Sci.* 2020;59(5):102820.
173. Kremer AE, Kremer AN, Willam C, et al. Successful treatment of COVID-19 infection with convalescent plasma in B-cell-depleted patients may promote cellular immunity. *Eur J Immunol.* 2021;51(10):2478-2484.
174. Lancman G, Mascarenhas J, Bar-Natan M. Severe COVID-19 virus reactivation following treatment for B cell acute lymphoblastic leukemia. *J Hematol Oncol.* 2020;13(1):131.
175. Lang-Meli J, Fuchs J, Mathe P, et al. Case Series: Convalescent Plasma Therapy for Patients with COVID-19 and Primary Antibody Deficiency. *J Clin Immunol.* 2022;42(2):253-265.
176. Lattanzio N, Acosta-Diaz C, Villasmil RJ, et al. Effectiveness of COVID-19 Convalescent Plasma Infusion Within 48 Hours of Hospitalization With SARS-CoV-2 Infection. *Cureus.* 2021;13(7):e16746.
177. Lazzari L, Oltolini C, Ciceri F, Giglio F. Complicated and persistent severe COVID-19 pneumonia in a recipient of allogeneic haematopoietic stem cell transplant. *BMJ Case Rep.* 2021;14(10).
178. Lee KT, Yeoh WC, Zainul NH, Syed Alwi SB, Low LL. Convalescent plasma as an adjunctive therapy for COVID-19: A single centre experience in Malaysia. *Med J Malaysia.* 2021;76(5):653-657.
179. Lemus HN, Alkayyali M, Kim E, Cunnigham-Rundles C, Pyburn D, Abrams R. Acute Cerebellitis and Myeloradiculitis Associated With SARS-CoV-2 Infection in Common Variable Immunodeficiency-A Case Report. *Neurohospitalist.* 2022;12(2):361-365.
180. Leon J, Merrill AE, Rogers K, et al. SARS-CoV-2 antibody changes in patients receiving COVID-19 convalescent plasma from normal and vaccinated donors. *Transfus Apher Sci.* 2022;61(2):103326.
181. Levy I, Lavi A, Zimran E, et al. COVID-19 among patients with hematological malignancies: a national Israeli retrospective analysis with special emphasis on treatment and outcome. *Leuk Lymphoma.* 2021;62(14):3384-3393.
182. Lima B, Gibson GT, Vullaganti S, et al. COVID-19 in recent heart transplant recipients: Clinicopathologic features and early outcomes. *Transpl Infect Dis.* 2020;22(5):e13382.
183. Lindemann M, Krawczyk A, Dolff S, et al. SARS-CoV-2-specific humoral and cellular immunity in two renal transplants and two hemodialysis patients treated with convalescent plasma. *J Med Virol.* 2021;93(5):3047-3054.
184. Lindemann M, Lenz V, Knop D, et al. Convalescent plasma treatment of critically ill intensive care COVID-19 patients. *Transfusion.* 2021;61(5):1394-1403.
185. Liu B, Ren KK, Wang N, Xu XP, Wu J. Timing of convalescent plasma therapy-tips from curing a 100-year-old COVID-19 patient using convalescent plasma treatment: A case report. *World J Clin Cases.* 2021;9(12):2890-2898.
186. Liu M, Chen Z, Dai MY, et al. Lessons learned from early compassionate use of convalescent plasma on critically ill patients with Covid-19. *Transfusion.* 2020;60(10):2210-2216.
187. Liu Y, Xie W, Li H, et al. [Critical coronavirus disease 2019 caused by Delta variant: a case report with literature review]. *Zhonghua Wei Zhong Bing Ji Jiu Yi Xue.* 2022;34(5):481-484.
188. Ljungquist O, Lundgren M, Iliachenko E, et al. Convalescent plasma treatment in severely immunosuppressed patients hospitalized with COVID-19: an observational study of 28 cases. *Infect Dis (Lond).* 2022;54(4):283-291.
189. London J, Boutboul D, Lacombe K, et al. Severe COVID-19 in Patients with B Cell Alymphocytosis and Response to Convalescent Plasma Therapy. *J Clin Immunol.* 2021;41(2):356-361.
190. Lopez V, Vazquez-Sanchez T, Casas C, et al. Risk Factors for Mortality in Stable Kidney Transplant Patients Infected by SARS-CoV-2 in the South of Spain. *Transplant Proc.* 2021;53(9):2685-2687.

 **© 2022 Senefeld JW et al.**

191. Lubnow M, Schmidt B, Fleck M, et al. Secondary hemophagocytic lymphohistiocytosis and severe liver injury induced by hepatic SARS-CoV-2 infection unmasking Wilson's disease: Balancing immunosuppression. *Int J Infect Dis.* 2021;103:624-627.
192. Luetkens T, Metcalf R, Planelles V, et al. Successful transfer of anti-SARS-CoV-2 immunity using convalescent plasma in an MM patient with hypogammaglobulinemia and COVID-19. *Blood Adv.* 2020;4(19):4864-4868.
193. Madariaga MLL, Guthmiller JJ, Schrantz S, et al. Clinical predictors of donor antibody titre and correlation with recipient antibody response in a COVID-19 convalescent plasma clinical trial. *J Intern Med.* 2021;289(4):559-573.
194. Magallanes-Garza GI, Valdez-Alatorre C, Davila-Gonzalez D, et al. Rapid improvement of a critically ill obstetric patient with SARS-CoV-2 infection after administration of convalescent plasma. *Int J Gynaecol Obstet.* 2021;152(3):439-441.
195. Magyari F, Pinczes LI, Payer E, et al. Early administration of remdesivir plus convalescent plasma therapy is effective to treat COVID-19 pneumonia in B-cell depleted patients with hematological malignancies. *Ann Hematol.* 2022;101(10):2337-2345.
196. Malecki P, Faltin K, Mania A, et al. Effects and Safety of Convalescent Plasma Administration in a Group of Polish Pediatric Patients with COVID-19: A Case Series. *Life (Basel).* 2021;11(3).
197. Malsy J, Veletzky L, Heide J, et al. Sustained Response After Remdesivir and Convalescent Plasma Therapy in a B-Cell-Depleted Patient With Protracted Coronavirus Disease 2019 (COVID-19). *Clin Infect Dis.* 2021;73(11):e4020-e4024.
198. Maor Y, Cohen D, Paran N, et al. Compassionate use of convalescent plasma for treatment of moderate and severe pneumonia in COVID-19 patients and association with IgG antibody levels in donated plasma. *EClinicalMedicine.* 2020;26:100525.
199. Marconato M, Abela IA, Hauser A, et al. Antibodies from convalescent plasma promote SARS-CoV-2 clearance in individuals with and without endogenous antibody response. *J Clin Invest.* 2022;132(12).
200. Martens T, Hens L, De Pauw M, Van Belleghem Y. Heart Transplantation Complicated by COVID-19 Infection. *Ann Thorac Surg.* 2022;113(4):e267-e269.
201. Martinez-Barranco P, Garcia-Roa M, Trelles-Martinez R, et al. Management of Persistent SARS-CoV-2 Infection in Patients with Follicular Lymphoma. *Acta Haematol.* 2022;145(4):384-393.
202. Martinez-Chinchilla C, Vazquez-Montero L, Palazon-Carrion N, et al. Persistence of SARS-CoV-2 Infection in Severely Immunocompromised Patients With Complete Remission B-Cell Lymphoma and Anti-CD20 Monoclonal Antibody Therapy: A Case Report of Two Cases. *Front Immunol.* 2022;13:860891.
203. Martinez-Resendez MF, Castilleja-Leal F, Torres-Quintanilla A, et al. Initial experience in Mexico with convalescent plasma in COVID-19 patients with severe respiratory failure, a retrospective case series. *medRxiv.* 2020:2020.2007.2014.20144469.
204. Martinot M, Jary A, Fafi-Kremer S, et al. Emerging RNA-Dependent RNA Polymerase Mutation in a Remdesivir-Treated B-cell Immunodeficient Patient With Protracted Coronavirus Disease 2019. *Clin Infect Dis.* 2021;73(7):e1762-e1765.
205. Marwah V, Choudhary R, Peter D, Bhati G. Pulmonary thromboembolism post-COVID convalescent plasma therapy: adding fuel to a smoldering fire! *Adv Respir Med.* 2021;89(3):347-349.
206. Mastroianni A, Greco S, Mauro MV, Chidichimo L, Vangeli V. Convalescent plasma transfusion for pregnant patients with COVID-19. *Lancet Microbe.* 2021;2(8):e350.
207. McCuddy M, Kelkar P, Zhao Y, Wicklund D. Acute Demyelinating Encephalomyelitis (ADEM) in COVID-19 Infection: A Case Series. *Neurol India.* 2020;68(5):1192-1195.
208. McKemey E, Shields AM, Faustini SE, et al. Resolution of Persistent COVID-19 After Convalescent Plasma in a Patient with B Cell Aplasia. *J Clin Immunol.* 2021;41(5):926-929.
 **© 2022 Senefeld JW et al.**

209. Mehta SA, Rana MM, Motter JD, et al. Incidence and Outcomes of COVID-19 in Kidney and Liver Transplant Recipients With HIV: Report From the National HOPE in Action Consortium. *Transplantation.* 2021;105(1):216-224.
210. Mekonnen ZK, Ashraf DC, Jankowski T, et al. Acute Invasive Rhino-Orbital Mucormycosis in a Patient With COVID-19-Associated Acute Respiratory Distress Syndrome. *Ophthalmic Plast Reconstr Surg.* 2021;37(2):e40-e80.
211. Mendes-Correa MC, Ghilardi F, Salomão MC, et al. SARS-CoV-2 shedding, infectivity and evolution in an immunocompromised adult patient. *medRxiv.* 2021:2021.2006.2011.21257717.
212. Merin NM, LeVee AA, Merlo CA, et al. The feasibility of multiple units of convalescent plasma in mechanically ventilated patients with COVID-19: A pilot study. *Transfus Apher Sci.* 2022;61(4):103423.
213. Meshram HS, Kute VB, Patel H, Banerjee S, Desai S, Chauhan S. Cytomegalovirus and Severe Acute Respiratory Syndrome Coronavirus 2 Co-infection in Renal Transplants: A Retrospective Study from a Single Center. *Saudi J Kidney Dis Transpl.* 2021;32(4):929-938.
214. Milosevic I, Jovanovic J, Stevanovic O. Atypical course of COVID-19 in patient with Bruton agammaglobulinemia. *J Infect Dev Ctries.* 2020;14(11):1248-1251.
215. Milota T, Sobotkova M, Smetanova J, et al. Risk Factors for Severe COVID-19 and Hospital Admission in Patients With Inborn Errors of Immunity - Results From a Multicenter Nationwide Study. *Front Immunol.* 2022;13:835770.
216. Mira E, Yarce OA, Ortega C, et al. Rapid recovery of a SARS-CoV-2-infected X-linked agammaglobulinemia patient after infusion of COVID-19 convalescent plasma. *J Allergy Clin Immunol Pract.* 2020;8(8):2793-2795.
217. Mirihagalle N, Parajuli P, Sundareshan V, et al. Sequential dosing of convalescent COVID-19 plasma with significant temporal clinical improvements in a persistently SARS-COV-2 positive patient. *Transfus Apher Sci.* 2021;60(5):103180.
218. Mohanty B, Sunder A, Satyanarayan B, Kumar M, Shukla R, Ahmed A. Success rate of Remdesivir, Convalescent Plasma, and Tocilizumab in moderate to severe Covid-19 pneumonia: our experience in a tertiary care center. *J Family Med Prim Care.* 2021;10(11):4236-4241.
219. Mohseni M, Albus M, Kaminski A, Harrison MF. A Case of COVID-19 Re-Infection in a Liver Transplant Patient. *Cureus.* 2021;13(5):e14916.
220. Moniuszko-Malinowska A, Czupryna P, Boczkowska-Radziwon B, et al. A 63-Year-Old Woman with SARS-CoV-2 Infection, Who Developed Severe COVID-19 Pneumonia and Was Supported with Convalescent Plasma Therapy. *Am J Case Rep.* 2020;21:e927662.
221. Monrad I, Sahlertz SR, Nielsen SSF, et al. Persistent Severe Acute Respiratory Syndrome Coronavirus 2 Infection in Immunocompromised Host Displaying Treatment Induced Viral Evolution. *Open Forum Infect Dis.* 2021;8(7):ofab295.
222. Moore JL, Ganapathiraju PV, Kurtz CP, Wainscoat B. A 63-Year-Old Woman with a History of Non-Hodgkin Lymphoma with Persistent SARS-CoV-2 Infection Who Was Seronegative and Treated with Convalescent Plasma. *Am J Case Rep.* 2020;21:e927812.
223. Moura LW, Liao AW, Negrini R, Zlotnik E. The use of convalescent plasma therapy in the management of a pregnant woman with COVID-19: a case report. *Einstein (Sao Paulo).* 2022;20:eRC6550.
224. Moutinho-Pereira S, Calisto R, Sabio F, Guerreiro L. High-titre convalescent plasma therapy for an immunocompromised patient with systemic lupus erythematosus with protracted SARS-CoV-2 infection. *BMJ Case Rep.* 2021;14(8).
225. Mukhina OA, Fomina DS, Parshin VV, et al. SARS-CoV-2 evolution in a patient with secondary B-cell immunodeficiency: A clinical case. *Hum Vaccin Immunother.* 2022;18(6):2101334.
226. Mullaguri N, Sivakumar S, Battineni A, Anand S, Vanderwerf J. COVID-19 Related Acute Hemorrhagic Necrotizing Encephalitis: A Report of Two Cases and Literature Review. *Cureus.* 2021;13(4):e14236.
 © 2022 Senefeld JW et al.

227. Naeem S, Gohh R, Bayliss G, et al. Successful recovery from COVID-19 in three kidney transplant recipients who received convalescent plasma therapy. *Transpl Infect Dis.* 2021;23(1):e13451.
228. Negi G, Kaur D, Jain A, et al. Convalescent Plasma The Experience And Journey From Blockbuster To Incognito: A Single Centre Experience. *Recent Adv Antiinfect Drug Discov.* 2022.
229. Ng TG, Degaetano E, Trivedi U, Akthar M. Barotrauma Linked to Coronavirus Disease 2019 Infection in Younger Patients: A Case Series. *Cureus.* 2021;13(4):e14573.
230. Nguyen MC, Lee EJ, Avery RK, et al. Transplant of SARS-CoV-2-infected Living Donor Liver: Case Report. *Transplant Direct.* 2021;7(8):e721.
231. Niu A, McDougal A, Ning B, et al. COVID-19 in allogeneic stem cell transplant: high false-negative probability and role of CRISPR and convalescent plasma. *Bone Marrow Transplant.* 2020;55(12):2354-2356.
232. Novacescu AN, Duma G, Sandesc D, Sorescu T, Licker M. The Successful Recovery of a Critically Ill COVID-19 Patient, Following the Combination of Therapeutic Plasma Exchange and Convalescent Plasma Transfusion: A Case Report. *Medicina (Kaunas).* 2022;58(8).
233. Nowak PJ, Forycka J, Cegielska N, et al. Glucocorticoids Induce Partial Remission of Focal Segmental Glomerulosclerosis but Not Interstitial Nephritis in COVID-19 Acute Kidney Injury in an APOL1 Low-Risk Genotype White Patient. *Am J Case Rep.* 2021;22:e933462.
234. Nussenblatt V, Roder AE, Das S, et al. Yearlong COVID-19 Infection Reveals Within-Host Evolution of SARS-CoV-2 in a Patient With B-Cell Depletion. *J Infect Dis.* 2022;225(7):1118-1123.
235. Nystrom K, Hjorth M, Fust R, et al. Specific T-cell responses for guiding treatment with convalescent plasma in severe COVID-19 and humoral immunodeficiency: a case report. *BMC Infect Dis.* 2022;22(1):362.
236. Oehler D, Bruno RR, Holst HT, et al. [COVID-19 after heart transplantation: experiences from a German transplantation center]. *Z Herz Thorax Gefasschir.* 2022;36(6):406-413.
237. Oliva A, Cancelli F, Brogi A, et al. Convalescent plasma for haematological patients with SARS-CoV-2 pneumonia and severe depletion of B-cell lymphocytes following anti-CD20 therapy: a single-centre experience and review of the literature. *New Microbiol.* 2022;45(1):62-72.
238. Olivares F, Munoz D, Fica A, et al. Clinical features of 47 patients infected with COVID-19 admitted to a Regional Reference Center. *Rev Med Chil.* 2020;148(11):1577-1588.
239. Olivares-Gazca JC, Priesca-Marin JM, Ojeda-Laguna M, et al. Infusion of Convalescent Plasma Is Associated with Clinical Improvement in Critically Ill Patients with Covid-19: A Pilot Study. *Rev Invest Clin.* 2020;72(3):159-164.
240. Oliveira E, Parikh A, Lopez-Ruiz A, et al. ICU outcomes and survival in patients with severe COVID-19 in the largest health care system in central Florida. *PLoS One.* 2021;16(3):e0249038.
241. Ordaya EE, Abu Saleh OM, Stubbs JR, Joyner MJ. Vax-Plasma in Patients With Refractory COVID-19. *Mayo Clin Proc.* 2022;97(1):186-189.
242. Ormazabal Velez I, Indurain Bermejo J, Espinoza Perez J, Imaz Aguayo L, Delgado Ruiz M, Garcia-Erce JA. Two patients with rituximab associated low gammaglobulin levels and relapsed covid-19 infections treated with convalescent plasma. *Transfus Apher Sci.* 2021;60(3):103104.
243. Padilla R, Arquiette J, Mai Y, Singh G, Galang K, Liang E. Clinical Outcomes of COVID-19 Patients Treated with Convalescent Plasma or Remdesivir Alone and in Combination at a Community Hospital in California's Central Valley. *J Pharm Pharm Sci.* 2021;24:210-219.
244. Pagan ME, Ramseyer AM, Whitcombe DD, et al. Management of Critically Ill Pregnant Patients with COVID-19 Infection in a Rural State. *Am J Perinatol.* 2022;39(2):165-171.

 © 2022 Senefeld JW et al.

245. Pal P, Ibrahim M, Niu A, et al. Safety and efficacy of COVID-19 convalescent plasma in severe pulmonary disease: A report of 17 patients. *Transfus Med.* 2021;31(3):217-220.
246. Pei S, Yuan X, Zhang Z, et al. Convalescent plasma to treat COVID-19: clinical experience and efficacy. *Aging (Albany NY).* 2021;13(6):7758-7766.
247. Pelayo J, Pugliese G, Salacup G, et al. Severe COVID-19 in Third Trimester Pregnancy: Multidisciplinary Approach. *Case Rep Crit Care.* 2020;2020:8889487.
248. Peng H, Gong T, Huang X, et al. A synergistic role of convalescent plasma and mesenchymal stem cells in the treatment of severely ill COVID-19 patients: a clinical case report. *Stem Cell Res Ther.* 2020;11(1):291.
249. Peppers BP, Shmookler A, Stanley J, et al. Serial Convalescent Plasma Infusions for the Initial COVID-19 Infections in the Appalachian Region of West Virginia. *Allergy Rhinol (Providence).* 2022;13:21526575221110488.
250. Plotnikov G, Waizman E, Tzur I, Yusupov A, Shapira Y, Gorelik O. The prognostic role of functional dependency in older inpatients with COVID-19. *BMC Geriatr.* 2021;21(1):219.
251. Pommeret F, Colomba J, Bigenwald C, et al. Bamlanivimab + etesevimab therapy induces SARS-CoV-2 immune escape mutations and secondary clinical deterioration in COVID-19 patients with B-cell malignancies. *Ann Oncol.* 2021;32(11):1445-1447.
252. Prasad RM, Srivastava S, Wang E, et al. Effect of Immunosuppressive Diseases and Rituximab Infusions on Allowing COVID-19 Infection to Relapse. *Perm J.* 2021;26(1):123-131.
253. Radhakrishnan Nair D, Coggeshall JW, Scalise ML. A case report on a severely ill COVID-19 patient with rapid clearing of chest infiltrates 43 hours after treatment with multiple modalities. *J Community Hosp Intern Med Perspect.* 2021;11(1):33-35.
254. Ragab D, Salah-Eldin H, Afify M, Soliman W, Badr MH. A case of COVID-19, with cytokine storm, treated by consecutive use of therapeutic plasma exchange followed by convalescent plasma transfusion: A case report. *J Med Virol.* 2021;93(4):1854-1856.
255. Rahardjo TM, Yogipranata E, Naswan AH, et al. Effectiveness of convalescent plasma therapy in eight non-intubated coronavirus disease 2019 patients in Indonesia: a case series. *J Med Case Rep.* 2021;15(1):564.
256. Rahimi-Levene N, Shapira J, Tzur I, et al. Predictors of mortality in COVID-19 patients treated with convalescent plasma therapy. *PLoS One.* 2022;17(7):e0271036.
257. Rahman F, Liu STH, Taimur S, et al. Treatment with convalescent plasma in solid organ transplant recipients with COVID-19: Experience at large transplant center in New York City. *Clin Transplant.* 2020;34(12):e14089.
258. Ramalingam S, Arora H, Gunasekaran K, Muruganandam M, Nagaraju S. A Unique Case of Spontaneous Pneumomediastinum in a Patient With COVID-19 and Influenza Coinfection. *J Investig Med High Impact Case Rep.* 2021;9:23247096211016228.
259. Rejeki MS, Sarnadi N, Wihastuti R, et al. Convalescent plasma therapy in patients with moderate-to-severe COVID-19: A study from Indonesia for clinical research in low- and middle-income countries. *EClinicalMedicine.* 2021;36:100931.
260. Ribeiro LC, Benites BD, Ulaf RG, et al. Rapid clinical recovery of a SARS-CoV-2 infected common variable immunodeficiency patient following the infusion of COVID-19 convalescent plasma. *Allergy Asthma Clin Immunol.* 2021;17(1):14.
261. Ripoll JG, Gorman EK, Juskewitch JE, et al. Vaccine-boosted convalescent plasma therapy for patients with immunosuppression and COVID-19. *Blood Adv.* 2022;6(23):5951-5955.
262. Rizvi S, Danic M, Silver M, LaBond V. Cytosorb filter: An adjunct for survival in the COVID-19 patient in cytokine storm? a case report. *Heart Lung.* 2021;50(1):44-50.
263. Rnjak D, Ravlic S, Sola AM, et al. COVID-19 convalescent plasma as long-term therapy in immunodeficient patients? *Transfus Clin Biol.* 2021;28(3):264-270.
264. Rodriguez JA, Bonnano C, Khatiwada P, Roa AA, Mayer D, Eckardt PA. COVID-19 Coinfection with Mycobacterium abscessus in a Patient with Multiple Myeloma. *Case Rep Infect Dis.* 2021;2021:8840536.

 © 2022 Senefeld JW et al.

265. Rodriguez Z, Shane AL, Verkerke H, et al. COVID-19 convalescent plasma clears SARS-CoV-2 refractory to remdesivir in an infant with congenital heart disease. *Blood Adv.* 2020;4(18):4278-4281.
266. Rodriguez-Pla A, Vikram HR, Khalid V, Wesselius LJ. COVID-19 pneumonia in a patient with granulomatosis with polyangiitis on rituximab: case-based review. *Rheumatol Int.* 2021;41(8):1509-1514.
267. Roshandel E, Sankanian G, Salimi M, et al. Plasma exchange followed by convalescent plasma transfusion in COVID-19 patients. *Transfus Apher Sci.* 2021;60(4):103141.
268. Rosner-Tenerowicz A, Fuchs T, Zimmer-Stelmach A, et al. Placental pathology in a pregnant woman with severe COVID-19 and successful ECMO treatment: a case report. *BMC Pregnancy Childbirth.* 2021;21(1):760.
269. Rufenacht S, Gantenbein P, Boggian K, et al. Remdesivir in Coronavirus Disease 2019 patients treated with anti-CD20 monoclonal antibodies: a case series. *Infection.* 2022;50(3):783-790.
270. Saad Shaukat MH, Petrasko P, Petrasko M, Rynders B, Pham S. Index Case of COVID-19 Myocarditis Requiring BiV-ICD for Primary Prevention of Sudden Cardiac Death. *S D Med.* 2021;74(8):380-383.
271. Saha A, Ahsan MM, Quader TU, et al. Characteristics, management and outcomes of critically ill COVID-19 patients admitted to ICU in hospitals in Bangladesh: a retrospective study. *J Prev Med Hyg.* 2021;62(1):E33-E45.
272. Sait AS, Chiang TP, Marr KA, et al. Outcomes of SOT Recipients With COVID-19 in Different Eras of COVID-19 Therapeutics. *Transplant Direct.* 2022;8(1):e1268.
273. Salazar E, Perez KK, Ashraf M, et al. Treatment of Coronavirus Disease 2019 (COVID-19) Patients with Convalescent Plasma. *Am J Pathol.* 2020;190(8):1680-1690.
274. Sampaio PPN, Ferreira RM, de Albuquerque FN, et al. Rescue Venoarterial Extracorporeal Membrane Oxygenation After Cardiac Arrest in COVID-19 Myopericarditis: A Case Report. *Cardiovasc Revasc Med.* 2021;28S:57-60.
275. Schenker C, Hirzel C, Walti LN, et al. Convalescent plasma and remdesivir for protracted COVID-19 in a patient with chronic lymphocytic leukaemia: a case report of late relapse after rapid initial response. *Br J Haematol.* 2022;196(3):e27-e29.
276. Schreiber A, Elango K, Hong K, Ahsan C. Cardiac transplant recipient with COVID-19 induced acute hypoxic respiratory failure: a case report. *Eur Heart J Case Rep.* 2021;5(6):ytab217.
277. Schwartz SP, Thompson P, Smith M, et al. Convalescent Plasma Therapy in Four Critically Ill Pediatric Patients With Coronavirus Disease 2019: A Case Series. *Crit Care Explor.* 2020;2(10):e0237.
278. Senefeld JW, Johnson PW, Kunze KL, et al. Access to and safety of COVID-19 convalescent plasma in the United States Expanded Access Program: A national registry study. *PLoS Med.* 2021;18(12):e1003872.
279. Sepulcri C, Dentone C, Mikulska M, et al. The Longest Persistence of Viable SARS-CoV-2 With Recurrence of Viremia and Relapsing Symptomatic COVID-19 in an Immunocompromised Patient-A Case Study. *Open Forum Infect Dis.* 2021;8(11):ofab217.
280. Sethi SM, Ahmed AS, Hanif S, Aqeel M, Zubairi ABS. Subcutaneous emphysema and pneumomediastinum in patients with COVID-19 disease; case series from a tertiary care hospital in Pakistan. *Epidemiol Infect.* 2021;149:e37.
281. Shah M, Kakar A, Gogia A, Langer S. Convalescent plasma, cytomegalovirus infection, and persistent leukopenia in COVID-19 recovery phase: What is the link? *J Postgrad Med.* 2021;67(2):100-102.
282. Shaikh DH, Patel H, Makker J, Badipatla K, Chilimuri S. Colonic Ileus, Distension, and Ischemia Due to COVID-19-Related Colitis: A Case Report and Literature Review. *Cureus.* 2021;13(2):e13236.

 © 2022 Senefeld JW et al.

283. Shankar R, Radhakrishnan N, Dua S, et al. Convalescent plasma to aid in recovery of COVID-19 pneumonia in a child with acute lymphoblastic leukemia. *Transfus Apher Sci.* 2021;60(1):102956.
284. Shankar V, Dhar P, George J, Sharma A, Raj A. Eclampsia and posterior reversible encephalopathy syndrome in a parturient complicated by SARS COVID-19 pneumonia. *Braz J Anesthesiol.* 2021;71(5):576-578.
285. Sharma DJ, Deb A, Sarma P, Mallick B, Bhattacharjee P. Comparative Safety and Efficacy of Remdesivir Versus Remdesivir Plus Convalescent Plasma Therapy (CPT) and the Effect of Timing of Initiation of Remdesivir in COVID-19 Patients: An Observational Study From North East India. *Cureus.* 2021;13(11):e19976.
286. Shen C, Wang Z, Zhao F, et al. Treatment of 5 Critically Ill Patients With COVID-19 With Convalescent Plasma. *JAMA.* 2020;323(16):1582-1589.
287. Shields AM, Anantharachagan A, Arumugakani G, et al. Outcomes following SARS-CoV-2 infection in patients with primary and secondary immunodeficiency in the UK. *Clin Exp Immunol.* 2022;209(3):247-258.
288. Sobelman CS, Valentine SL, Kremer T. Management of COVID-19 in an adolescent demonstrates lasting effects of extreme prematurity on pulmonary function. *Respir Med Case Rep.* 2021;33:101394.
289. Soleimani Z, Soleimani A. ADRS due to COVID-19 in midterm pregnancy: successful management with plasma transfusion and corticosteroids. *J Matern Fetal Neonatal Med.* 2022;35(16):3040-3043.
290. Spinicci M, Mazzoni A, Borchi B, et al. AIDS patient with severe T cell depletion achieved control but not clearance of SARS-CoV-2 infection. *Eur J Immunol.* 2022;52(2):352-355.
291. Spinicci M, Mazzoni A, Coppi M, et al. Long-term SARS-CoV-2 Asymptomatic Carriage in an Immunocompromised Host: Clinical, Immunological, and Virological Implications. *J Clin Immunol.* 2022;42(7):1371-1378.
292. Steiner S, Schwarz T, Corman VM, et al. SARS-CoV-2 T Cell Response in Severe and Fatal COVID-19 in Primary Antibody Deficiency Patients Without Specific Humoral Immunity. *Front Immunol.* 2022;13:840126.
293. Swenson E, Wong LK, Jhaveri P, et al. Active surveillance of serious adverse events following transfusion of COVID-19 convalescent plasma. *Transfusion.* 2022;62(1):28-36.
294. Szwebel TA, Veyer D, Robillard N, et al. Usefulness of Plasma SARS-CoV-2 RNA Quantification by Droplet-based Digital PCR to Monitor Treatment Against COVID-19 in a B-cell Lymphoma Patient. *Stem Cell Rev Rep.* 2021;17(1):296-299.
295. Taha Y, Wardle H, Evans AB, et al. Persistent SARS-CoV-2 infection in patients with secondary antibody deficiency: successful clearance following combination casirivimab and imdevimab (REGN-COV2) monoclonal antibody therapy. *Ann Clin Microbiol Antimicrob.* 2021;20(1):85.
296. Tan L, Kang X, Zhang B, et al. Plasma therapy cured a COVID-19 patient with long duration of viral shedding for 49 days: The clinical features, laboratory tests, plasma therapy, and implications for public health management. *MedComm (2020).* 2020;1(1):77-80.
297. Thabrani A, Hadi WS, Thobari JA, et al. Convalescent plasma as a treatment modality for Coronavirus Disease 2019 in Indonesia: A case reports. *Ann Med Surg (Lond).* 2021;66:102444.
298. Thomopoulos TP, Rosati M, Terpos E, et al. Kinetics of Nucleocapsid, Spike and Neutralizing Antibodies, and Viral Load in Patients with Severe COVID-19 Treated with Convalescent Plasma. *Viruses.* 2021;13(9).
299. Thota DR, Ray B, Hasan M, Sharma K. Cryptococcal Meningoencephalitis During Convalescence From Severe COVID-19 Pneumonia. *Neurohospitalist.* 2022;12(1):96-99.
300. Tirnea L, Bratosin F, Vidican I, et al. The Efficacy of Convalescent Plasma Use in Critically Ill COVID-19 Patients. *Medicina (Kaunas).* 2021;57(3).
 **© 2022 Senefeld JW et al.**

301. Tran DH, Peng CC, Wolde-Rufael DA, et al. A successful case of extracorporeal membrane oxygenation for COVID-19: walking home without oxygen supplementation. *J Community Hosp Intern Med Perspect.* 2021;11(4):480-484.
302. Tremblay D, Seah C, Schneider T, et al. Convalescent Plasma for the Treatment of Severe COVID-19 Infection in Cancer Patients. *Cancer Med.* 2020;9(22):8571-8578.
303. Trimarchi H, Gianserra R, Lampo M, Monkowski M, Lodolo J. Eculizumab, SARS-CoV-2 and atypical hemolytic uremic syndrome. *Clin Kidney J.* 2020;13(5):739-741.
304. Umapathi T, Quek WMJ, Yen JM, et al. Encephalopathy in COVID-19 patients; viral, parainfectious, or both? *eNeurologicalSci.* 2020;21:100275.
305. Valentini R, Fernandez J, Riveros D, et al. [Convalescent plasma as a therapy for severe COVID-19 pneumonia]. *Medicina (B Aires).* 2020;80 Suppl 6:9-17.
306. Van Damme KFA, Tavernier S, Van Roy N, et al. Case Report: Convalescent Plasma, a Targeted Therapy for Patients with CVID and Severe COVID-19. *Front Immunol.* 2020;11:596761.
307. van Oers NSC, Hanners NW, Sue PK, et al. SARS-CoV-2 infection associated with hepatitis in an infant with X-linked severe combined immunodeficiency. *Clin Immunol.* 2021;224:108662.
308. Vlachogianni G, Hassapopoulou-Matamis H, Politis C, Fylaki E, Mentis A. A case of COVID-19 Convalescent Plasma Donation in Greece: Directed donation for compassionate use in the donor's critically ill father. *Transfus Clin Biol.* 2020;27(4):269-270.
309. Wang B, Van Oekelen O, Mouhieddine TH, et al. A tertiary center experience of multiple myeloma patients with COVID-19: lessons learned and the path forward. *J Hematol Oncol.* 2020;13(1):94.
310. Wang M, Yang X, Yang F, et al. Convalescent plasma therapy in critically ill coronavirus disease 2019 patients with persistently positive nucleic acid test, case series report. *Medicine (Baltimore).* 2020;99(36):e21596.
311. Wei B, Hang X, Xie Y, et al. Long-term positive severe acute respiratory syndrome coronavirus 2 ribonucleic acid and therapeutic effect of antivirals in patients with coronavirus disease: Case reports. *Rev Soc Bras Med Trop.* 2020;53:e20200372.
312. Weinbergerova B, Demel I, Visek B, et al. Successful early use of anti-SARS-CoV-2 monoclonal neutralizing antibodies in SARS-CoV-2 infected hematological patients - A Czech multicenter experience. *Hematol Oncol.* 2022;40(2):280-286.
313. Weinbergerova B, Mayer J, Kabut T, et al. Successful early treatment combining remdesivir with high-titer convalescent plasma among COVID-19-infected hematological patients. *Hematol Oncol.* 2021;39(5):715-720.
314. Werthman-Ehrenreich A. Mucormycosis with orbital compartment syndrome in a patient with COVID-19. *Am J Emerg Med.* 2021;42:264 e265-264 e268.
315. Whang SN, Shah VD, Pu L, et al. Convalescent Plasma for COVID-19: A Single Center Prospective Experience with Serial Antibody Measurements and Review of the Literature. *Pathogens.* 2022;11(9).
316. Wirz OF, Roltgen K, Stevens BA, et al. Use of Outpatient-Derived COVID-19 Convalescent Plasma in COVID-19 Patients Before Seroconversion. *Front Immunol.* 2021;12:739037.
317. Wright Z, Bersabe A, Eden R, Bradley J, Cap A. Successful Use of COVID-19 Convalescent Plasma in a Patient Recently Treated for Follicular Lymphoma. *Clin Lymphoma Myeloma Leuk.* 2021;21(1):66-68.
318. Wu Y, Hong K, Ruan L, et al. Patients with Prolonged Positivity of SARS-CoV-2 RNA Benefit from Convalescent Plasma Therapy: A Retrospective Study. *Virol Sin.* 2020;35(6):768-775.
319. Xu TM, Lin B, Chen C, Liu LG, Xue Y. Non-optimal effectiveness of convalescent plasma transfusion and hydroxychloroquine in treating COVID-19: a case report. *Virol J.* 2020;17(1):80.

© 2022 Senefeld JW et al.

320. Yang B, Fulcher JA, Ahn J, et al. Clinical Characteristics and Outcomes of Coronavirus Disease 2019 Patients Who Received Compassionate-Use Leronlimab. *Clin Infect Dis.* 2021;73(11):e4082-e4089.
321. Yang B, Yang J, Zhou L, et al. Inflammatory cytokine depletion in severe coronavirus disease 2019 infectious pneumonia: A case report. *Medicine (Baltimore).* 2020;99(49):e23449.
322. Yaqoub S, Ahmad S, Mansouri Z, et al. Management of life-threatening acute respiratory syndrome and severe pneumonia secondary to COVID-19 in pregnancy: A case report and literature review. *Clin Case Rep.* 2021;9(1):137-143.
323. Yatam Ganesh S, Nachimuthu N. Treatment Experience With Inhaled Corticosteroids in Combination with Remdesivir and Dexamethasone Among COVID-19 Patients Admitted to a Rural Community Hospital: A Case Series. *Cureus.* 2020;12(11):e11787.
324. Ye M, Fu D, Ren Y, et al. Treatment with convalescent plasma for COVID-19 patients in Wuhan, China. *J Med Virol.* 2020;92(10):1890-1901.
325. Yi SG, Rogers AW, Saharia A, et al. Early Experience With COVID-19 and Solid Organ Transplantation at a US High-volume Transplant Center. *Transplantation.* 2020;104(11):2208-2214.
326. Yokoyama APH, Wendel S, Bonet-Bub C, et al. COVID-19 convalescent plasma cohort study: Evaluation of the association between both donor and recipient neutralizing antibody titers and patient outcomes. *Transfusion.* 2021;61(8):2295-2306.
327. Yulianti M, Johan C, Singh G, Tenda ED, Herikurniawan H, Wijaya I. Effectiveness of Convalescent Plasma Therapy in Treating COVID-19: an Evidence-based Case Report. *Acta Med Indones.* 2021;53(4):497-504.
328. Zeng H, Wang D, Nie J, et al. The efficacy assessment of convalescent plasma therapy for COVID-19 patients: a multi-center case series. *Signal Transduct Target Ther.* 2020;5(1):219.
329. Zhang B, Liu S, Tan T, et al. Treatment With Convalescent Plasma for Critically Ill Patients With Severe Acute Respiratory Syndrome Coronavirus 2 Infection. *Chest.* 2020;158(1):e9-e13.
330. Zhang L, Pang R, Xue X, et al. Anti-SARS-CoV-2 virus antibody levels in convalescent plasma of six donors who have recovered from COVID-19. *Aging (Albany NY).* 2020;12(8):6536-6542.
331. Zhang LB, Pang RR, Qiao QH, et al. Successful recovery of COVID-19-associated recurrent diarrhea and gastrointestinal hemorrhage using convalescent plasma. *Mil Med Res.* 2020;7(1):45.
332. Zhang LL, Liu Y, Guo YG, et al. Convalescent Plasma Rescued a Severe COVID-19 Patient with Chronic Myeloid Leukemia Blast Crisis and Myelofibrosis. *Turk J Haematol.* 2021;18(1):74-76.
333. Zimmerli A, Monti M, Fenwick C, et al. Case Report: Stepwise Anti-Inflammatory and Anti-SARS-CoV-2 Effects Following Convalescent Plasma Therapy With Full Clinical Recovery. *Front Immunol.* 2021;12:613502.
334. Zimmermann J, Glueck OM, Fertmann JM, et al. COVID-19 in Recent Lung Transplant Recipients: Clinical Outcomes and Management Strategies. *Transplant Proc.* 2022;54(6):1504-1516.
335. Zlamal M, Stechovska K, Holub M. A case of severe course of COVID-19 treated with experimental therapy. *Cas Lek Cesk.* 2020;159(5):181-184.

 **© 2022 Senefeld JW et al.**
